# Gene-vegetarianism interactions in calcium, testosterone, and eGFR identified in genome-wide analysis across 30 biomarkers

**DOI:** 10.1101/2022.10.21.22281358

**Authors:** Michael Francis, Kaixiong Ye

## Abstract

Existing vegetarian cohort studies have not considered the effects of genetic differences on health outcomes. First, we reduced inconsistencies among self-identified vegetarians in UK Biobank by utilizing two dietary surveys. Vegetarians were matched 1:4 with nonvegetarians for traditional association analyses, revealing significant effects of vegetarianism in 15/30 biomarkers. Cholesterol measures plus Vitamin D were significantly lower in vegetarians, while triglycerides were higher. A genome-wide association study revealed no genome-wide significant (GWS) associations with vegetarianism status. We performed genome-wide gene-vegetarianism interaction analyses for 30 biomarker traits (N=147,253). We detected a GWS interaction in calcium at rs72952628 (*P*=4.47×10^−8^). rs72952628 is in *MMAA*, a B_12_ metabolism gene; B_12_ has high deficiency potential in vegetarians. Gene-based interaction tests revealed two significant genes, *RNF168* in testosterone (*P*=1.45×10^−6^) and *DOCK4* in eGFR (*P*=6.76×10^−7^), which have previously been associated with testicular and renal traits, respectively. These findings indicate genotype can influence biomarker levels across vegetarians.

## Introduction

Vegetarianism is a superordinate term for a variety of animal-restricted dietary practices, typically referring to lacto-ovo vegetarianism, which permits plant-based food, dairy, and eggs, and excludes meat, fish, and seafood ^1^. Estimates indicate that in Western countries, interest in and adherence to plant-based diets have increased over the past decade ^2-4^. This has occurred for several reasons, including health benefits, taste preferences, ethical concerns with slaughtering animals and factory farming, environmental concerns related to pollution and greenhouse gas emissions, and perceived moral accreditation ^4,5^. It is now typical for nutrition professionals to recommend vegetarianism to the general public *en masse* ^4,6,7^.

Recent large meta-analyses have found health benefits associated with vegetarianism, including improved blood lipids, and reductions in body mass index (BMI), heart disease, type 2 diabetes, and certain cancers, though no significant differences have been found in all-cause mortality ^1,8-10^. As the authors of these meta-analyses have pointed out, many vegetarian observational studies are confounded by information and selection biases ^1,8,9^. We have identified the most commonly occurring biases in previous studies, and hope to alleviate these issues in the current analysis.

Heterogeneous and imprecise questionnaire design in defining vegetarianism is a recurring source of information bias in previous cohort studies. Self-reported vegetarians are known to vary widely in their strictness of following a diet that contains no meat or fish ^11^. There are issues of trustworthiness in dietary questionnaire response, particularly in the direction of over-reporting “healthy” behaviors ^12,13^. One way to greatly improve the reliability of survey measurement is to integrate multiple dietary assessments to define intake, as opposed to using a single survey instance ^14-16^.

Vegetarians may also be more health conscious in general than omnivores, which introduces a selection bias sometimes called the “healthy user effect” ^17^. When lifestyle factors adjacent to vegetarianism are not properly controlled for, it can lead to overestimating the effect of vegetarianism. One outstanding example of this bias in vegetarianism studies, specifically those conducted in the US, has been an over-generalization of results from Seventh Day Adventists (SDAs) ^1,8,9,18,19^, who in addition to vegetarianism, observe many healthy lifestyle practices, such as increased emphasis on exercise, and avoidance of all tobacco, drugs, and alcohol. Meta-analyses reveal that non-SDA vegetarians consistently show less health benefits than SDAs ^1,8,9^. Matching participants on relevant lifestyle characteristics can help alleviate this issue ^20^. Large-scale databases like the UK Biobank (UKB) offer the opportunity to match vegetarians to omnivores while still maintaining sufficient power.

In addition to the aforementioned biases, there has been no consideration of genetics in large epidemiological studies of vegetarianism. Genetics and ancestry are known to play an important role in metabolic processes, i.e., nutrigenetics ^21^. There are two aspects of genetics we consider in this analysis. First, we examined whether there is a genetic component to vegetarianism status. Heritable components have been associated with plant-eating dietary preferences ^22^. Significant variants have been associated with quantitative measures of plant-eating ^23,24^, though a recent GWAS of vegetarianism found none ^25^.

Perhaps more meaningful than finding a genetic predisposition towards certain dietary habits, is identifying how a diet relates to the genetic background of an individual. Gene-diet interactions (GDI) are a type of gene by environment interaction (GEI or GxE), where diet is the environmental exposure. GDIs have been identified using exposures of overall dietary patterns for some serum biomarkers ^26^, but no gene-vegetarianism interactions have been reported to-date.

This study consists of four parts. First, by utilizing both dietary surveys administered to UKB participants, we defined a high-quality cohort of vegetarians that were most likely to be vegetarian at the time of the serum biomarker collection. Participants’ vegetarianism status was defined based on four criteria: self-identified as vegetarian on the first 24-hour recall survey (24HR), did not eat meat or fish on the first 24HR, did not eat meat or fish on the initial assessment (IA), and had no major dietary changes over the past 5 years. Second, we estimated exposure effects of vegetarianism in a matched sample of vegetarian and nonvegetarian Europeans across 30 serum biomarkers for diabetes, cancer, cardiovascular, skeletal, renal, and liver diseases. Third, we performed a genome-wide association study (GWAS) to search for variants that may explain vegetarianism preference on a genetic level. Finally, we performed the first genome-wide gene-diet interaction study (GWIS) of vegetarianism across 30 biomarkers, and identified genome-wide significant gene-vegetarianism interactions on calcium, testosterone, and estimated glomerular filtration rate (eGFR). This study provides the first large-scale evidence that genetic factors play a role in differential health outcomes among vegetarians.

## Methods

### Ethics

UK Biobank (UKB) approved the use of medical and genetic data under Project ID 48818. UKB received ethical approval from the research ethics committee (reference ID 11/ NW/0382) and obtained written informed consent from participants. This project using existing UKB data was approved by the Institutional Review Board (IRB) at the University of Georgia. Data analysis was performed on a University of Georgia high performance computing server with strict data protection protocols and two-factor authentication. Participants that withdrew their consent as of Feb. 22^nd^, 2022 were removed (N=114).

### Vegetarianism designation

UKB is a prospective cohort study containing > 500,000 participants between ages 40 and 70, who were recruited in England, Scotland, and Wales between 2006 and 2010. All UKB Field and Category references can be located in their publicly available data dictionary (https://biobank.ndph.ox.ac.uk/ukb/). Dietary data was collected in two separate surveys. All participants answered the touchscreen questionnaire on “Diet” during their initial visit to the Assessment Centre (Category ID 100052). Additionally, the “Diet by 24-hour recall” section of the “Online follow-up questionnaire” (24HR; Category ID 100090) was administered to a subset of participants on a voluntary basis, during the last phase of the initial assessment (Instance 0; N=70,689) and subsequently via email, for a total of up to five rounds between April 2009 and June 2012 (N=210,966 unique participants) ^27^.

Our goal was to identify a subset of participants most likely to have consistently followed a strict vegetarian or vegan diet at the time of the blood draw for biomarker measurement at the initial assessment. Vegetarians and vegans were grouped together in all analyses because of the limited number of vegans. In this study vegetarians/vegans were defined as meeting all four of the following criteria. First, in a participant’s first instance of taking the 24HR, in response to the question “Do you routinely follow a special diet?” (Field 20086), they must have indicated “Vegetarian diet (no meat, no poultry and no fish)” and/or “Vegan diet”. Next, on that same first instance of the 24HR, a participant must have also answered “No” to “Did you eat any meat or poultry yesterday? Think about curry, stir-fry, sandwiches, pie fillings, sausages/burgers, liver, pate or mince,” (Field 103000) as well as to “Did you eat any fish or seafood yesterday? e.g. at breakfast, takeaway with chips, smoked fish, fish pate, tuna in sandwiches.” (Field 103140). Third, on the initial dietary assessment survey, participants must have answered “Never” to all of the questions asking how often meat or fish was eaten (Fields 1329, 1339, 1349, 1359, 1369, 1379, and 1389). Finally, on the initial assessment, participants must have answered “No” to the question “Have you made any major changes to your diet in the last 5 years?” (Field 1538).

### Participants

Only participants designated as having European (EUR) ancestry by the Pan UKBB project ^28^ were used in analyses to avoid population stratification. Participants were removed on the following quality control parameters: mismatches between self-reported and genetic sex, poor quality genotyping as flagged by UKB, sex chromosome aneuploidy, and/or having a high degree of genetic kinship (ten or more third-degree relatives identified). Additionally, we removed the minimum number of participants to eliminate all related pairs.

### Phenotype data

Continuous serum biochemistry markers were obtained from Category 17518. Oestradiol and rheumatoid factor (Fields 30800, 30820) were excluded due to limited participant data (<20% of participants). Glucose (Field 30740) was excluded due to inconsistencies in fasting times among participants and a limited number of participants with fasting times larger than 7h. Total cholesterol, LDL-C, and apolipoprotein B were divided by an adjustment factor (0.749, 0.684, and 0.719, respectively) for those who self-reported use of statins ^29^. Three derived traits were also included. Free testosterone was calculated with the Vermeulen equation ^30^, bioavailable testosterone was calculated with the Morris equation ^31^, and the CKD-EPI Creatinine-Cystatin Equation (2021) was used to calculate estimated glomerular filtration rate (eGFR) ^32^. All traits were transformed using direct rank-based inverse normal transformation (RINT) with random separation of ties.

### Genotype data

Genotype data was provided with initial quality control and imputation with Haplotype Reference Consortium (HRC) and 1000 Genomes variants by UKB (v3) as previously described ^33^. Additionally, we removed variants with imputation quality score (INFO) < 0.5, minor allele frequency (MAF) < 1%, missing genotype per individual > 5%, missing genotype per variant > 2%, or Hardy-Weinberg equilibrium (HWE) *P* < 1×10^−6^. Variant filtering and genotype file format conversions were performed using PLINK2 alpha-v2.3 ^34^. After quality control, 7,918,739 variants remained. All genomic positions in this study refer to the Genome Reference Consortium Human Build 37 (GRCh37; hg19).

### Sample matching and estimating vegetarianism effects

To select controls for the analysis of vegetarianism exposure effects, cases were pre-processed to match four controls with nearest-neighbor (greedy) matching without replacement, using MatchIt v4.4.0.9004 ^35^. Matching distance between participants was calculated by general linearized model, and was performed on the basis of age, sex, body mass index (BMI; kg/m^2^), alcohol use frequency (<3 drinks/week or ≥ 3 drinks/week), previous smoking status (yes/no), current smoking status (yes/no), standardized Townsend deprivation index, and the first five genetic principal components. Sixteen vegetarians with incomplete covariate information were excluded, leaving a total of 2,312 EUR vegetarians.

Matching was followed by regression using the same vector of covariates; using the same covariates is recommended to reduce the dependence of regression estimates on modeling decisions, increase precision, reduce bias, and increase robustness of the effect estimate ^20,36^. Vegetarianism effects estimates were computed by linear model (no interaction) in R v4.2.1 with cluster-robust standard errors implemented by Sandwich v3.0-2 ^37^. Sex-stratified models of the same matched participants were also run. Forestplot v3.0.0 was used to make forest plots.

### Genome-wide association study of vegetarianism

Genome-wide association study (GWAS) was performed using Regenie v3.1.2 ^38^. Vegetarianism status as defined above was used as a binary trait. A whole genome regression model was fit at a subset of genetic markers from non-imputed UKB genotype calls. Variants used in model fitting were filtered in PLINK2 alpha-v2.3 ^34^ by these criteria: MAF < 0.01, minor allele count < 100, genotype missingness < 0.1, HWE exact test *P*-value < 10^−15^. Covariates used for both model fitting and GWAS (standard model) were age, sex, genotyping batch, alcohol use frequency, previous smoker (yes/no), current smoker (yes/no), standardized Townsend deprivation index, and the first ten genetic principal components as provided by UKB. A BMI-adjusted model was separately run to compare sensitivity models for confounding effects of BMI. Individuals with missing phenotype or covariate data were excluded. Firth correction was applied for *P* < 0.01 to reduce the bias in the maximum-likelihood estimates, using a penalty term from Jeffrey’s Prior as described previously ^39^. Genomic control (λ) was calculated for *P*-values using the median of the chi-squared test statistics divided by the expected median of the chi-squared distribution.

### Genome-wide interactions with vegetarianism

GEM (Gene–Environment interaction analysis in Millions of samples) v1.4.3 ^40^ was used to perform genome-wide interaction study (GWIS) of 30 continuous biomarker traits, using vegetarianism status as a binary exposure variable. Covariates used in GWIS were the same as in GWAS above. Individuals with missing phenotype or covariate data were excluded. Robust standard error correction as implemented by GEM was performed in all models to correct for initially observed heteroskedasticity. Initial genomic control (λ) using non-robust standard errors ranged from 0.895-1.255, likely due to heteroskedasticity, therefore robust standard errors as implemented by GEM were used for all models. Interaction effects and *P*-values refer to 1 degree of freedom (1df) tests of variant effects in a gene-environment interaction model with. Marginal effects refer to the association between genetic effects and phenotype in a model without interaction. A BMI-adjusted model was separately run for all traits. Correlation between standard and BMI-adjusted models was assessed using a two-sided Spearman’s rank correlation coefficient. We did not interpret 2df joint effects, though they are available in our public summary statistics.

Variants were queried for associations with gene expression levels in tissues using Genotype-Tissue Expression (GTEx) Project (GTEx) Analysis Release V8 (dbGaP Accession phs000424.v8.p2). Fastman v0.1.0 was used to generate Manhattan plots ^41^. Hudson (v1.0.0) was used to create interactive Manhattan plots ^42^.

### Gene-based interaction analyses

MAGMA v1.10 ^43^ was used to aggregate *P*-values from individual variant associations (for vegetarianism) and 1 df interactions (for 30 biomarkers) to genic regions. Variants were mapped to a total of 18,208 genes using a window of +2 Kb upstream and -1 Kb downstream of the transcription start and stop sites to allow for the inclusion of nearby regulatory variants. Linkage disequilibrium was estimated using reference data from the 1000 Genomes British population of EUR ancestry. The “multi model” method of aggregation was used to apply both “mean” and “top” models and select the one with the best fit ^44^.

## Results

### Identifying a reliable sample of vegetarians

We searched the UK Biobank (UKB) to find a reliable subset of participants that were most likely to be vegetarian at the initial assessment (IA), when blood samples were collected for biomarker measurement. Two separate dietary surveys were part of UKB data collection, one at the IA which was taken by all UKB participants (N=502,413), and one in the 24-hour recall survey (24HR), which was administered after the IA in five waves or “instances”, between April 2009 and June 2012 (N=210,967 unique participants; Figure 1A). Participants were invited to take the 24HR between one and five times on a voluntary basis (Figure 1B).

**Figure 1.**
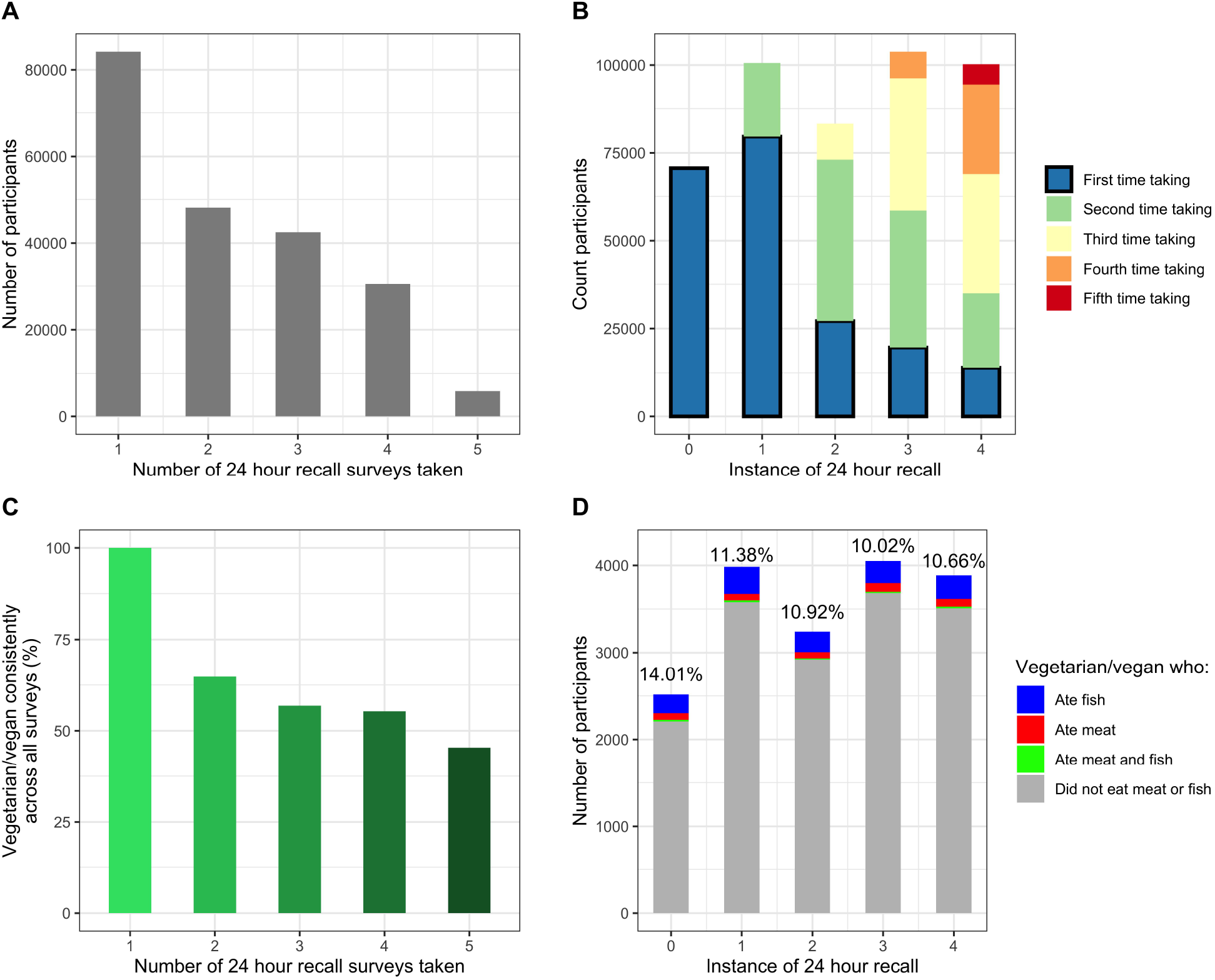
Identifying vegetarians. (A) Participants were invited to take the 24-hour recall survey (24HR) between one and five times on a voluntary basis. (B) The 24HR we considered were restricted to the first-time participants took that survey, because this was the closest time point to the blood draw of biomarkers at the initial assessment. (C) For participants who self-identified as vegetarian in at least one 24HR and took multiple 24HRs, they were less likely to self-identify as vegetarian in all surveys. (D) The percentage of vegetarians who indicated eating meat or fish on the same 24HR as identifying as vegetarian ranged between 10.02-14.01%.

We used four criteria to designate participants as vegetarian. Our first criterion was whether a participant indicated they routinely followed a vegetarian or vegan diet; this question was only asked on the 24HR. A total of 9,115 participants self-identified in at least one 24HR that they were either vegetarian or vegan (hereafter collectively referred to as “vegetarian” due to a small number of vegans). We found an inverse relationship between the percentage of participants who consistently self-identified as vegetarian in every 24HR they took, and the number of times participants took the 24HR (Figure 1C). For example, of the participants who identified as vegetarian at least once and participated in two instances of the 24HR, only 64.8% self-identified as vegetarian both times (1,380 of 2,130); for participants that took the 24HR in all five instances, only 45.4% consistently identified as vegetarian every time (168 of 370). Because we were interested in biomarker levels at the IA time point, we considered identification as vegetarian in the earliest instance of the 24HR as sufficient for passing this criterion.

Next, the 24HR asked whether a participant ate meat or fish yesterday. To find intra-survey discrepancies of vegetarianism status, we identified those who identified as vegetarian and also self-reported eating meat or fish on the same instance of the 24HR. The percentage of these participants ranged from 10.02-14.01% per survey instance (Figure 1D). Participants who reported eating meat on their first 24HR were disqualified from our “reliable” vegetarianism status. Similarly, in our third criterion, we disqualified vegetarians who did not answer “Never” to questions asking their frequency of eating meat or fish on the IA.

Lastly, because of the high amount of dietary fluctuation we found in self-identified vegetarians, we also required vegetarians to have answered “No” to a question on the IA which asked whether they had any major dietary changes over the past five years. Overall, out of 9,115 UKB participants who self-identified as vegetarian or vegan on at least one 24HR, we found 3,205 met our criteria of not reporting eating meat on IA, nor on the nearest 24HR to the blood draw time point, plus had not reported major dietary changes (Table 1).

**Table 1.**
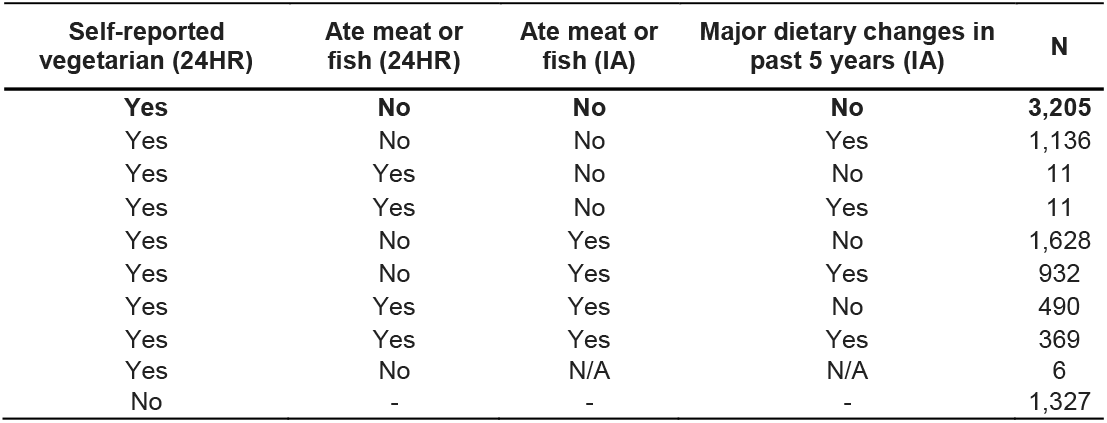
Selecting high quality vegetarians. Vegetarians were selected on four criteria: self-identifying as vegetarian on the first 24-hour recall survey (24HR) that they participated in, not eating meat or fish on the first 24HR, not eating meat or fish on the initial assessment (IA), and no major dietary changes over the past 5 years. A total of 3,205 UK Biobank participants met these criteria (top row, bold). This table shows counts of participants from all UK Biobank participants who took the 24HR (N = 210,967). After filtering by ancestry, the total of 3,205 was reduced to 2,312 European vegetarians, the number used in the analyses that follow.

### Sample matching and estimating vegetarianism effects on serum biomarkers

After quality controlling participants and keeping only those who were part of the largest ancestry group, European, using Pan UKBB designations ^28^, 2,328 vegetarians and 153,047 nonvegetarians remained (Supplementary Table 1). Raw (untransformed) values for 30 traits were plotted, and some exhibited apparent differences between vegetarians and nonvegetarians (Supplementary Figure 1). However, the covariates selected for our effects estimation model (age, sex, BMI, alcohol use frequency, previous smoker, current smoker, Townsend index, and the first five genetic principal components) were highly imbalanced between the two groups (Supplementary Table 2). For example, the average ages of nonvegetarians and vegetarians were 56.5 and 52.7 years, respectively. Similarly, nonvegetarians were 54.1% female, compared to 66.2% in vegetarians. The covariates with highest standardized mean differences (SMD) between the two groups were age (−0.482) and BMI (−0.501). Therefore, prior to estimating the effects of vegetarianism across the 30 traits, we matched each vegetarian to four nonvegetarians along these covariates. After matching, the absolute SMD (ASMD) in all model covariates was < 0.05 standard deviations (Supplementary Figure 2). The variance ratio of the distance of propensity scores between unmatched and matched vegetarians was improved from 2.1203 to 1.0216. Similarly, the maximum empirical cumulative density function (eCDF) difference (Kolmogorov-Smirnov statistic, *D*_*n*_) was improved from 0.3038 to 0.0013 (Supplementary Table 2). These measures indicate that covariate balance was achieved between matched vegetarians and nonvegetarians.

Participants were filtered for those who had complete covariate data. The standardized effect of vegetarianism was estimated across 30 serum biomarker traits with rank-based inverse normal transformation in 2,312 vegetarians and 9,248 matched nonvegetarians. Fifteen of these traits had significant effects at the Bonferroni corrected *P*-value threshold of 0.05/30 = 0.0017, while five additional trait effects were nominally significant (*P* < 0.05) (Figure 2, Supplementary Table 3). Effects of vegetarianism were significant and negative across all cholesterol measures, including total cholesterol, low-density lipoprotein cholesterol (LDL), high-density lipoprotein cholesterol (HDL), plus Apolipoproteins A and B (ApoA, ApoB); while lipoprotein (a) (Lp (a)) was nominally significant. A significant positive effect of vegetarianism was observed on triglycerides (*β* = 0.223; *P* = 4.0×10^−26^). Vegetarianism had a significant negative effect on the steroid hormone Vitamin D (*β* = -0.388; *P* = 2.1×10^−49^), and with the growth hormone-regulating insulin-like growth factor 1 (IGF-1). Sex-related hormone measures of testosterone (total T, bioavailable-T, and free-T) and sex hormone binding globulin (SHBG) were not significant in the combined nor sex-stratified effects estimation (Supplementary Figure 3).

**Figure 2.**
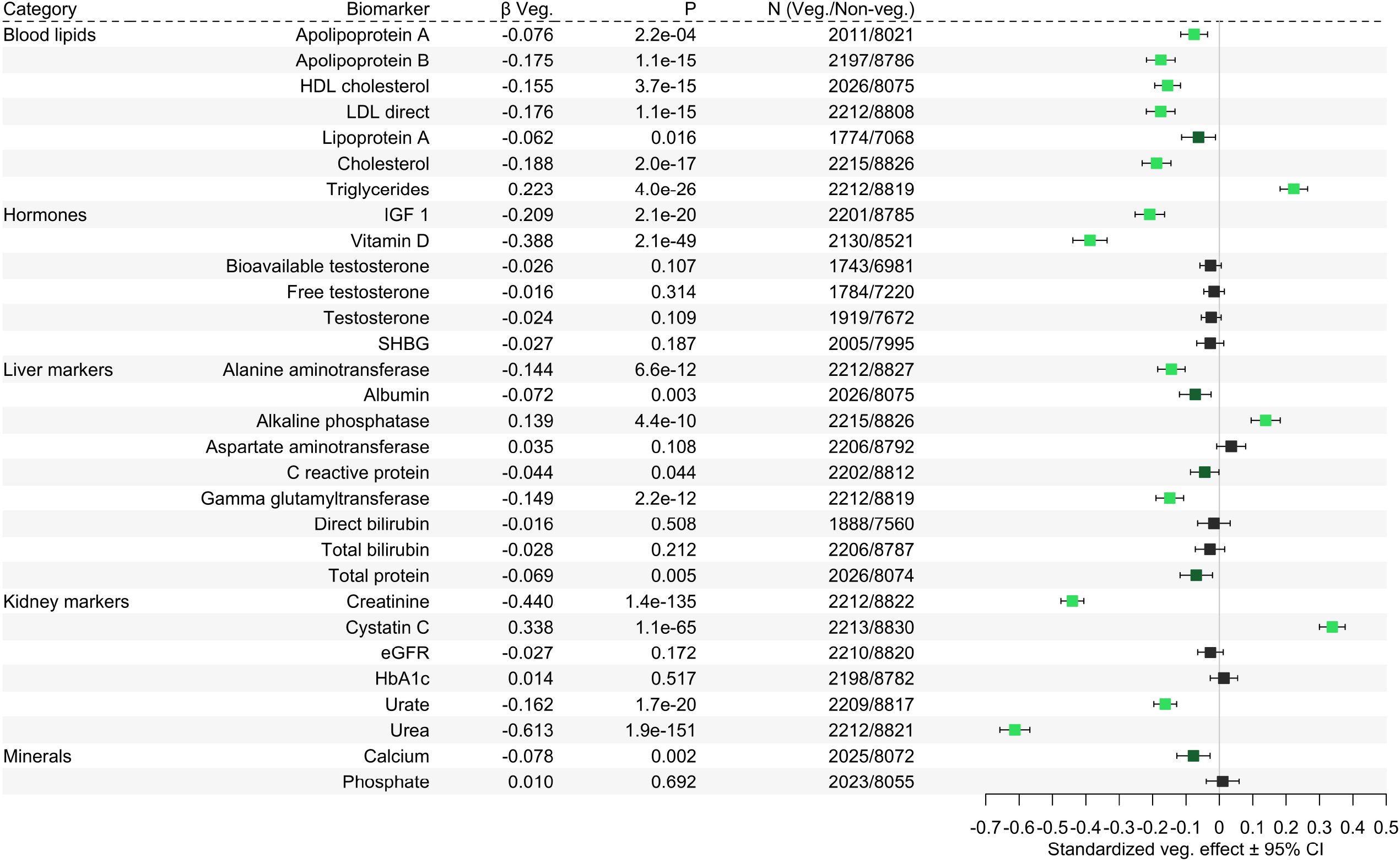
Forest plot of estimated vegetarianism effects. Vegetarians were matched 1:4 with nonvegetarians and effects of vegetarianism were estimated across thirty biomarkers. Error bars show 95% confidence intervals. Light green dots indicate Bonferroni-corrected significance (*P* < 0.0017), dark green dots show nominal significance (*P* < 0.05), and black dots are not significant.

Alanine aminotransferase (ALT) and gamma-glutamyl transferase (GGT) were associated with significant negative effects of vegetarianism, while a positive effect was observed with alkaline phosphatase (ALP). Effects on other liver-associated markers such as albumin, aspartate aminotransferase, C-reactive protein, direct bilirubin, total bilirubin, and total serum protein, were not significant. Kidney markers associated with protein metabolism and breakdown, such as creatinine, urate and urea, displayed negative effects from vegetarianism, while cystatin C was associated with a strong positive effect, and eGFR did not have a significant association. HbA1c (glycated haemoglobin) was not significantly associated. Vegetarianism had a negative effect on serum calcium that nearly reached the multiple testing threshold (*P* = 0.002), while phosphate was not significantly associated. Sex-stratified analysis revealed the effect of vegetarianism was driven by one sex in three traits. ApoA was significant only in males, ALP and Lp (a) were significant only in females; C-reactive protein was nearly significant in females (Supplementary Figure 3; Supplementary Table 3).

### Genome-wide association study of vegetarianism

A total of 7,918,739 variants were tested in a GWAS of 152,764 European UKB participants, using vegetarianism as a binary trait as defined in Table 1. In standard and BMI-adjusted models, *P*-values were highly correlated (*R* = 0.97; Supplementary Figure 4). Potential inflation from imbalanced case:control ratio (2,312 vegetarians, 152,764 nonvegetarians) was adjusted for by regenie (λ = 1.032 in both models). No associations with vegetarianism reached genome-wide significance (GWS; *P* < 5×10^−8^; Supplementary Figure 5). The two most significant variants in both models were indels, at 4:183448129_AT_A (*P*_standard_ = 1.65×10^−7^; *P*_adj-BMI_ = 1.36×10^−7^) and 11:870094_CG_C (*P*_standard_ = 1.61×10^−7^; *P*_adj-BMI_ = 2.10×10^−7^). GWAS *P*-values were aggregated into genic regions using MAGMA. No genes achieved statistical significance after multiple testing correction. The most significant genes in each model were *HLA-DPB1* (*P*_standard_ = 1.12×10^−5^; *P*_adj-BMI_ = 5.73×10^−5^) and *YWHAZ* (*P*_standard_ = 1.44×10^−4^; *P*_adj-BMI_ = 4.66×10^−5^).

### Genome-wide gene-vegetarianism interactions

#### Variant level

Gene-environment interactions using vegetarianism status (Table 1) as the environmental exposure was performed across 30 serum biomarker traits (*N* = 117,356-147,253) in standard and BMI-adjusted models (Supplementary Table 4). For each GWIS, 7,934,157 variants were tested for marginal effects, interaction effects (1 degree of freedom test), and joint main and interaction effects (2 degrees of freedom test). We were specifically interested in interaction effects and their corresponding *P*-values, as these most directly demonstrate the interaction of vegetarianism with genetic variants. Genomic control (λ) for robust *P*-values ranged from 0.985-1.024, indicating no issues of inflation (Supplementary Table 4).

Across the 30 traits analyzed for gene-vegetarianism interactions, only one variant was significant at GWS, and no variants reached significance at the stricter threshold corrected for the number of traits analyzed (1.67×10^−9^; Supplementary Figure 6; Supplementary Table 4). In calcium, interaction of rs72952628 (chr4:146,637,234) reached GWS in the standard model and nearly in the BMI-adjusted model (*P*-int_standard_ = 4.47×10^−8^; *P*-int _adj-BMI_ = 6.29×10^−8^; Figure 3A), while the marginal P-value was high (*P*-marginal_standard_ = 0.0269; *P*-marginal_adj-BMI_ *=* 0.0233), indicating predominately interaction effects at this locus. This variant is located in the intron of *C4orf51*, and in moderate linkage disequilibrium (*r*^*2*^: 0.605-0.719) with variants in exon 7 of *MMAA* (Figure 3B). In a genotype-stratified analysis, vegetarianism was associated with a 0.135-unit decrease [-0.180 to -0.0897] of serum calcium in homozygotes of the major allele (CC), but with a 0.298-unit increase [0.136 to 0.459] in heterozygotes (Figure 3C).

**Figure 3.**
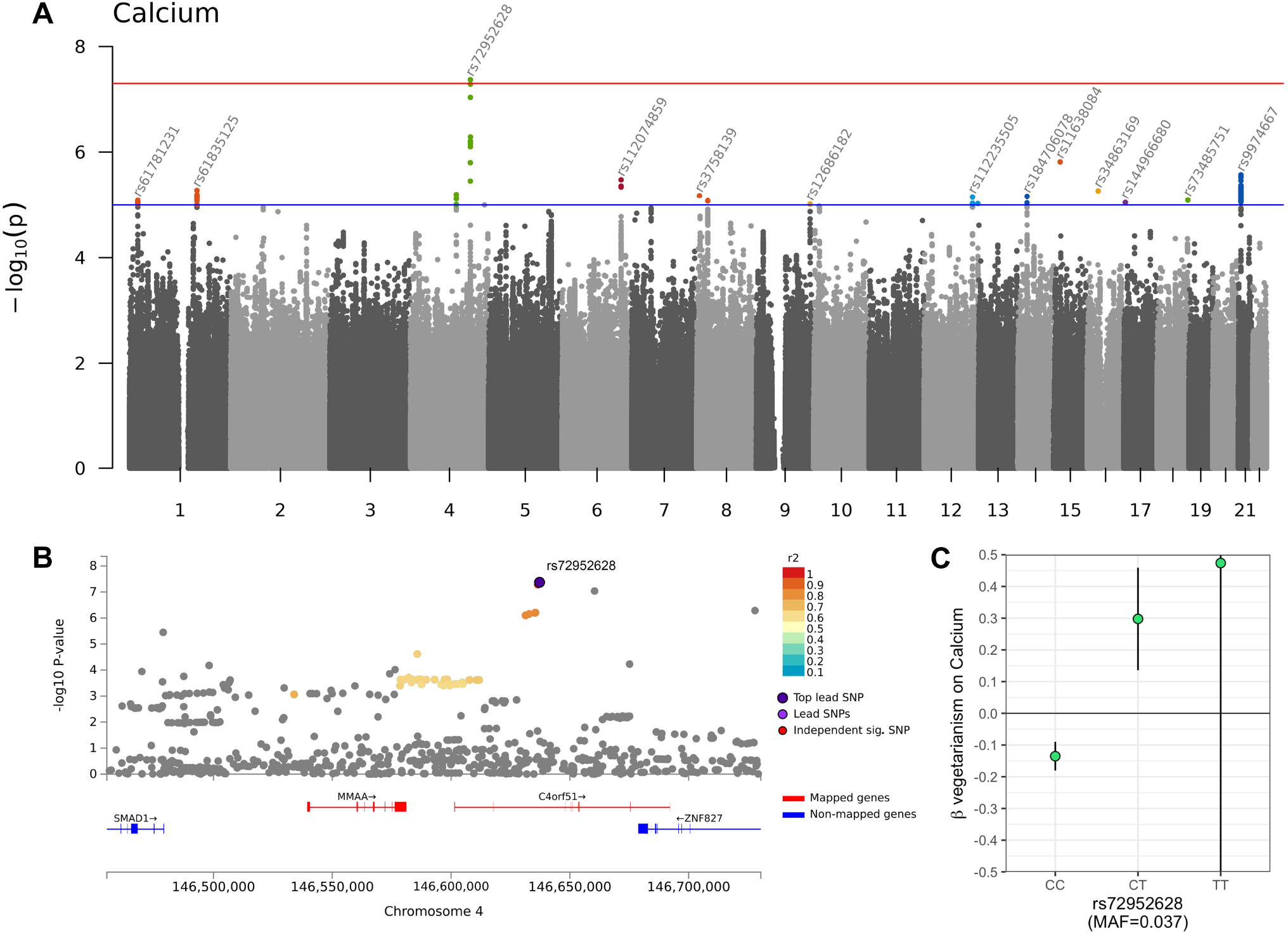
Calcium gene-vegetarianism interaction at rs72952628. (A) Manhattan plot of *P*-values for gene-vegetarianism interaction on calcium. One variant, rs72952628 (chr4:146,637,234), passed the genome-wide significance threshold of *P* < 5×10^−8^. (B) The regional Manhattan plot of rs72952628 shows that it is in linkage disequilibrium with variants in *C4orf51* and *MMAA*. (C) The effect of vegetarianism on calcium, stratified by genotype. The homozygous minor allele genotype, TT, has a large error bar because of its infrequency in the sample (n = 207). Error bars show 95% confidence intervals; units of calcium are SD.

*MMAA* encodes a GTPase involved in one-carbon metabolism of vitamin B_12_ (B_12_; also known as cobalamin). Specifically, it helps mediate the transport of cobalamin (Cbl) into mitochondria for the final steps of adenosylcobalamin (AdoCbl) synthesis. The most prominent cause of B_12_ deficiency is inadequate dietary intake, and this is especially common among vegetarians and vegans since the majority of dietary B_12_ is derived from animal sources ^45^. GTEx single-tissue expression Quantitative Trait Loci (eQTL) analysis for rs72952628 showed an exclusive and significant association with *MMAA* gene expression in four tissue types, and nearly reaching the GTEx multiple testing significance threshold in liver tissue (*P* = 4.77×10^−4^), where the heterozygote CT is consistently associated with higher *MMAA* expression (Supplementary Figure 7). Across the 54 tissues and cells examined by GTEx, *MMAA* had the highest expression in the liver (Supplementary Figure 8A).

There are fifteen or more gene products involved in B_12_ transport and processing ^45^. Of these, two have calcium-binding domains, cubilin, and CD320. In the distal ileum, binding of the IF-B_12_ complex to the cubilin receptor is calcium-dependent ^46^. However, a closer candidate in the B_12_ pathway for calcium involvement is CD320. In the liver, CD320 receptor mediates transcobalamin-bound B_12_ cellular uptake, a process which is Ca^2+^ dependent ^47^. This occurs in the same cells where MMAA is active in the mitochondria, including but not exclusive to liver cells.

### Gene level interactions

Interaction *P*-values of GWIS variants were aggregated into genic regions using MAGMA for each of the 30 biomarkers. Variants were mapped to 18,208 genes, making the significant *P*-value threshold corrected for the number of genes as (0.05 / 18,208 = 2.75×10^−6^), and that threshold additionally corrected for the number of traits as (2.75×10^−6^ / 30 = 9.15×10^−8^). Genomic control (λ) for these aggregated models ranged from 0.898-1.098 (Supplementary Figure 9; Supplementary Table 4). Two genes in two traits were significant at the threshold corrected for the number of genes: *RNF168* in total testosterone (*P*_standard_ = 1.45×10^−6^, *P*_adj-BMI_ = 1.03×10^−6^; Figure 4A), and *ZNF277* in eGFR (*P*_standard_ = 6.76×10^−7^, *P*_adj-BMI_ = 9.28×10^−6^; Figure 4B). No genes in the analysis were significant at the more conservative significance level corrected for the number of traits (Supplementary Table 4).

**Figure 4.**
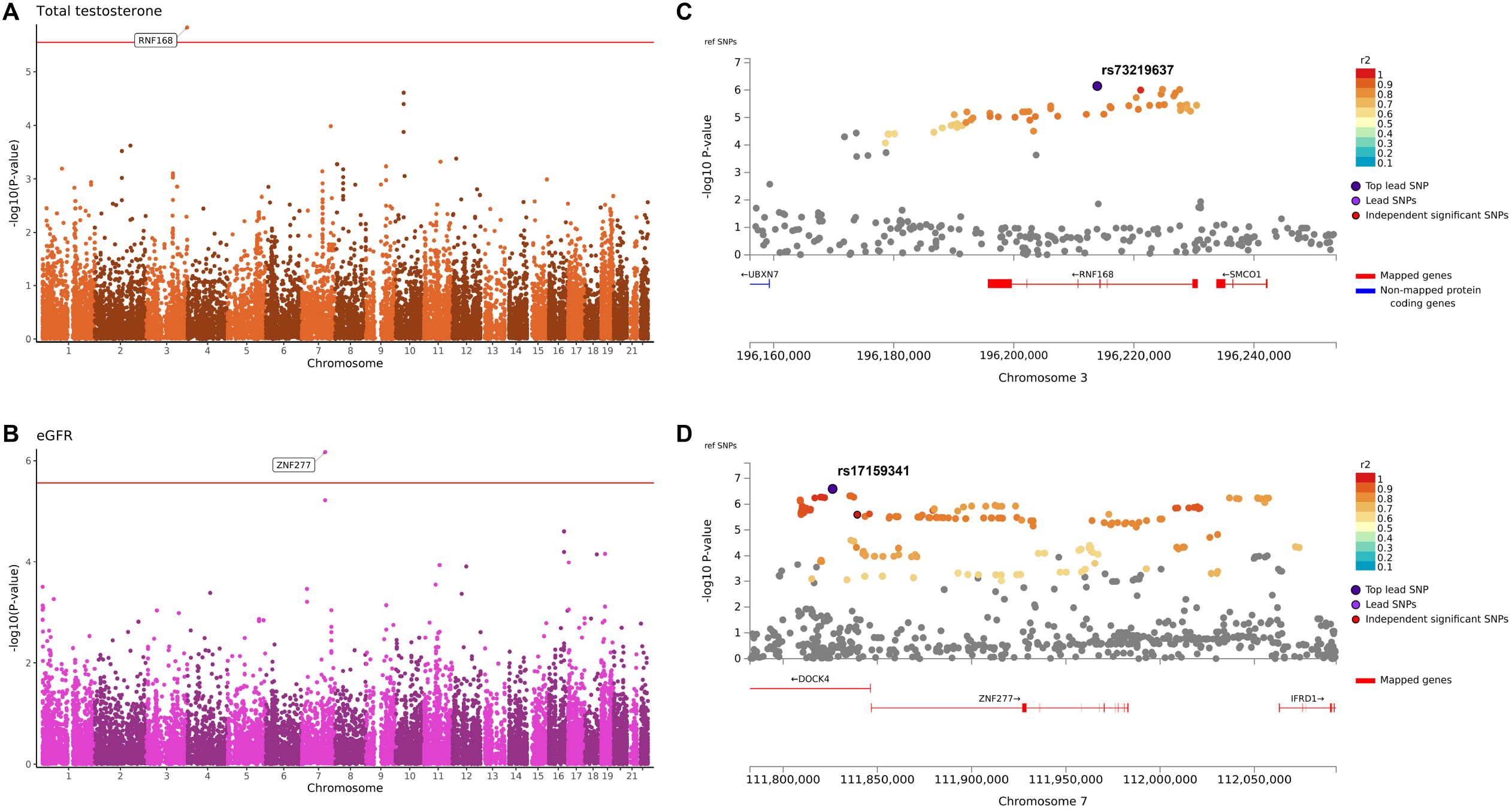
Significant gene-level gene-vegetarianism interactions. Gene-level Manhattan plots for two traits, (A) testosterone and (B) estimated glomerular filtration rate (eGFR), which had gene-vegetarianism interactions that reached significance at a level corrected for the number of genes tested (red line at *P* = 2.75×10^−6^). Local Manhattan plots show the top variant-level interactions at (C) *RNF168* in testosterone and (D) *ZNF277* / *DOCK4* in eGFR. Red genes indicate these genes were positionally mapped to the locus of significant interaction variants. Variants in linkage disequilibrium with the top lead variant are color-coded according to their *r*^*2*^ values.

*RNF168* had the highest expression levels in the testis in GTEx (Supplementary Figure 8B). *RNF168* has previously been associated with testosterone levels at the top variant rs5855544 in UKB GWAS ^29^; however, rs5855544 exceeded our genotype missingness threshold and therefore was not included in this analysis. The top interaction variant at this gene locus was rs73219637 (*P*-int_standard_ = 1.46×10^−7^; *P*-int_adj-BMI_ = 2.31×10^−7^; *P*-marginal_standard_ = 0.435; *P*-marginal_adj-BMI_ *=* 0.338; Figure 4C). The rs73219637 heterozygote (TC) was associated with an increased expression of *RNF168* in the testis (*P* = 4.81×10^−3^), though this did not pass the GTEx multiple testing significance cutoff. The RNF168 protein is involved in the repair of DNA double-strand breaks. Mutation of this gene is associated with Riddle syndrome, symptoms of which include increased radiosensitivity, immunodeficiency, motor control and learning difficulties, facial dysmorphism, and short stature. A mouse model of Riddle syndrome found RNF168 deficiency caused decreased spermatogenesis, and *RNF168* was identified as a tumor suppressing candidate gene in testicular embryonal carcinomas ^48^.

While *ZNF277* contains a number of variants with suggestive interaction *P*-values, the lead variant in this region is rs17159341 (*P*-int_standard_ = 2.58×10^−7^; *P*-int_adj-BMI_ = 8.61×10^−7^; *P*-marginal_standard_ = 0.089; *P*-marginal_adj-BMI_ *=* 0.102; Figure 4D), found in the first intron of *DOCK4. DOCK4* appears to be a more relevant candidate gene than *ZNF277*. Though *DOCK4* has not been directly associated with eGFR in GWAS studies, it has been associated with several traits related to kidney health, such as diastolic blood pressure, type 2 diabetes, dehydroepiandrosterone sulphate measurement (a marker for adrenal disorders) and “Water consumption (glasses per day).” A recent study demonstrated that *in vivo* and *in vitro* DOCK4 expression was found to increase with high-glucose, and that DOCK4 could reverse USP36-induced epithelial-to-mesenchymal transition effect, which is involved in diabetic renal fibrosis and nephropathy ^49^.

## Discussion

In this study we developed a multi-step approach to evaluate the health impacts of vegetarian diets, using both traditional and genetic epidemiological methods, the latter of which has not previously been applied to vegetarianism. Initially, we found several patterns in the UKB participant data that indicated rigorous quality control was necessary to identify “reliable” vegetarians. For example, at each instance of the 24HR, we found that about 10-14% of self-identified vegetarians indicated eating fish, or less often meat, or both, on that same instance of the dietary survey (Figure 1D). Additionally, of 1,229 participants who indicated they “have never eaten meat in [their] lifetime,” (IA, Field 3680), 132 (10.7%) also indicated on the same instance of the dietary questionnaire that they occasionally eat oily fish, with 83 of these participants (6.8%) even indicating they eat oily fish once a week or more (Supplementary figure 10). It seems likely that a portion of vegetarians consider eating fish as be compatible with the diet, despite this contradicting the common usage of the term. We also observed that in the three-year period of 24HR administration between April 2009 and June 2012, many participants either stopped identifying as vegetarian, or began identifying as one (Figure 1C). Duration of vegetarianism adherence has been shown to impact health measures in several studies (i.e., vegetarianism is a “time-dependent exposure”) ^8,19,50-52^. Our results show that self-identification of vegetarianism should be treated with caution in dietary surveys, and single-criterion designations of vegetarianism can produce noise and potentially spurious associations ^51^.

After matching vegetarians to nonvegetarians, we estimated the effects of vegetarianism on 30 serum biomarkers in a traditional model that did not consider genotypic effects. We found vegetarianism had significant effects on 15 of these biomarker traits (Figure 2). The majority of results from our effects estimation can be explained within the context of the restricted dietary cholesterol, increased dietary fiber, and different amino acid profiles found in the plant-based components of vegetarian diets. For example, vegetarianism had significant negative effects on serum levels of total cholesterol, all lipoproteins (LDL, HDL, Lp (a), ApoA, ApoB), and Vitamin D, which is synthesized from cholesterol. Although serum cholesterol is mainly derived from *de novo* synthesis in the liver, our results suggest that intake of animal protein can nonetheless produce significant differences in serum levels of cholesterol and related molecules. These differences may also be due to higher levels of fiber in plant-based diets, which reduces cholesterol and overall inflammation ^53^. Interestingly, vegetarianism had a significant and moderate triglyceride-raising effect. This adds to previous evidence that a vegetarian diet may actually raise triglycerides ^54,55^, though recent large meta-analyses had opposite findings ^1,8^. This positive effect on triglycerides may be explained by lower Vitamin D ^56^, or higher dietary intake of simple carbohydrates ^53^. Conversely, without considering genetic differences, we replicated previous findings ^57^ that vegetarianism does not have significant effects on the cholesterol-derived sterol hormone testosterone (total T, bioavailable-T, and free-T), nor on the testosterone inhibitor SHBG, in both full and sex-stratified models.

Our results did not clearly indicate benefit nor harm of vegetarianism on biomarkers commonly associated with liver function. For example, we found that vegetarianism had a significant negative effect on ALT and GGT, lower levels of which are associated with healthier liver function. Conversely, we observed a significant positive effect on ALP. Increased levels of ALP have been observed in the context of chronic kidney disease (CKD) and Vitamin D deficiency. Several studies have shown a decrease in ALP can be achieved by administering activated Vitamin D compounds ^58^.

Improved kidney biomarkers have been associated with increased plant protein intake ^53^. Creatinine and urea, byproducts of protein metabolism, had a significant negative effect of vegetarianism. This can be explained by lower overall protein intake, amino acid composition, or increased fiber intake in vegetarian diets ^53^. Vegetarianism also had a significant negative effect on urate (AKA “uric acid”), which can cause gout, kidney stones, and kidney injury in high amounts, but is also a serum antioxidant ^59^. Higher consumption of fiber in plant-based diets has been associated with higher eGFR and a lower risk of developing CKD ^53^.

The effect of vegetarianism on serum calcium was small, negative, and marginally significant (*β* = -0.078; *P* = 0.002). Serum calcium is regulated by calcitriol (1,25-dihydroxycholecalciferol), the active form of Vitamin D made in the kidneys. Calcitriol increases serum calcium by increasing the uptake of calcium from the intestines, and may also increase calcium excretion via decreased parathyroid synthesis ^60^. Calcium deficiency is a known risk in vegetarian diets, though it can be improved by increased dairy consumption, and is also indirectly dependent on intake of sodium, caffeine, and total protein ^61^.

We did not find a “vegetarianism gene,” nor any variant that was significantly associated with vegetarianism. This null finding is similar to a recent GWAS in a Japanese cohort ^25^. Variants in *HLA-DPB1*, the most significant hit in the gene-based test, have been previously associated with cognitive empathy ^62^, which could potentially be involved with one’s decision to become vegetarian. This connection, while interesting, is speculative, and more evidence is necessary. We also found that for all three significant interaction loci at rs72952628, *RNF168* and *DOCK4*, there were no significant genetic associations in our GWAS of vegetarianism, nor significant genetic marginal effects on their respective biomarker traits. This strengthens the likelihood of interaction effects driving the signals at these loci.

We performed GWIS across 30 biomarkers, and identified significant gene-vegetarianism interactions in three traits: a variant-level interaction in calcium, and gene-level interactions in testosterone and eGFR. These represent the first gene-vegetarianism interactions identified to-date. Only one variant-level interaction at rs72952628 reached GWS (*P*-int = 4.47×10^−8^). This SNP was found to be significantly associated with expression changes in *MMAA*, a protein in the B_12_ metabolism pathway. B_12_ deficiency is the highest nutritional concern in vegetarians, and dietary intake plays a primary role in B_12_ availability ^1,45^. And, though we did not directly query B_12_ levels, its metabolism pathway was implicated in our results. We have suggested *CD320* as a calcium-dependent protein in this pathway; *CD320* serves as the cellular gateway for transcobalamin-bound B_12_ to the cell ^47^. Similarly, we have proposed *RNF168* and *DOCK4* as the most likely candidate genes based on gene expression and experimental evidence related to testosterone and eGFR, respectively. More experimental evidence is needed to validate these proposals, as there may be less direct mechanisms involved in these interactions.

Two of three traits with significant gene-vegetarianism interactions, eGFR and calcium, are closely related to kidney function. This is likely due to the major differences in levels, composition, and bioavailability of proteins and minerals, plus the higher overall alkalinity found in vegetarian diets, which directly impact kidney function ^53^. Meanwhile Vitamin D, whose activated form regulates serum calcium, as well as testosterone, which had significant gene-vegetarianism interactions, are steroids synthesized from cholesterol. Interestingly, none of the three traits found to have gene-vegetarianism interactions showed significant effects of vegetarianism (at *P* < 0.0017) in the traditional, non-genetic epidemiological analysis. This emphasizes the importance of genetic interactions in modeling the phenotypic effects of an exposure.

Our study has several limitations. First, we performed a one-stage analysis (discovery only) without replication. The UKB is among the first datasets which both contain dietary data and are sufficiently powered for a GWIS. The benefit in our study of being able to utilize multiple dietary surveys and criteria in defining vegetarians from UKB, also caused us to be unable to produce an equally rigorous set of vegetarians for replication. Nonetheless, we consider these one-stage results valuable, for several reasons. We have clearly demonstrated the noise in a single-criterion definition of vegetarianism in UKB ^51^, which is relevant in interpreting other vegetarianism studies. Next, the use of algorithmic matching in our traditional epidemiological analysis, which simulates the experimental design of a large-scale randomized control trial, established a greater balance between vegetarians and nonvegetarians than has been achieved in previous analyses ^1,8,9^. Finally, despite being unreplicated, our GWIS results are valuable in the exploratory context of identifying gene-vegetarianism interactions as an important and novel nutrigenetic phenomenon.

Another limitation of this study is that although we found significant interactions that passed GWS thresholds, none of these met a threshold further corrected for thirty traits. In this multi-trait GWIS, the multiple testing burden was high, on top of an already strict GWS threshold. GWIS have sample size requirements which require approximately four times more participants to achieve the same power as in a GWAS with comparable effect sizes ^63,64^. We suspect that future studies with larger sample sizes will produce a higher number of significant loci. This is supported by several biologically plausible interactions, for example, *BRINP3* in Vitamin D (*P* = 3.88×10^−6^), and *INTU* in SHBG (*P* = 3.93×10^−6^) which nearly reached gene-level significance.

In contrast with increasingly common recommendations that vegetarianism is universally beneficial for all people ^4,6,7,65^, we found several significant biomarker signals corresponding to potentially worse health in vegetarians. Our traditional epidemiological analysis showed a triglyceride-raising effect of vegetarianism; raised triglycerides are a symptom of metabolic syndrome and a risk factor for heart disease and stroke. Lower Vitamin D and higher ALP were also observed, both of which have been associated with negative health outcomes as described above. Two traits, urate and testosterone, have been associated with depression ^59,66^; depression has been repeatedly associated with vegetarianism in observational studies ^67^. These findings are most relevant for those who are in the same age range (40 to 70 years old) as our study cohort; vegetarian and vegan diets for children ^68^ and pregnant women ^69^ are associated with more serious risks of malnutrition.

The emerging paradigms of precision medicine and precision nutrition suggest that genetic makeup should help inform optimal disease treatment strategies. We identified three novel gene-vegetarianism interactions in this study and used available functional analyses to put these interactions into plausible biological context. But as in any genome-wide study, these statistically significant interactions must be externally replicated and verified experimentally. We hope that future researchers can use our results, analysis protocol, and open computational pipeline in future studies to conduct replications and meta-analyses, and to inform clinical trials.

## Data Availability

Full and annotated code used in this analysis, gene-level summary statistics, and interactive Manhattan plots are publicly available at https://michaelofrancis.github.io/VegetarianGDI/.
Summary statistics for GWAS and GWIS at GWAS Catalog (https://www.ebi.ac.uk/gwas/). The corresponding accession numbers can be found in Supplementary table S5.

## Acknowledgements

Research reported in this publication was supported by the National Institute of General Medical Sciences of the National Institute of Health under award numbers T32GM007103 (MF) and R35GM143060 (KY). The content is solely the responsibility of the authors and does not necessarily represent the official views of the National Institutes of Health.

Special thanks to the Georgia Advanced Computing Resource Center (GACRC) at the University of Georgia for supporting our data analyses.

## Competing interests

The authors declare no competing interests.

## Author contributions

MF designed and performed the analysis. KY supervised the project.

## Data sharing

Full and annotated code used in this analysis, gene-level summary statistics, and interactive Manhattan plots are publicly available at https://michaelofrancis.github.io/VegetarianGDI/. Summary statistics for GWAS and GWIS at GWAS Catalog (https://www.ebi.ac.uk/gwas/). The corresponding accession numbers can be found in Supplementary Table S5.

## Supplementary figures

**S1. Boxplots of unadjusted trait values**. Comparing raw values of vegetarians (as defined by Table 1) and nonvegetarians across 30 biomarker traits. Boxplots show first decile, first quartile, median, third quartile, and last decile. Dot and label refer to mean. Units of each biomarker (“value”) can be found in table S1. Matched cohort details are found in table S2 and S3. (A) Full cohort. (B) Stratified by sex.

**S2. Love plot of covariates before and after matching**. Plot shows the absolute standardized mean difference of model covariates in nonvegetarians before and after matching with vegetarians for effects estimation. After matching, the ASMD in all model covariates were < 0.05 standardized units. BMI=body mass index; AlcoholFreq = frequency of alcohol usage (<3 drinks/week or ≥ 3 drinks/week); zTownsend = standardized Townsend deprivation index; PCA = genetic principal component; distance = matching distance between participants was calculated by general linearized model.

**S3. Sex-stratified forest plot**. Effects estimation for vegetarianism in BMI-adjusted model. Participants stratified by male or female. Error bars indicate 95% confidence interval. Bonferroni-corrected significance threshold at *P* = 0.0017. Full data is shown in **S3 table**.

**S4. Correlation plot comparing *P*-values of BMI-adjusted model**. Vegetarianism GWAS −log_10_(*P*) between BMI-adjusted versus standard (without BMI) models were compared. Each point represents one variant. Spearman’s Rho (*R*) and correlation *P*-value shown. Correlation coefficients for interaction analysis are found in **S4 table**.

**S5. Vegetarianism genome-wide association Manhattan plots**. Manhattan plots and QQ plots showing the −log_10_(*P*) of genetic effects with vegetarianism as a binary trait outcome. Genomic control (λ) for each model is shown in the QQ plots. Plots correspond to (A) Variant-level GWAS, (B) variant-level (BMI adjusted) GWAS, (C) gene-level GWAS where *P*-values were aggregated by MAGMA and (D) gene-level GWAS (BMI-adjusted). Top variants in a 60 Mb window that exceeded the genome-wide suggestive threshold (*P* = 1×10^−5^; blue line) are annotated. Top genes (*P* < 1×10^−4^) in a 5 Mb window were annotated. No variants or genes for vegetarianism as a trait were significant.

**S6. Variant-level gene-vegetarianism interaction Manhattan plots**. Manhattan plots and QQ plots showing the variant-level −log_10_(*P*) of genome-wide gene-vegetarianism interaction effects in thirty serum biomarker traits. The blue line corresponds to the genome-wide suggestive threshold (*P* < 1×10^−5^). In the standard interaction model (A), one trait, calcium, had a significant variant above the genome-wide significance threshold (*P* < 5×10^−8^; red line). No variants were significant in the BMI-adjusted model (B).

**S7. eQTLs for MMAA and rs72952628**. Violin plots showing expression quantitative trait loci (eQTLs) in four tissues which were significant at the GTEx multiple testing threshold (adipose: subcutaneous, colon: sigmoid, muscle: skeletal, and cells: cultured fibroblasts) plus liver tissue, which nearly reached significance. In all five of these tissues, the heterozygote (CT) shows higher median normalized expression.

**S8. Bulk tissue gene expression for interaction genes**. Candidate genes for significant interactions with vegetarianism in either the variant-level or gene-level analyses. Transcripts per million (TPM) shown in tissues ranked from low to high for the genes (A) *MMAA*, (B) *RNF168*, and (C) *DOCK5*.

**S9. Gene-level gene-vegetarianism interaction Manhattan plots**. Manhattan plots and QQ plots showing the gene-level −log_10_(*P*) of genome-wide gene-vegetarianism interaction effects in thirty serum biomarker traits. The red line corresponds to the genome-wide significance threshold(*P* < 2.75×10^−6^; red line). In the standard interaction model (A) two traits, estimated glomerular filtration rate (eGFR) and testosterone, had a significant gene. (B) Testosterone had one significant gene in the BMI-adjusted model.

**S10. Fish eating frequency of those who have “never eaten meat” in their lifetime**. Bar plot shows non-oily-fish- and oily-fish-eating frequency, reported at the initial assessment, for those who reported on that same dietary survey that they had “never eaten meat in [their] lifetime” (N=1,230).

## Supplementary tables

**S1. Participant characteristics**. Categorical covariate (top), continuous covariate (middle) and phenotype data for 155,375 European UK Biobank participants used in analyses. Continuous variables are represented as: mean (standard deviation). Values are shown as full cohort and stratified by vegetarianism as defined in Table 1.

**S2. Matching summary**. Top: matchit function call used for 1:4 matching of vegetarian and nonvegetarian participants for use in effects estimation analysis. Middle: Summary of balance for all data and for matched data shows the results of matching on relevant lifestyle factors and genetic principal components one through five. Bottom: Sample sizes in control (nonvegetarian) and treated (vegetarian) samples before and after matching. Std. Mean Diff. = standardized mean difference; Var. Ratio = variance ratio; eCDF Mean = empirical cumulative density functions to assess imbalance across entire covariate distribution; eCDF Max = maximum eCDF difference, also known as the Kolmogorov-Smirnov statistic.

**S3. Vegetarianism effects on biomarkers**. Effects of vegetarianism across 30 traits in full and sex-stratified matched groups. Left = BMI-adjusted models, right = models without BMI. BetaVeg = the effect of vegetarianism; SE = standard error; BMI = body mass index; M = male only; F = female only.

**S4. Summarize GWAS/GWIS**. Top: Most significant hits for GWAS of vegetarianism as a trait in variant-level and gene-level analysis. Most significant interaction hits across 30 traits in variant-level and gene-level analysis. All traits analyzed in standard and BMI-adjusted models. Start and stop coordinates of genes represent the +2 Kbp upstream and -1 Kbp downstream window of variant P-value aggregation. GC λ = genomic control; A0 = non effect allele; A1 = effect allele; A1Freq = frequency of effect allele; P interaction = *P*-value of 1df interaction test for variant calculated with robust standard errors; NSNPS = number of SNPs annotated to the top gene; NPARAM = number of relevant parameters used in model; P interaction (MULTI) = gene *P*-value for best fit of “mean” and “top” models.

**S5. GWAS catalog accessions**. GWAS Catalog accession codes for all variant-level GWAS and GWIS summary statistics generated in this study.

## Notes

### Competing Interest Statement

The authors have declared no competing interest.

### Author Declarations

This project using existing UKB data was approved by the Institutional Review Board (IRB) at the University of Georgia.

### Summary of Updates

Textual and formatting updates; no changes to data.

