## Supplementary Figures S1-10 for "Gene-vegetarianism interactions in calcium, testosterone, and eGFR identified in genome-wide analysis across 30 biomarkers": Supplementary Figs 1-10.pdf

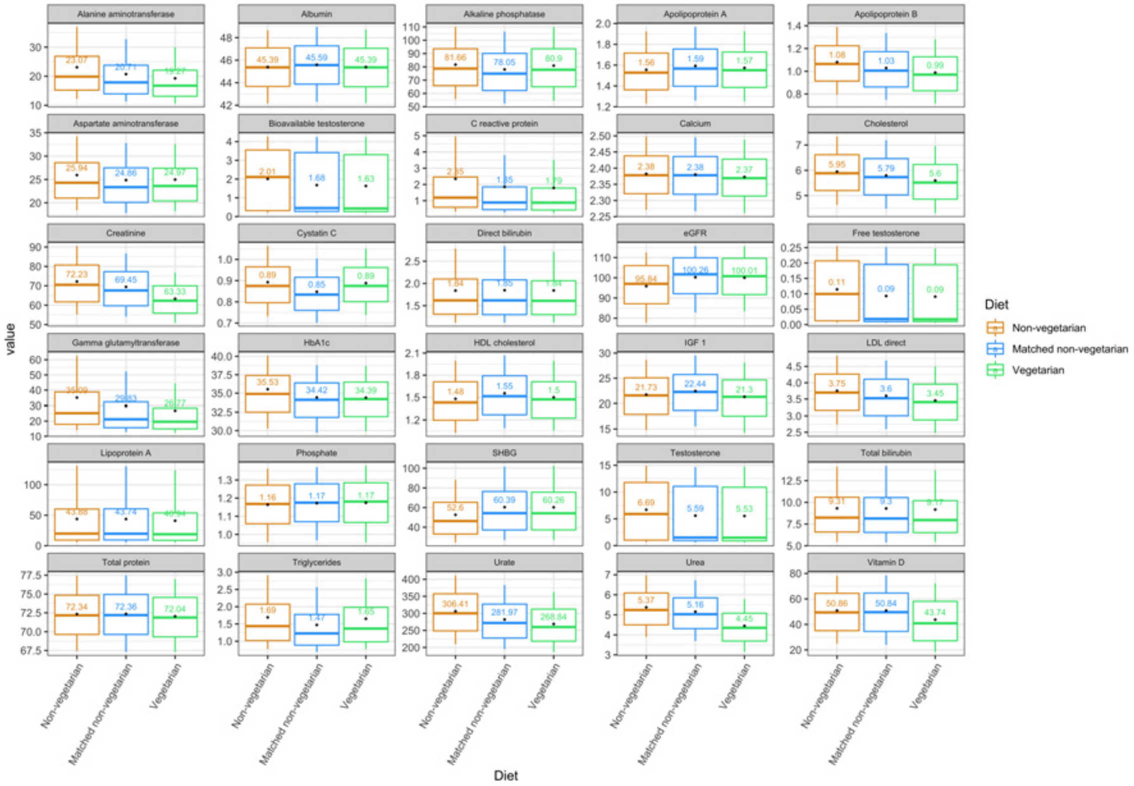
Alanine aminotransferase
Albumin
Alkaline phosphatase
Apolipoprotein A
Apolipoprotein B
Aspartate aminotransferase
Bioavailable testosterone
C reactive protein
Calcium
Cholesterol
Creatinine
Cystatin C
Direct bilirubin
eGFR
Free testosterone
Gamma glutamyltransferase
HbA1c
HDL cholesterol
IGF 1
LDL direct
Lipoprotein A
Phosphate
SHBG
Testosterone
Total bilirubin
Total protein
Triglycerides
Urate
Urea
Vitamin D
value
Diet
Non-vegetarian
Matched non-vegetarian
Vegetarian

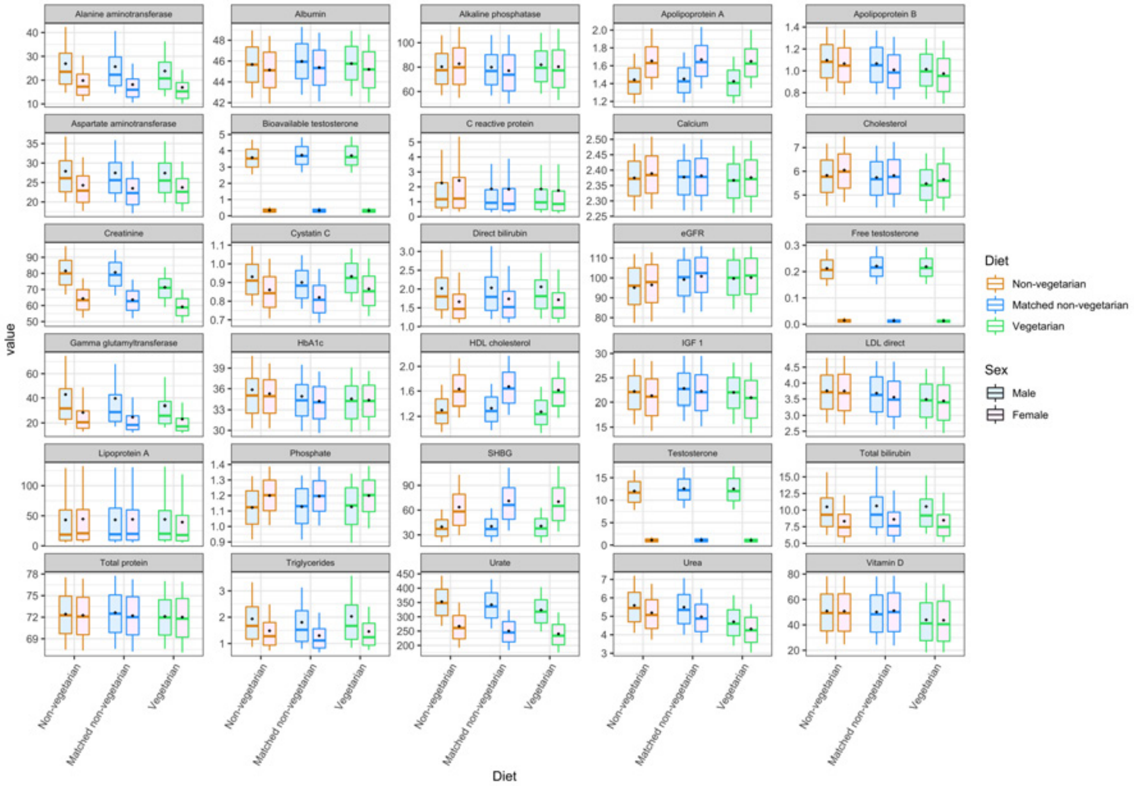
Alanine aminotransferase
Albumin
Alkaline phosphatase
Apolipoprotein A
Apolipoprotein B
Aspartate aminotransferase
Bioavailable testosterone
C reactive protein
Calcium
Cholesterol
Creatinine
Cystatin C
Direct bilirubin
eGFR
Free testosterone
Gamma glutamyltransferase
HbA1c
HDL cholesterol
IGF 1
LDL direct
Lipoprotein A
Phosphate
SHBG
Testosterone
Total bilirubin
Total protein
Triglycerides
Urate
Urea
Vitamin D
value
Diet
Non-vegetarian
Matched non-vegetarian
Vegetarian
Sex
Male
Female

**S2**

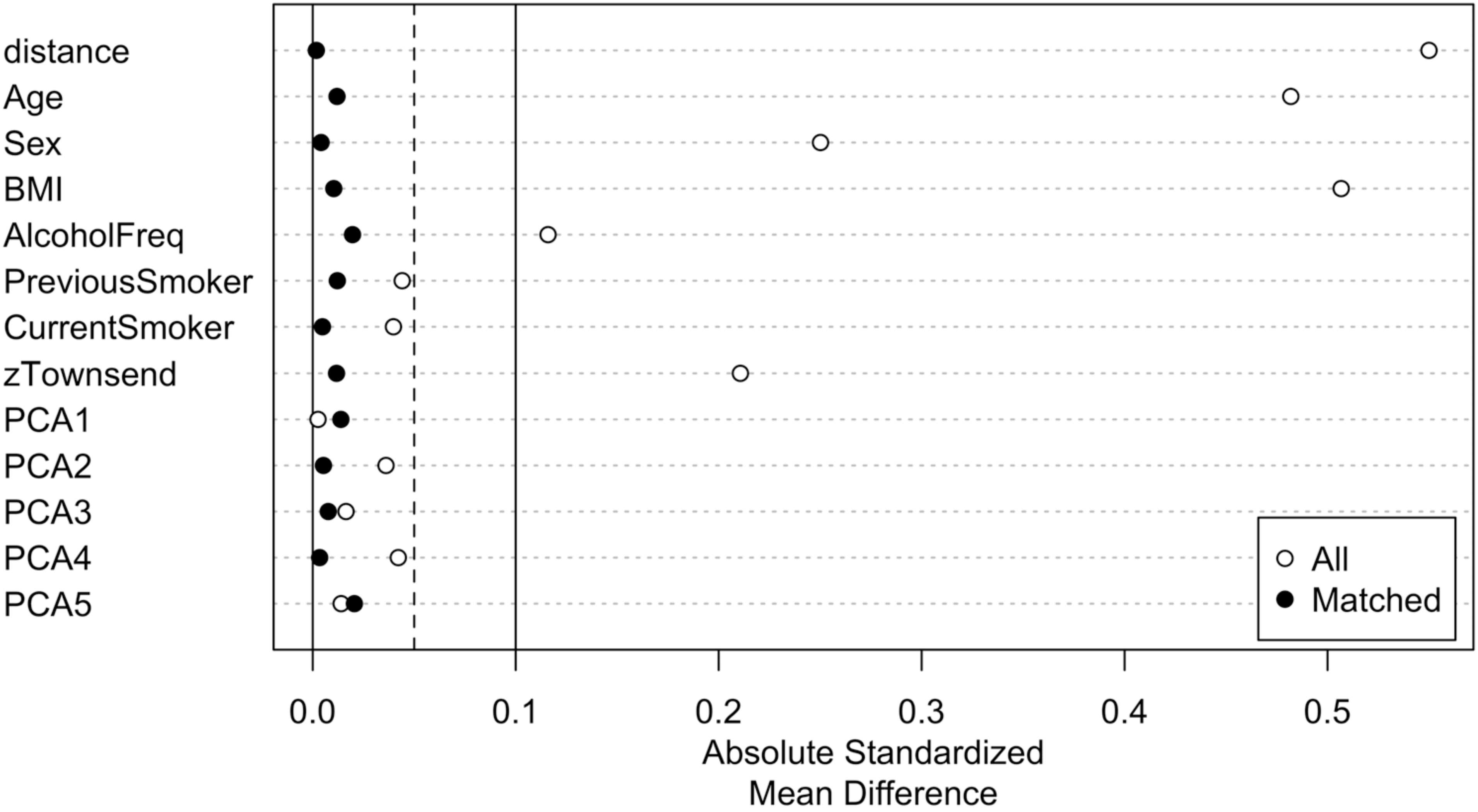
distance
Age
Sex
BMI
AlcoholFreq
PreviousSmoker
CurrentSmoker
zTownsend
PCA1
PCA2
PCA3
PCA4
PCA5
All
Matched
0.0
0.1
0.2
0.3
0.4
0.5
Absolute Standardized
Mean Difference

## S3

| Category | Trait | P (male) | P (female) |
|---|---|---|---|
| Blood lipids | Apolipoprotein A | 3.6e-04 | 0.046 |
| | Apolipoprotein B | 4.6e-07 | 5.0e-10 |
| | HDL cholesterol | 2.5e-09 | 6.8e-08 |
| | LDL direct | 1.5e-08 | 6.8e-09 |
| | Lipoprotein A | 0.473 | 4.8e-04 |
| | Cholesterol | 4.7e-09 | 3.2e-10 |
| | Triglycerides | 8.0e-09 | 5.0e-20 |
| Hormones | IGF 1 | 3.3e-05 | 5.3e-17 |
| | Vitamin D | 9.5e-16 | 1.5e-35 |
| | Bioavailable testosterone | 0.275 | 0.042 |
| | Free testosterone | 0.311 | 0.293 |
| | Testosterone | 0.280 | 0.035 |
| | SHBG | 0.264 | 0.389 |
| Liver markers | Alanine aminotransferase | 1.3e-04 | 2.1e-08 |
| | Albumin | 0.040 | 0.032 |
| | Alkaline phosphatase | 0.005 | 5.7e-08 |
| | Aspartate aminotransferase | 0.997 | 0.030 |
| | C reactive protein | 0.583 | 0.002 |
| | Gamma glutamyltransferase | 3.5e-06 | 5.3e-08 |
| | Direct bilirubin | 0.768 | 0.611 |
| | Total bilirubin | 0.715 | 0.265 |
| | Total protein | 0.005 | 0.154 |
| Kidney markers | Creatinine | 1.0e-86 | 2.8e-61 |
| | Cystatin C | 1.8e-19 | 5.0e-49 |
| | eGFR | 0.950 | 0.094 |
| | HbA1c | 0.307 | 0.069 |
| | Urate | 8.3e-10 | 1.7e-12 |
| | Urea | 3.6e-58 | 9.2e-99 |
| Minerals | Calcium | 0.007 | 0.057 |
| | Phosphate | 0.846 | 0.472 |

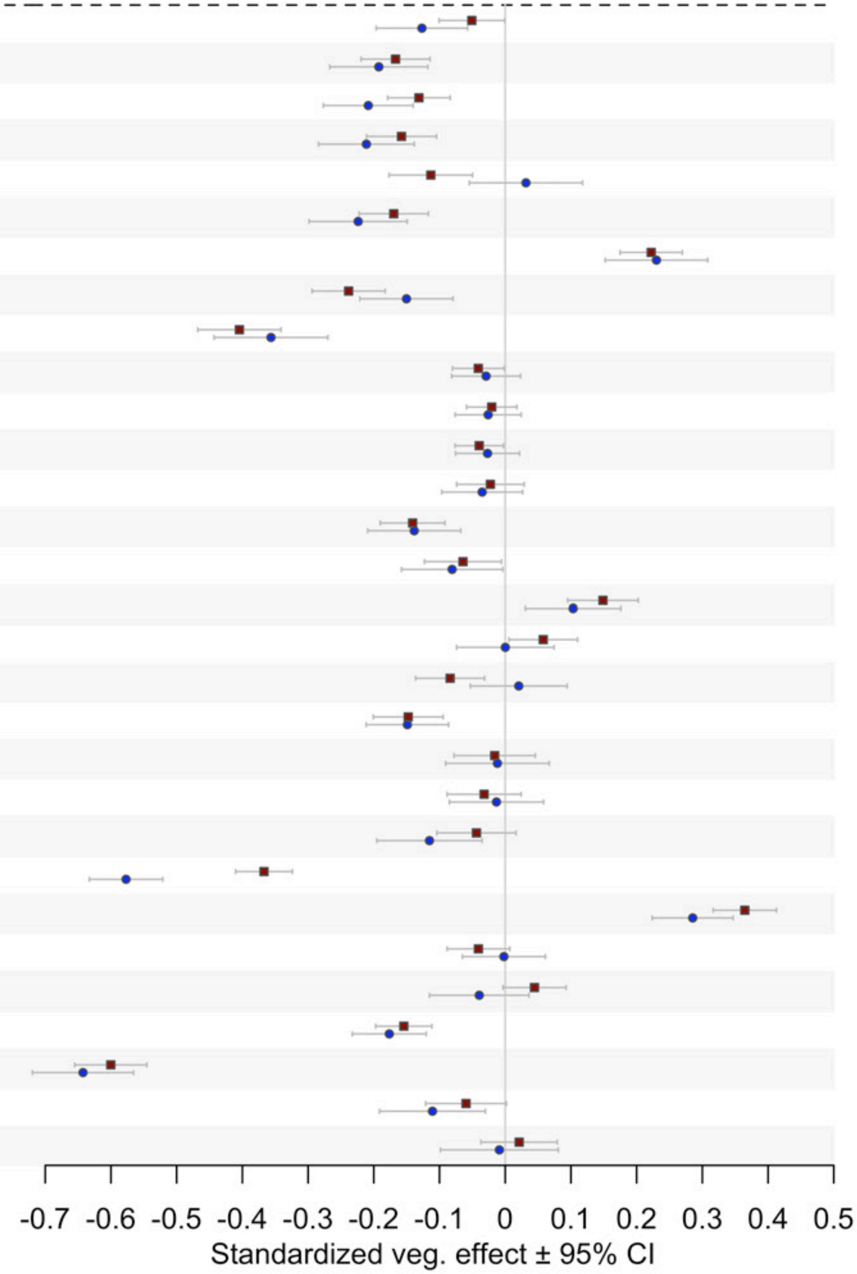

**S4**

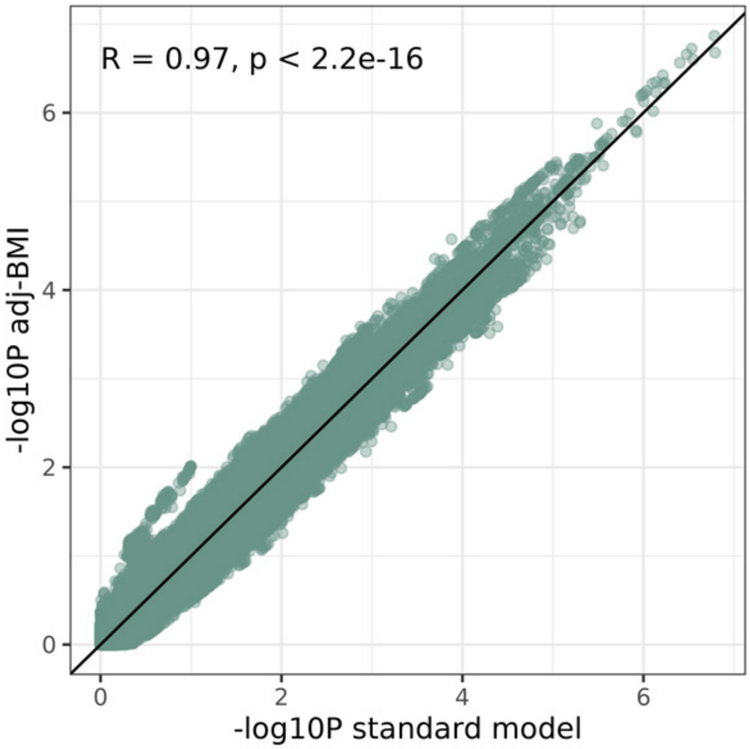

**S5**

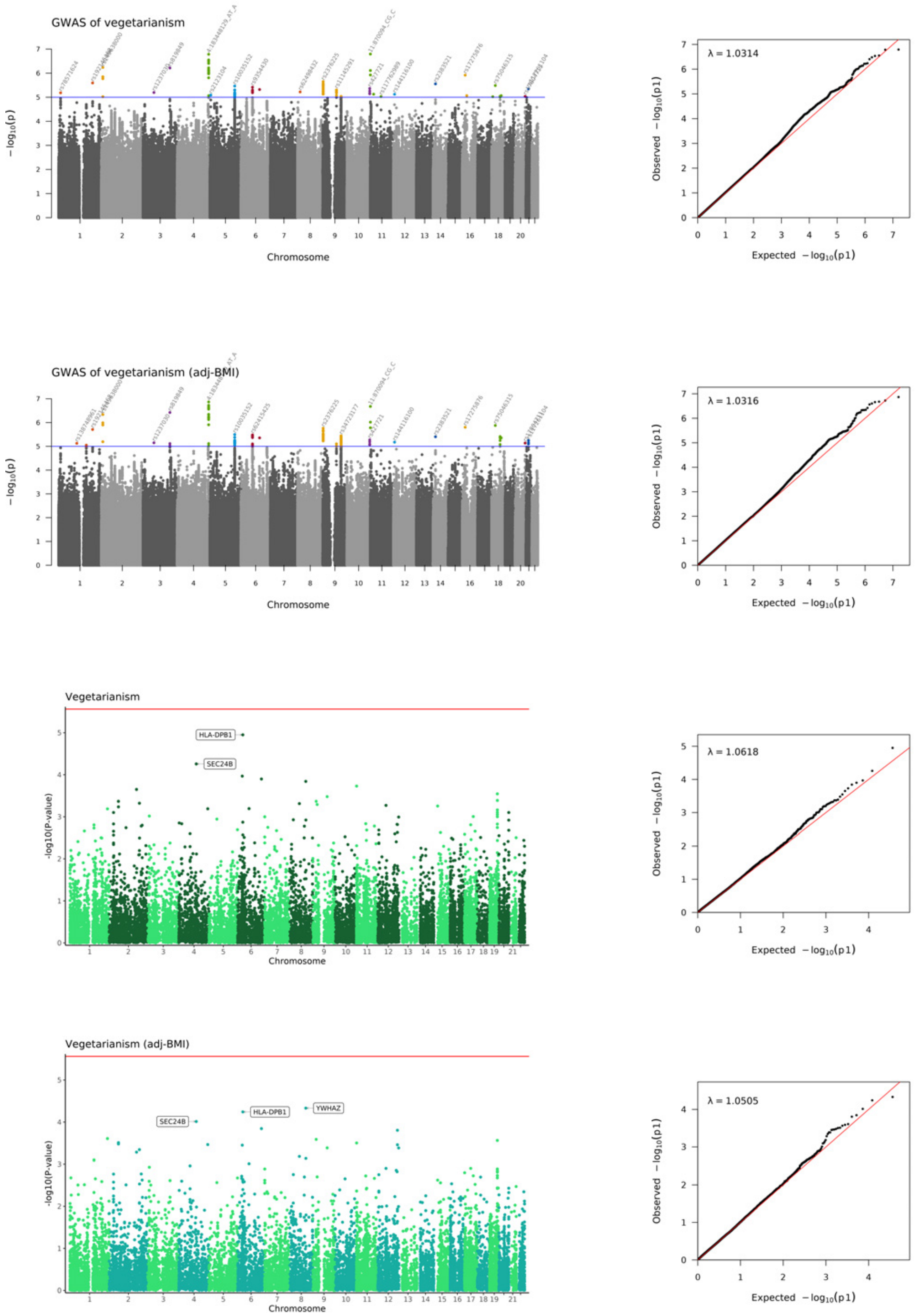
GWAS of vegetarianism
− log10(p)
Chromosome
λ = 1.0314
Observed − log10(p1)
Expected − log10(p1)
GWAS of vegetarianism (adj-BMI)
− log10(p)
Chromosome
λ = 1.0316
Observed − log10(p1)
Expected − log10(p1)
Vegetarianism
HLA-DPB1
SEC24B
-log10(P-value)
Chromosome
λ = 1.0618
Observed − log10(p1)
Expected − log10(p1)
Vegetarianism (adj-BMI)
SEC24B
HLA-DPB1
YWHAZ
-log10(P-value)
Chromosome
λ = 1.0505
Observed − log10(p1)
Expected − log10(p1)

# S6

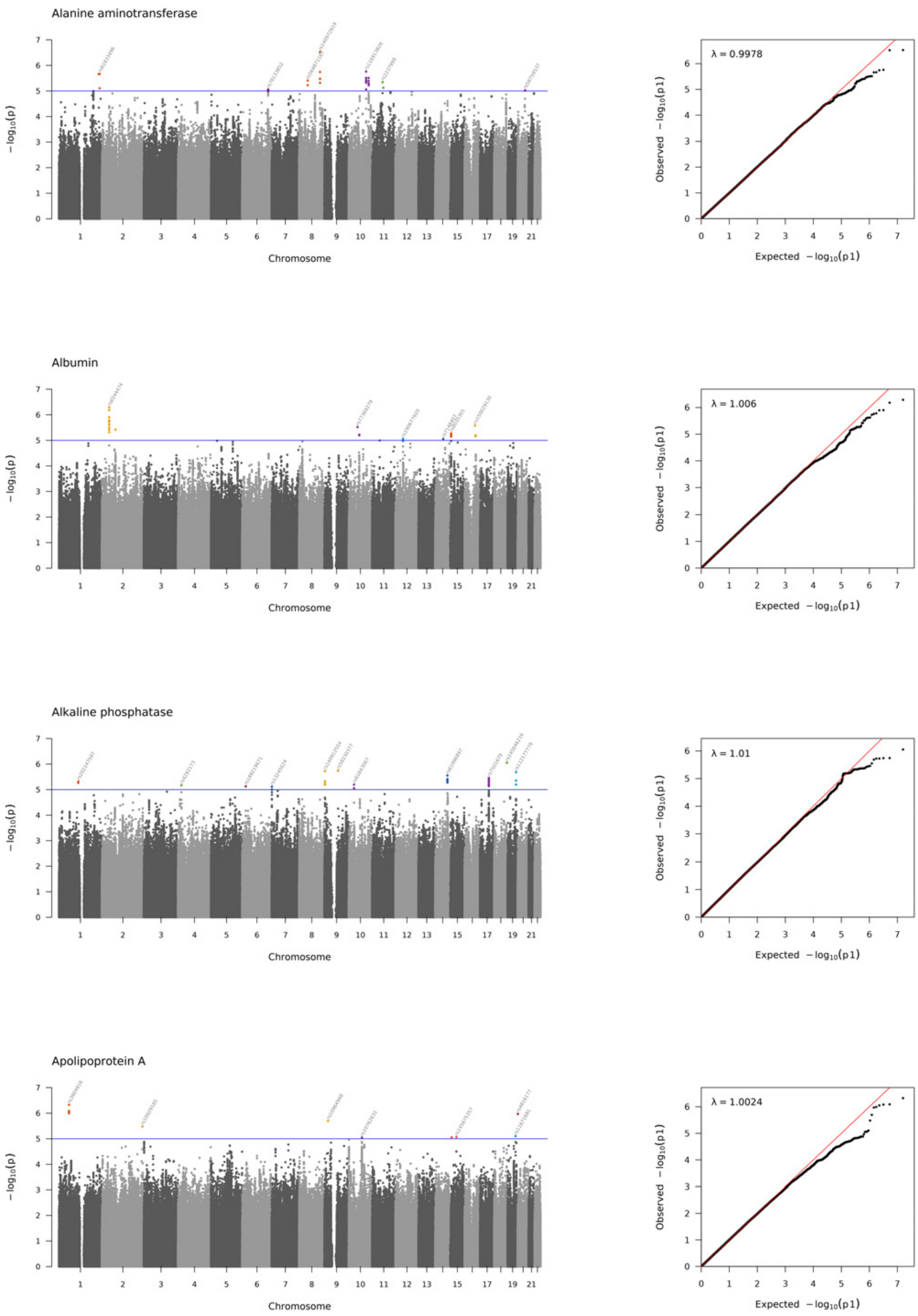
Alanine aminotransferase
λ = 0.9978
Albumin
λ = 1.006
Alkaline phosphatase
λ = 1.01
Apolipoprotein A
λ = 1.0024
− log10(p)
Chromosome
Observed − log10(p1)
Expected − log10(p1)

Apolipoprotein B

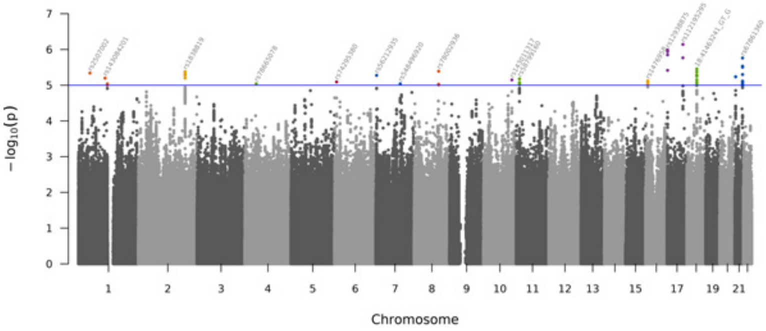


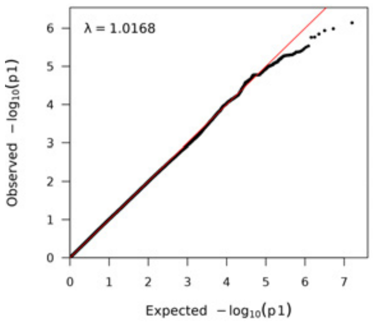


Aspartate aminotransferase

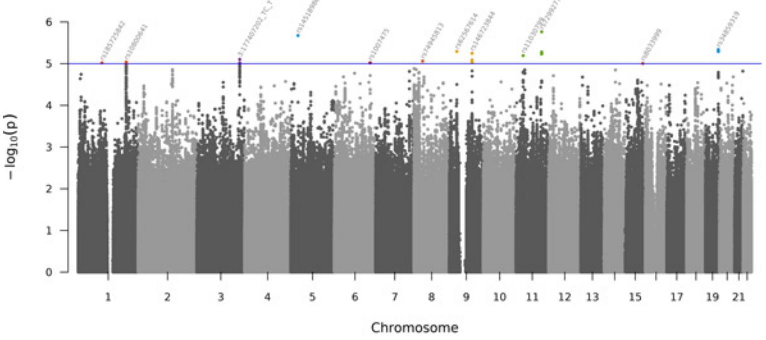


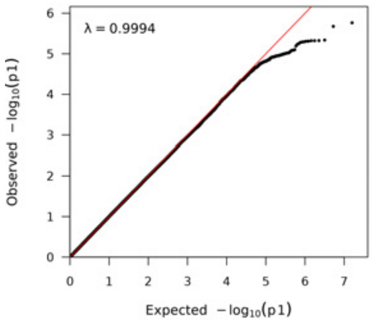


Bioavailable testosterone

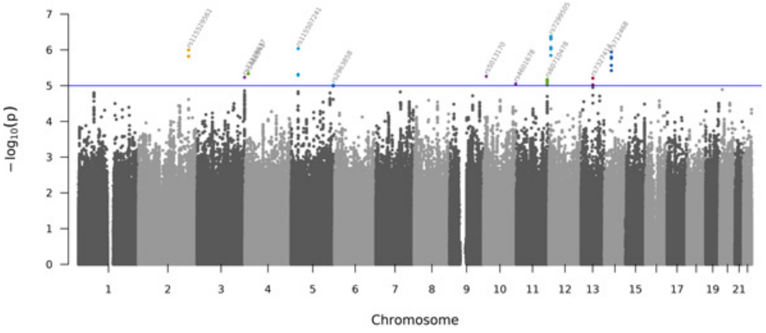


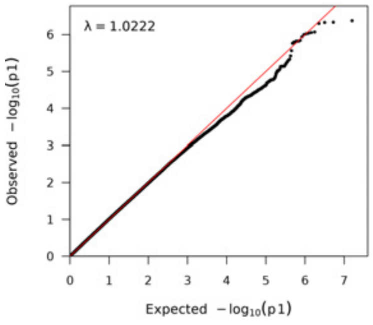


C-reactive protein

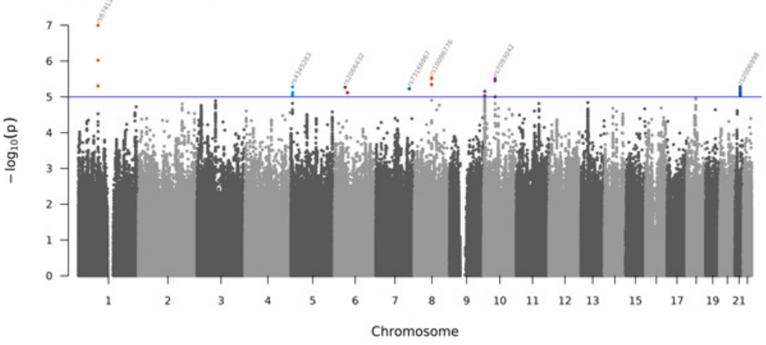


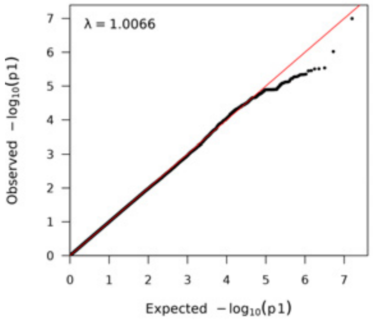

Calcium

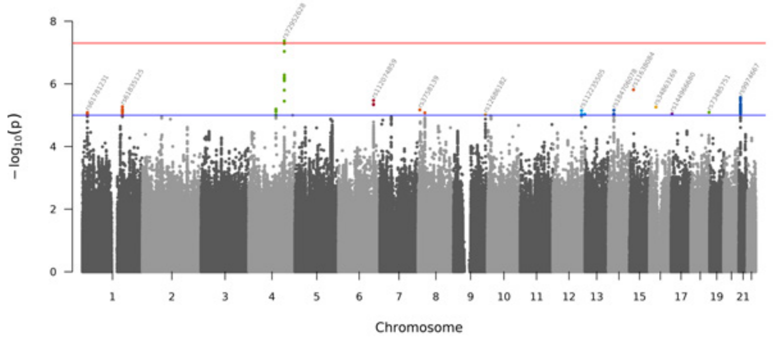


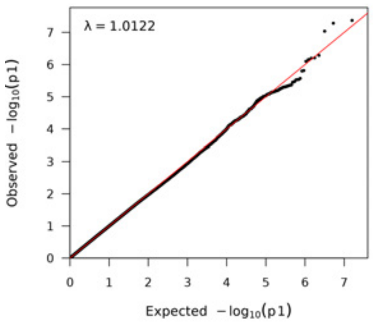


Cholesterol

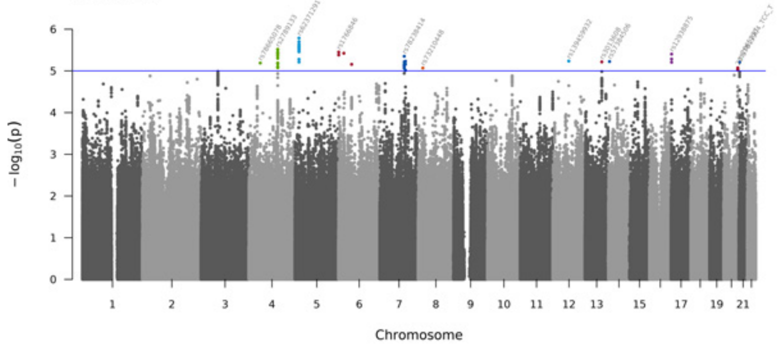


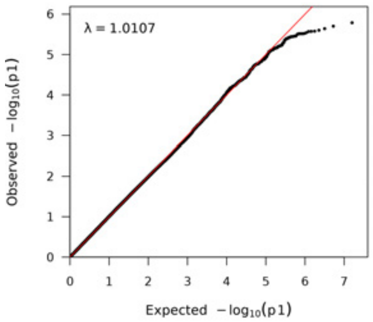


Creatinine

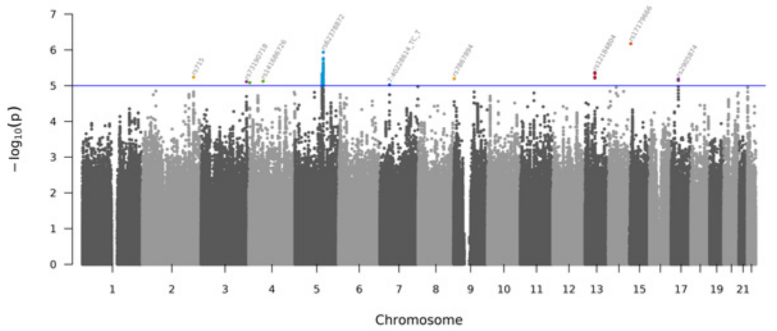


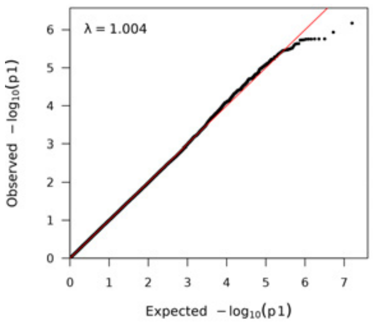


Cystatin C

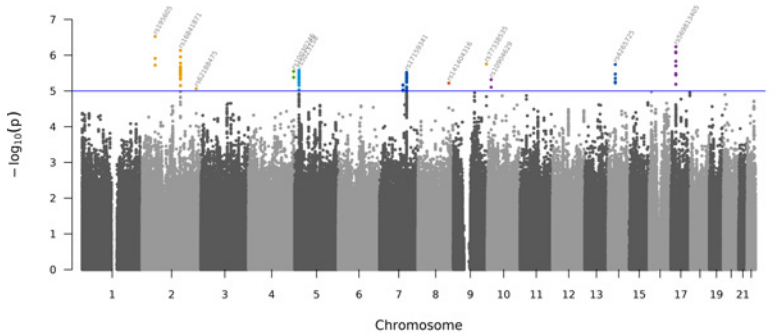


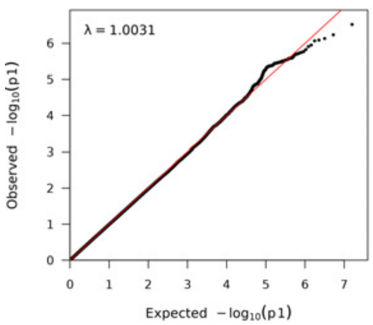

Direct bilirubin

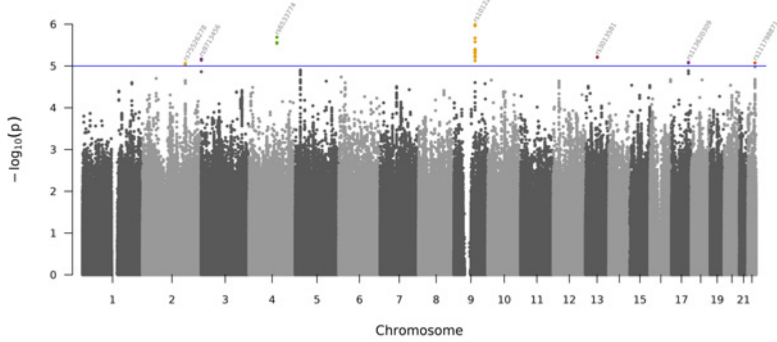


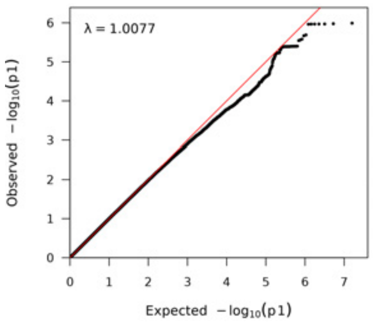


eGFR

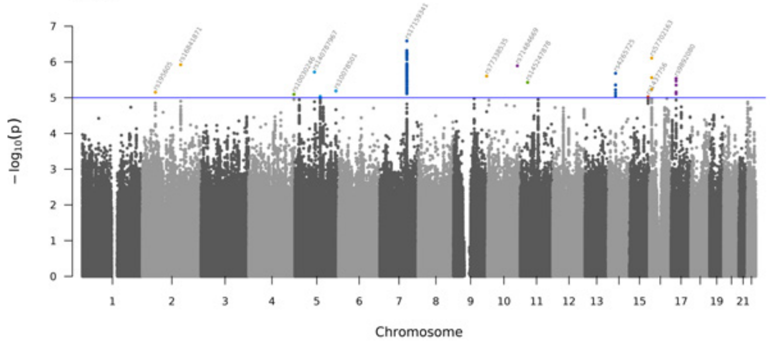


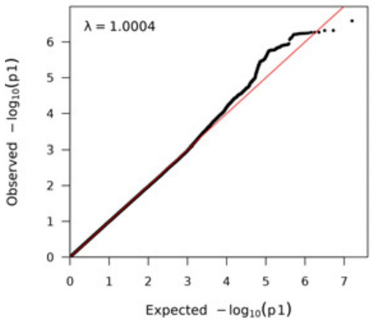


Free testosterone

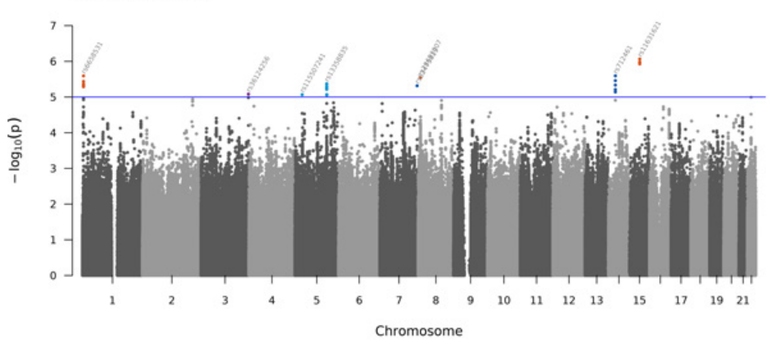


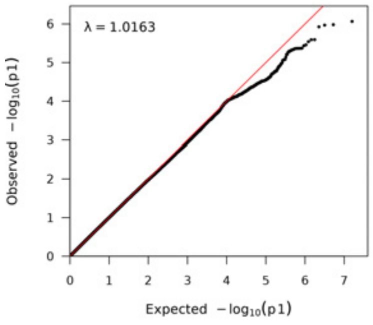


Gamma glutamyltransferase

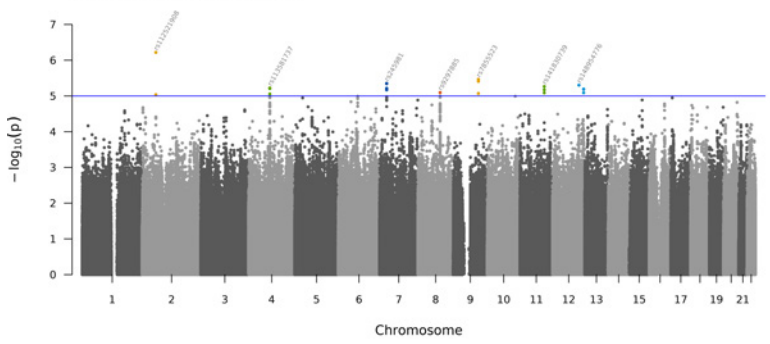


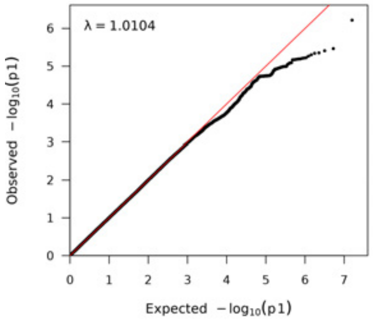

HbA1c

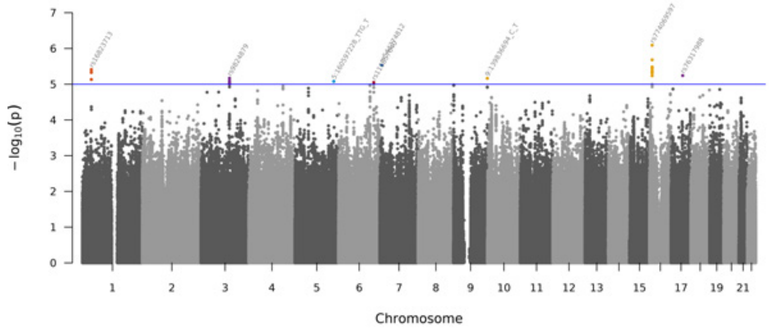


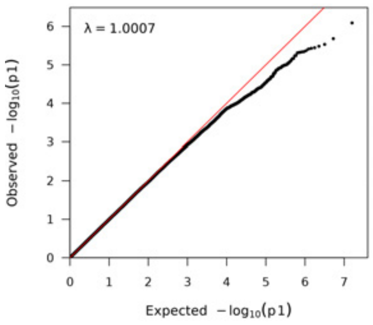


HDL cholesterol

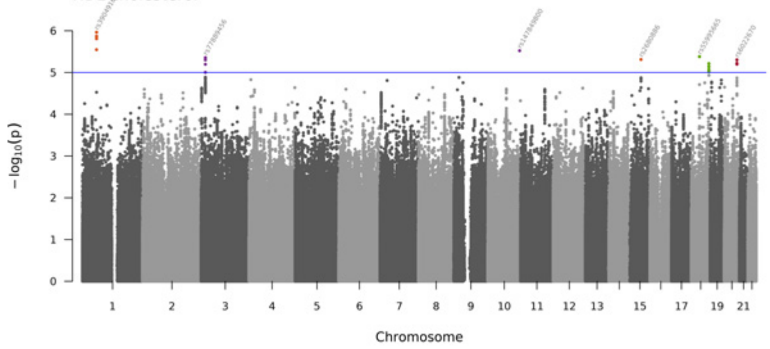


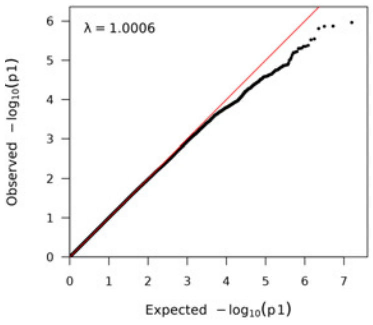


IGF 1

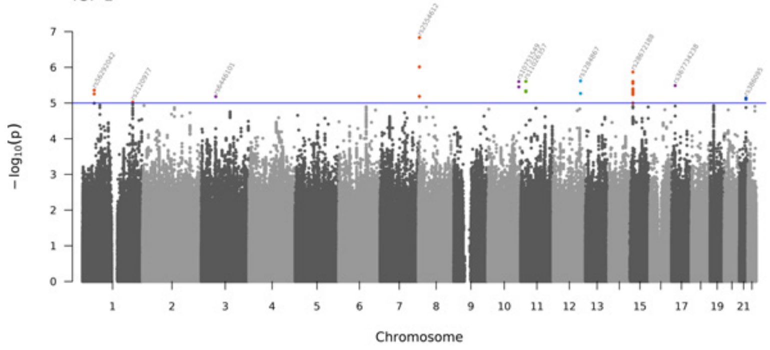


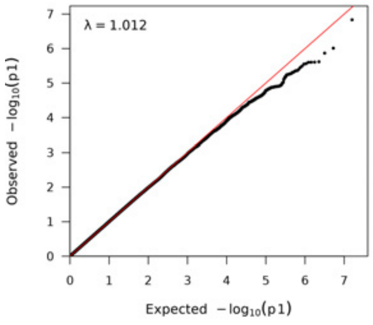


LDL direct

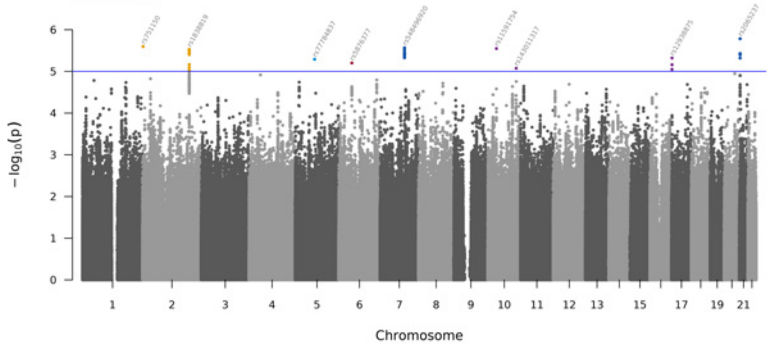


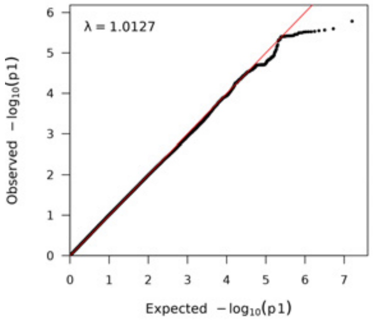

Lipoprotein A

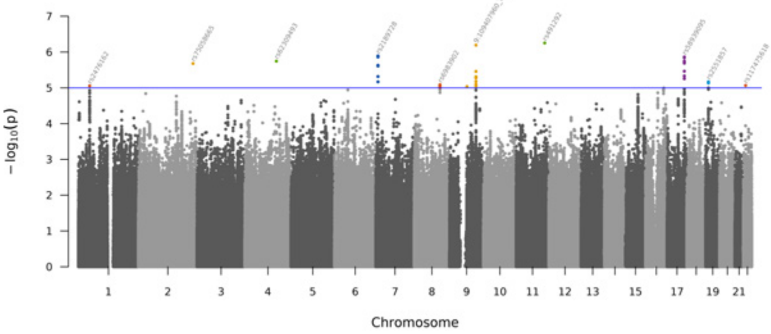


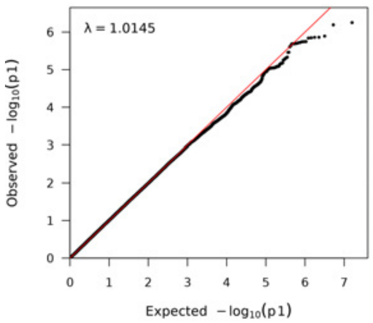


Phosphate

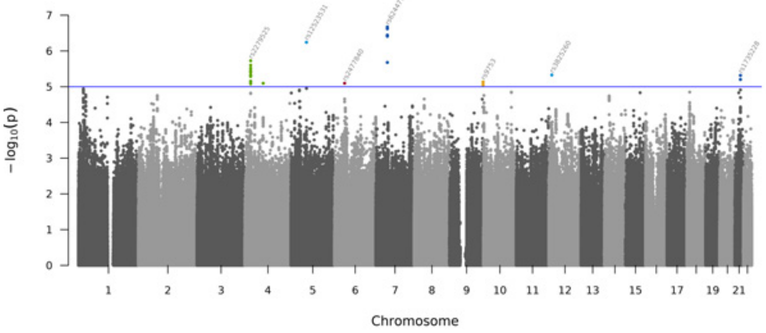


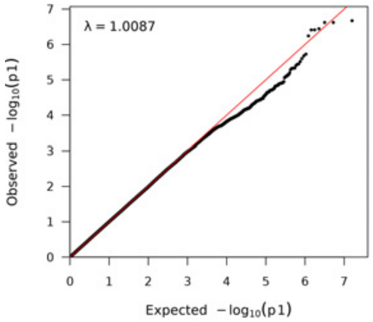


SHBG

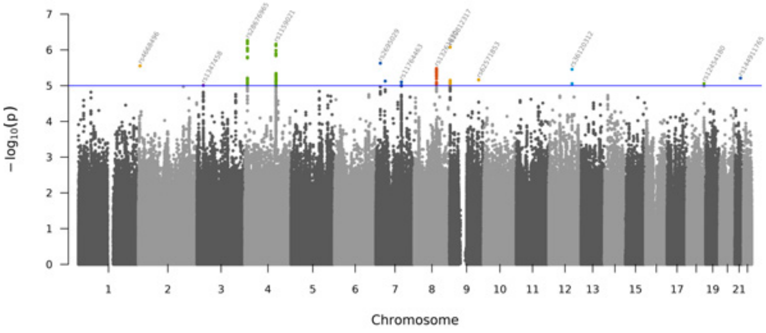


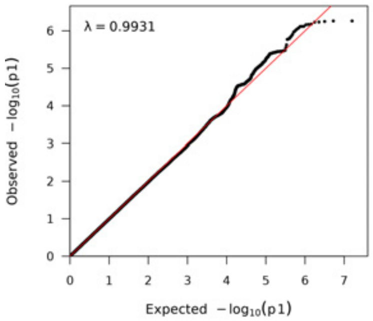


Total testosterone

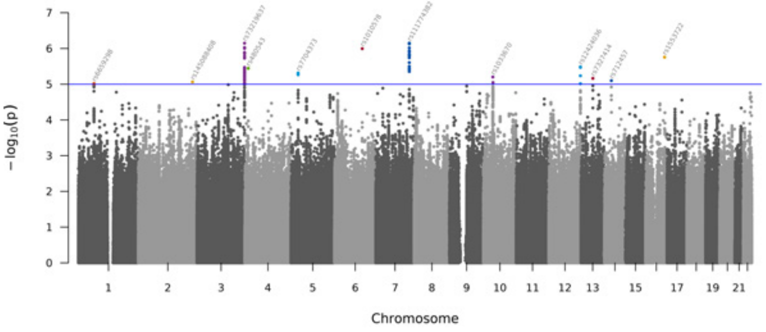


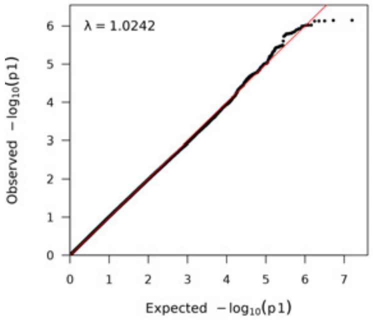

Total bilirubin

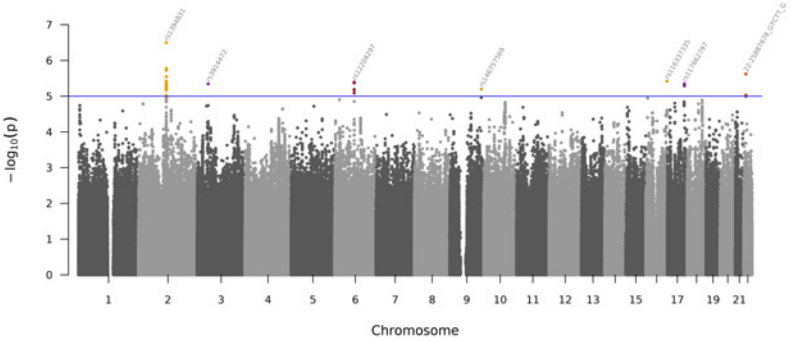


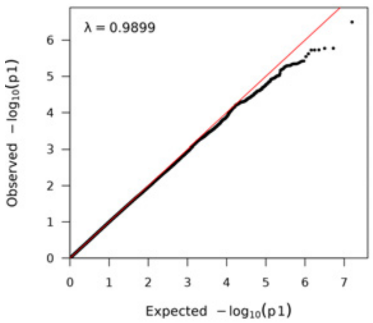


Total protein

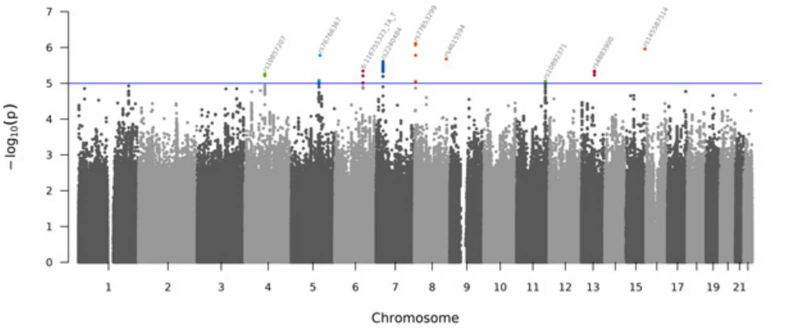


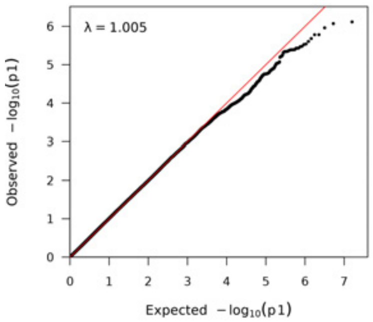


Triglycerides

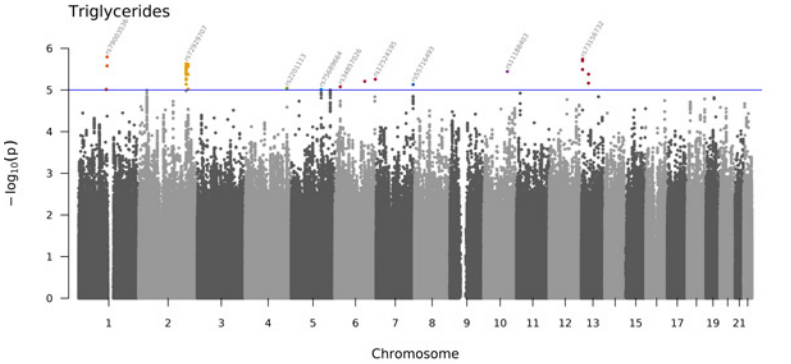


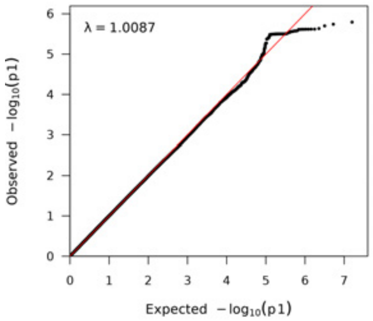


Urate

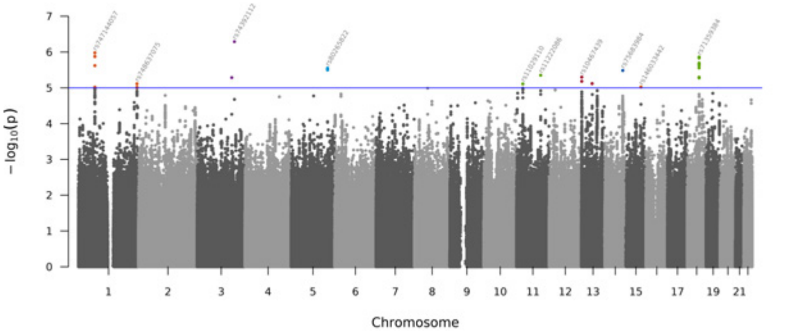


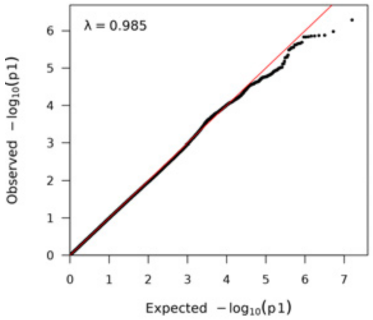

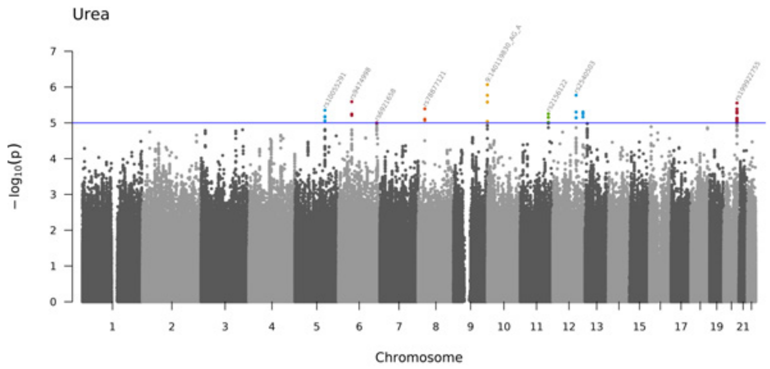
Urea
−log10(p)
Chromosome
1
2
3
4
5
6
7
8
9
10
11
12
13
15
17
19
21

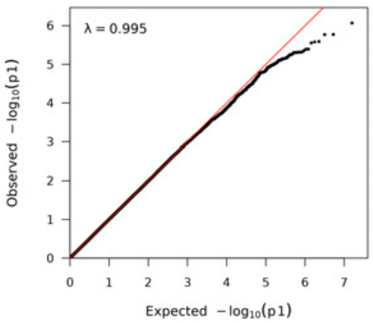
λ = 0.995
Observed −log10(p1)
Expected −log10(p1)

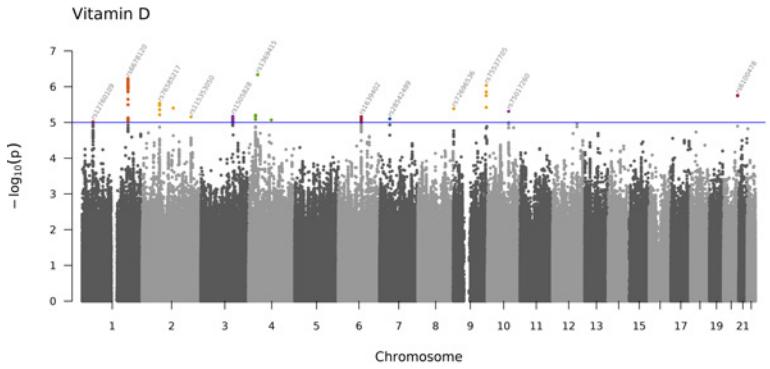
Vitamin D
−log10(p)
Chromosome
1
2
3
4
5
6
7
8
9
10
11
12
13
15
17
19
21

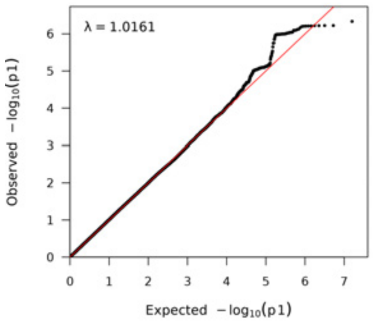
λ = 1.0161
Observed −log10(p1)
Expected −log10(p1)

Alanine aminotransferase (adj-BMI)

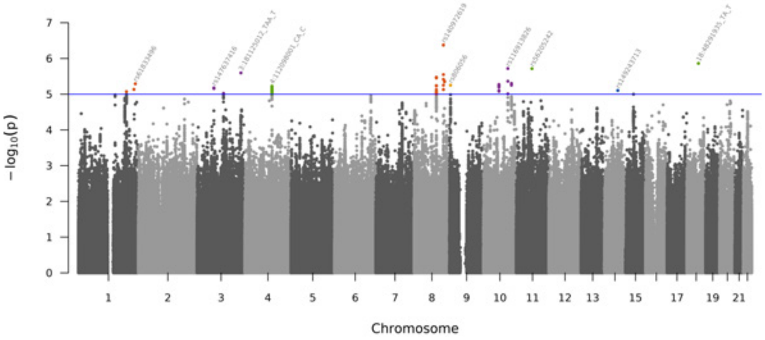


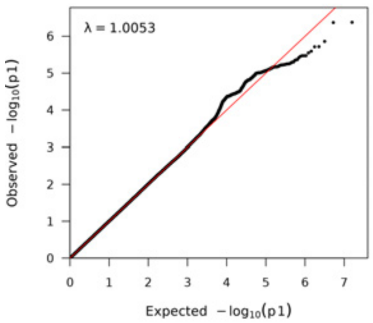


Albumin (adj-BMI)

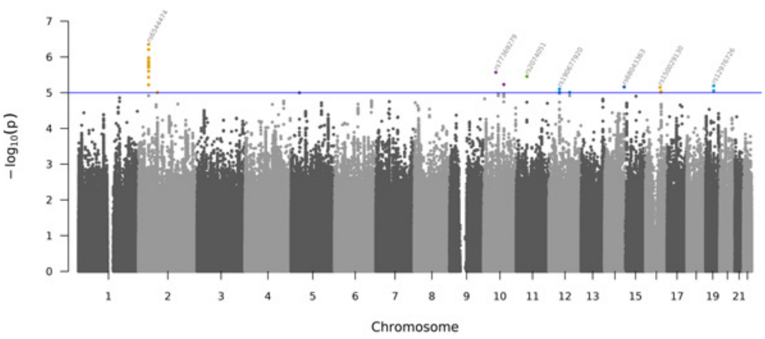


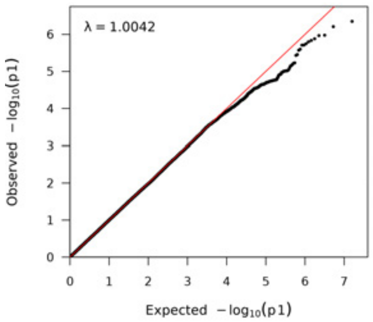


Alkaline phosphatase (adj-BMI)

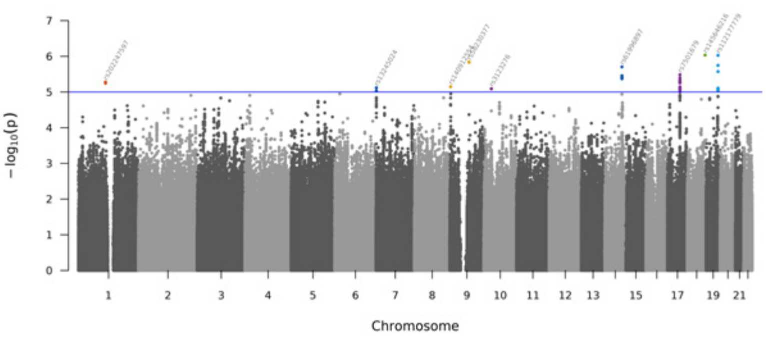


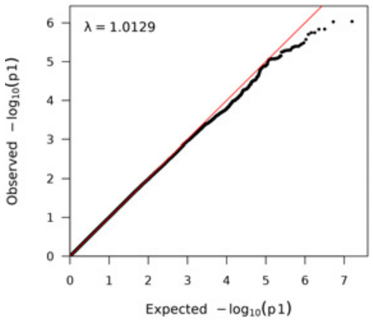


Apolipoprotein A (adj-BMI)

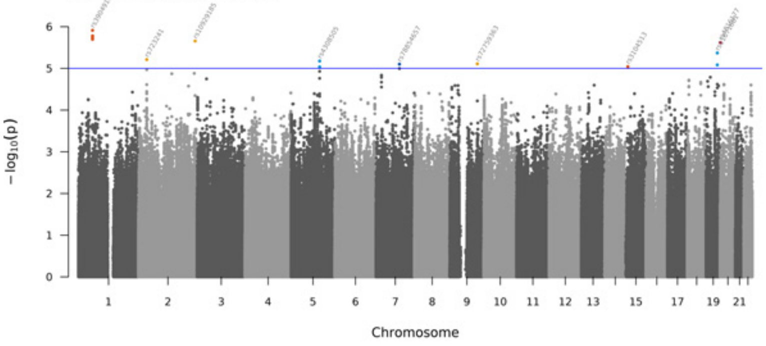


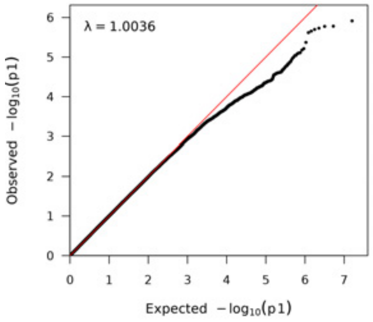

Apolipoprotein B (adj-BMI)

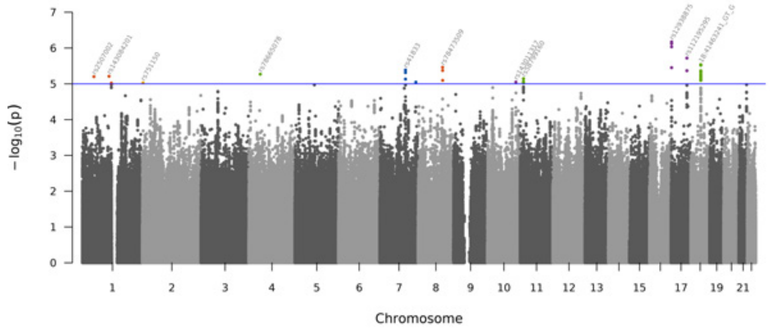


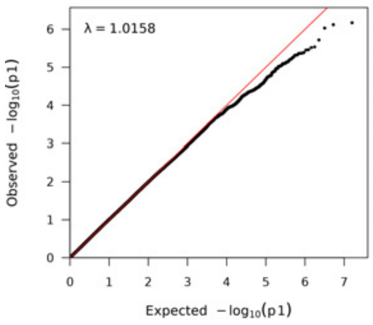


Aspartate aminotransferase (adj-BMI)

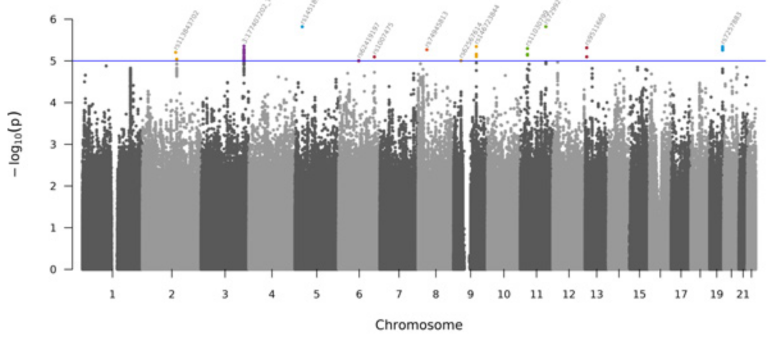


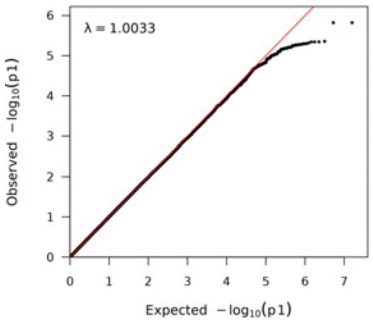


Bioavailable testosterone (adj-BMI)

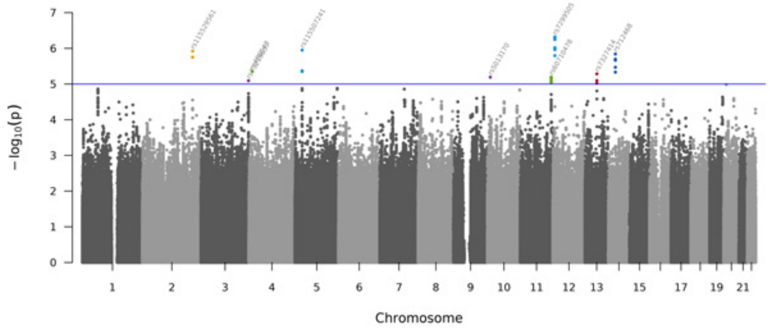


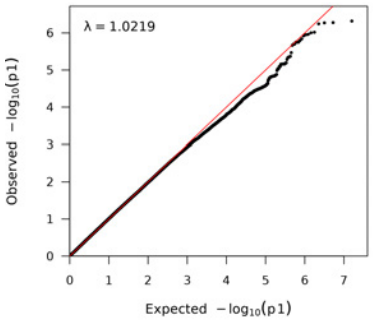


C-reactive protein (adj-BMI)

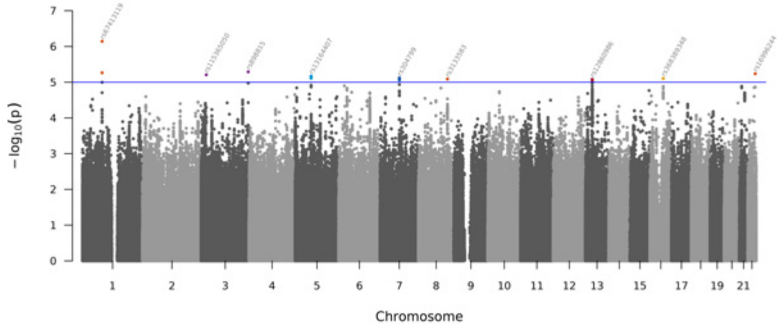


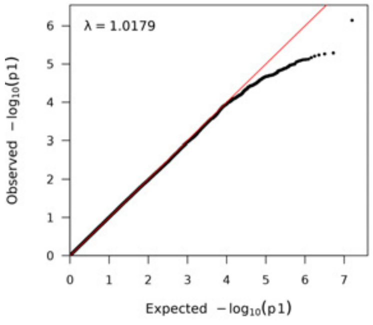

Calcium (adj-BMI)

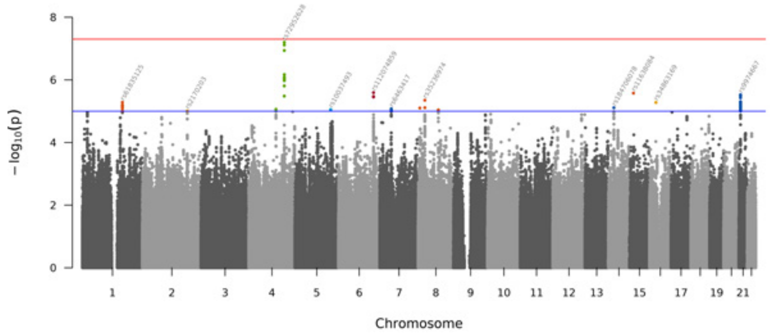


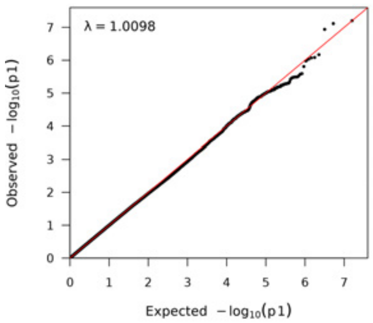


Cholesterol (adj-BMI)

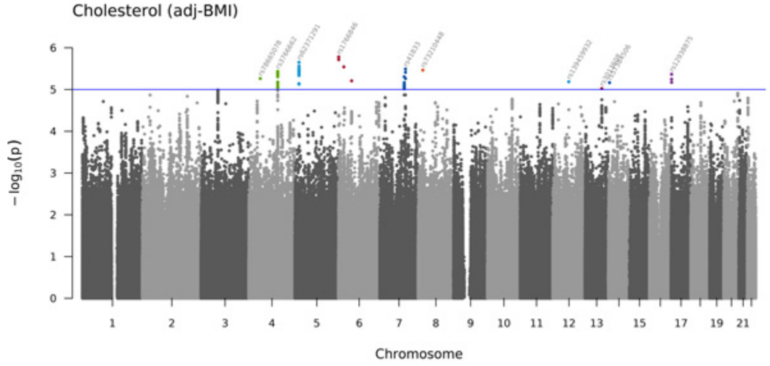


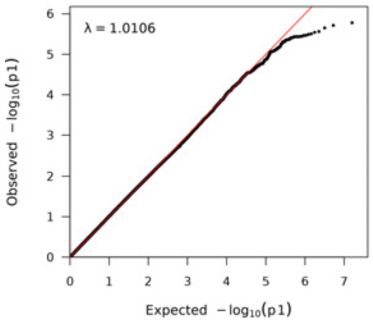


Creatinine (adj-BMI)

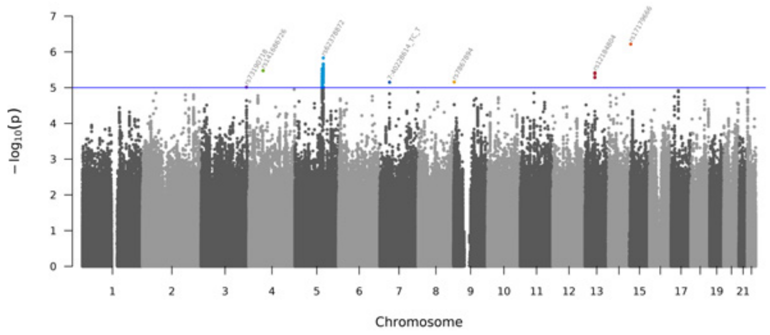


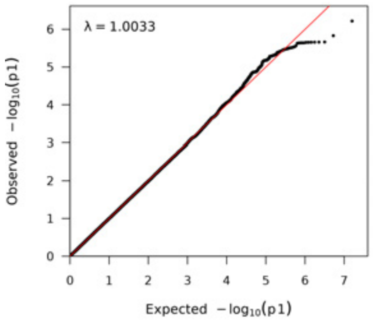


Cystatin C (adj-BMI)

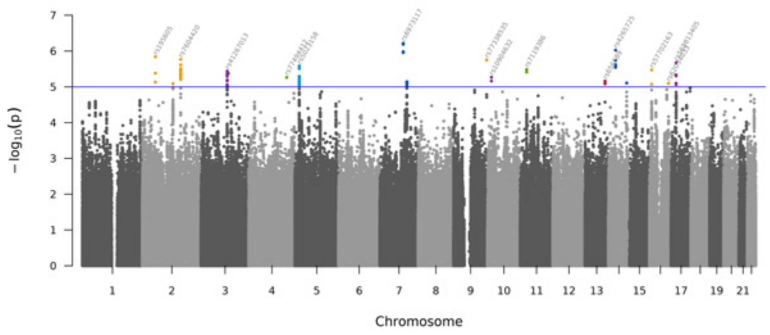


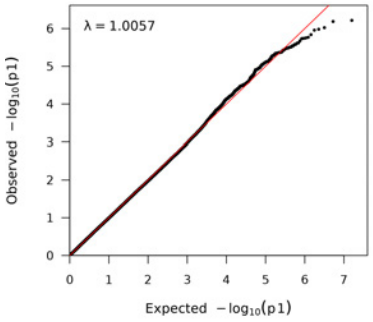

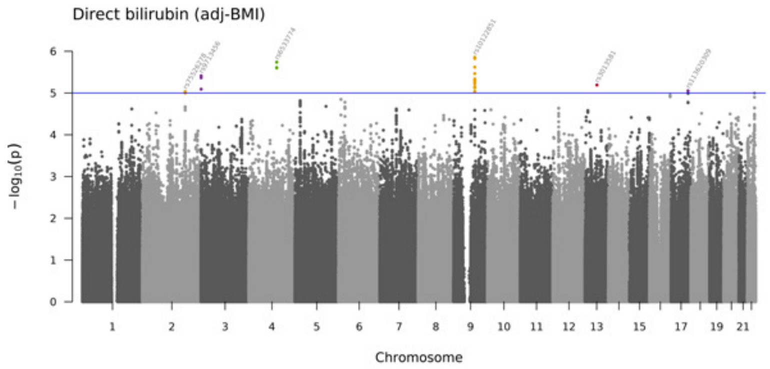
Direct bilirubin (adj-BMI)
$-\log_{10}(p)$
Chromosome

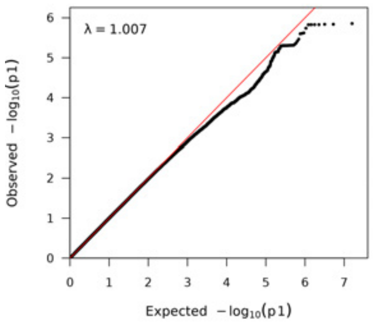
λ = 1.007
Observed $-\log_{10}(p1)$
Expected $-\log_{10}(p1)$

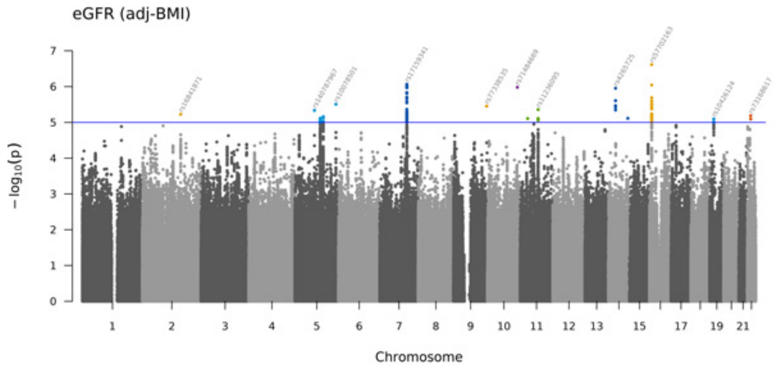
eGFR (adj-BMI)
$-\log_{10}(p)$
Chromosome

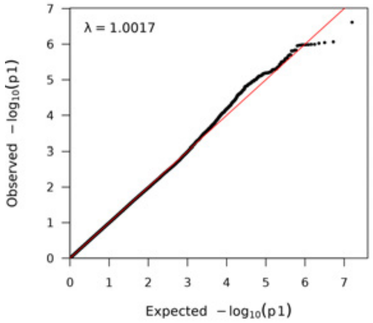
λ = 1.0017
Observed $-\log_{10}(p1)$
Expected $-\log_{10}(p1)$

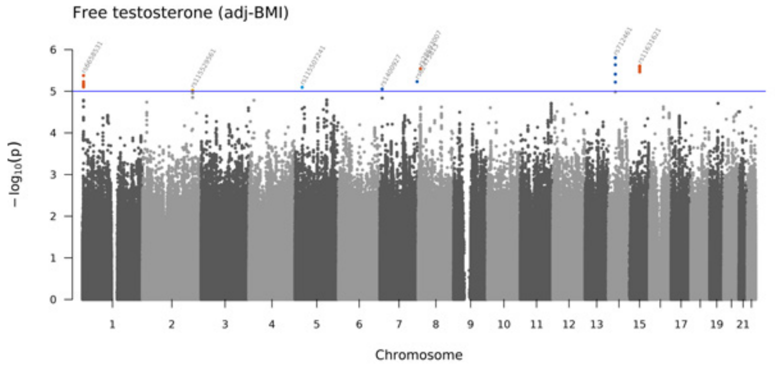
Free testosterone (adj-BMI)
$-\log_{10}(p)$
Chromosome

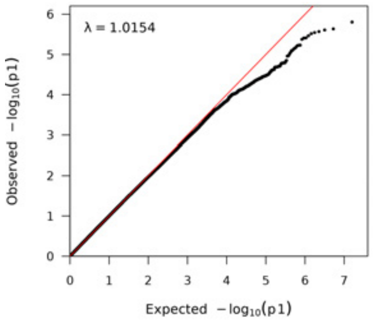
λ = 1.0154
Observed $-\log_{10}(p1)$
Expected $-\log_{10}(p1)$

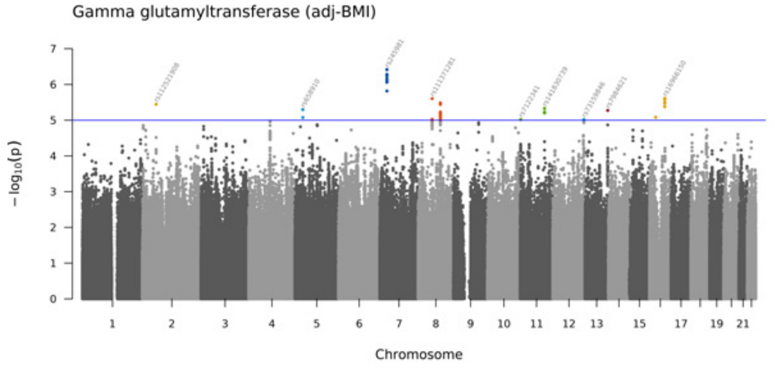
Gamma glutamyltransferase (adj-BMI)
$-\log_{10}(p)$
Chromosome

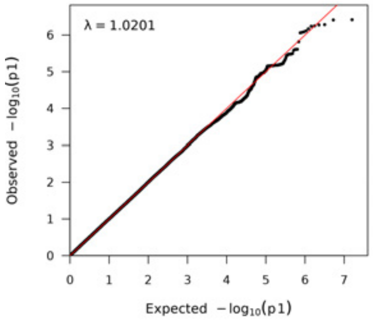
λ = 1.0201
Observed $-\log_{10}(p1)$
Expected $-\log_{10}(p1)$

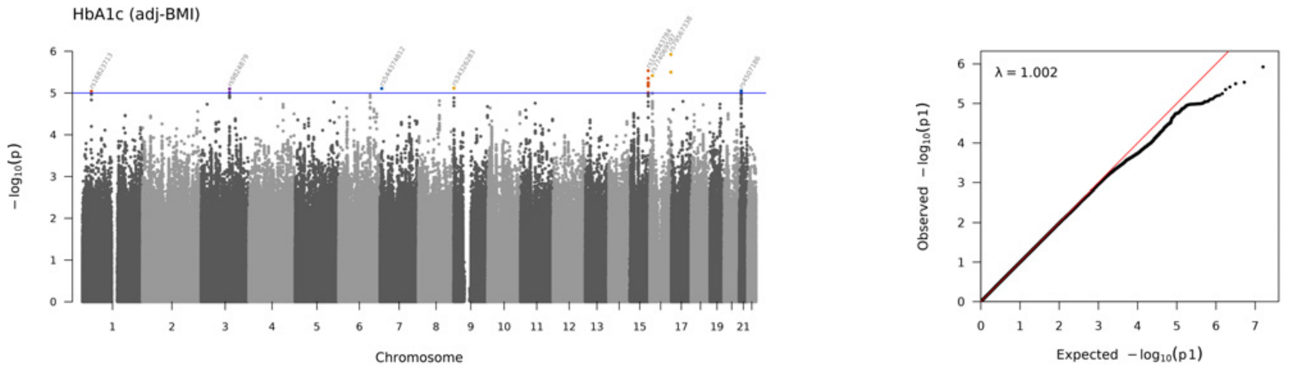
HbA1c (adj-BMI)
$-\log_{10}(p)$
Chromosome
λ = 1.002
Observed $-\log_{10}(p1)$
Expected $-\log_{10}(p1)$

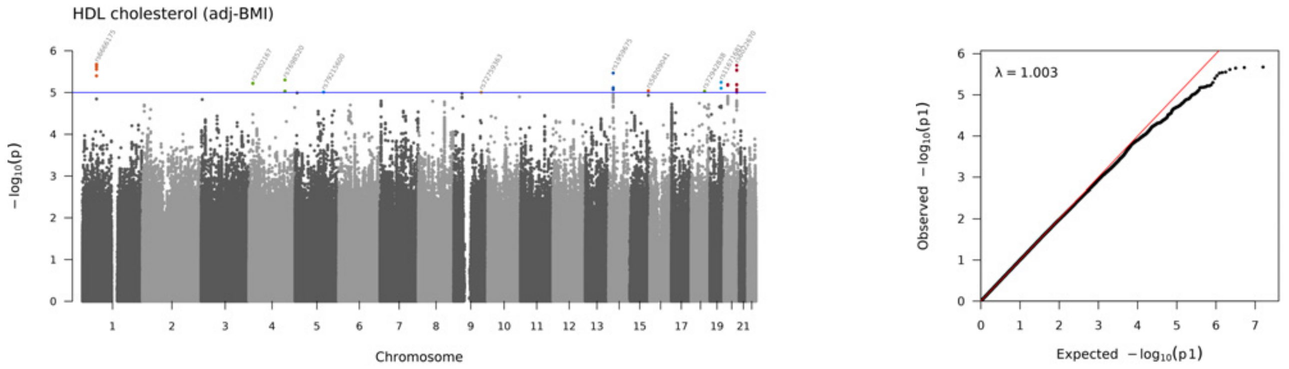
HDL cholesterol (adj-BMI)
$-\log_{10}(p)$
Chromosome
λ = 1.003
Observed $-\log_{10}(p1)$
Expected $-\log_{10}(p1)$

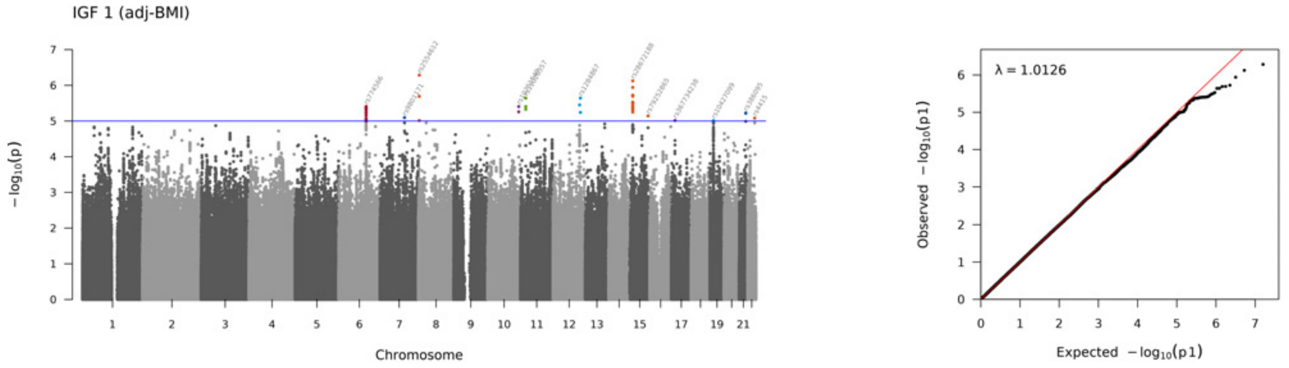
IGF 1 (adj-BMI)
$-\log_{10}(p)$
Chromosome
λ = 1.0126
Observed $-\log_{10}(p1)$
Expected $-\log_{10}(p1)$

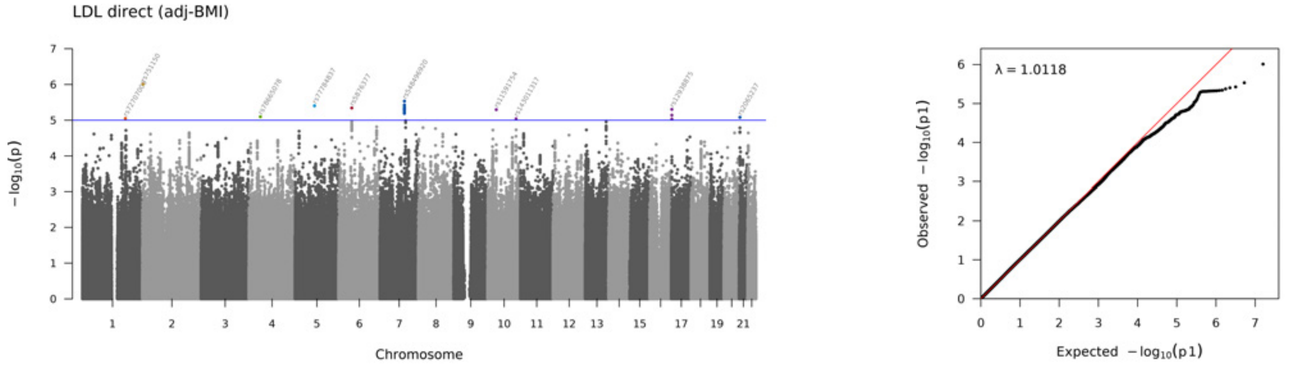
LDL direct (adj-BMI)
$-\log_{10}(p)$
Chromosome
λ = 1.0118
Observed $-\log_{10}(p1)$
Expected $-\log_{10}(p1)$

Lipoprotein A (adj-BMI)

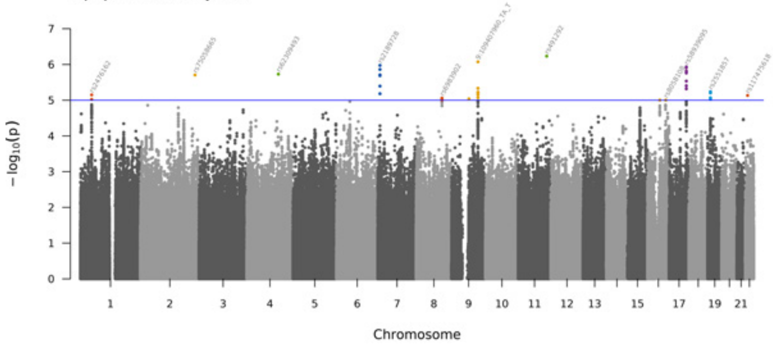


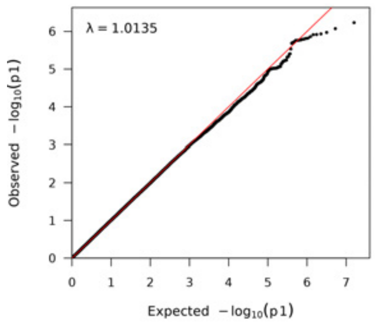


Phosphate (adj-BMI)

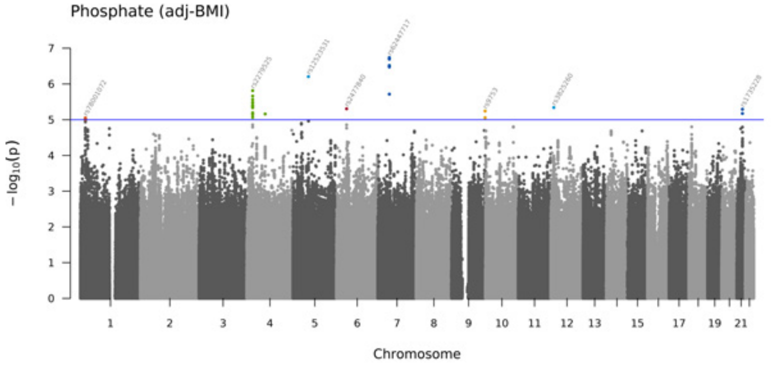


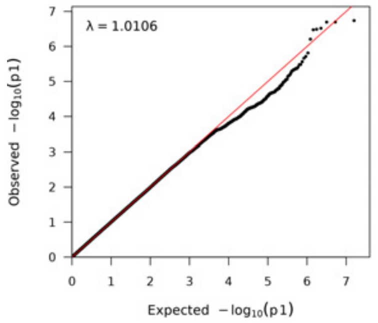


SHBG (adj-BMI)

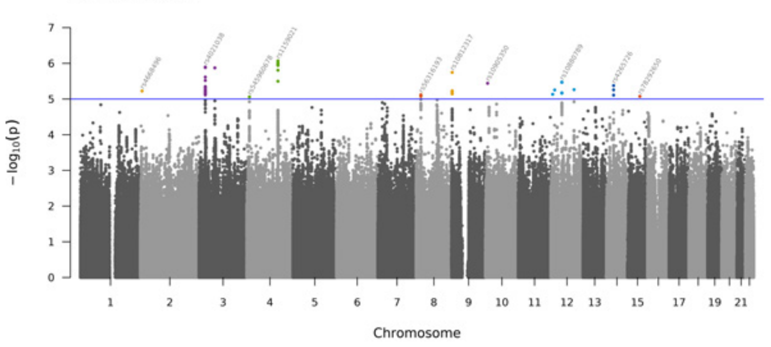


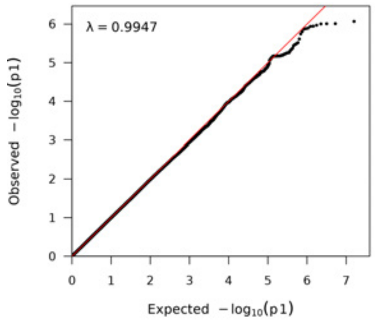


Total testosterone (adj-BMI)

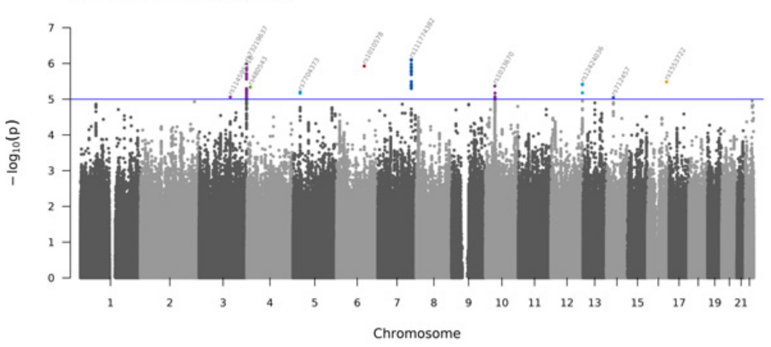


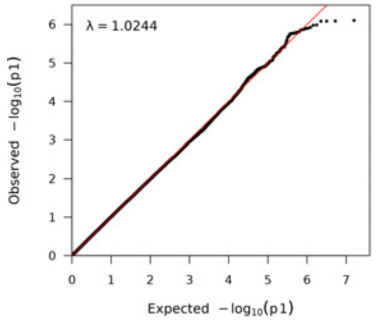

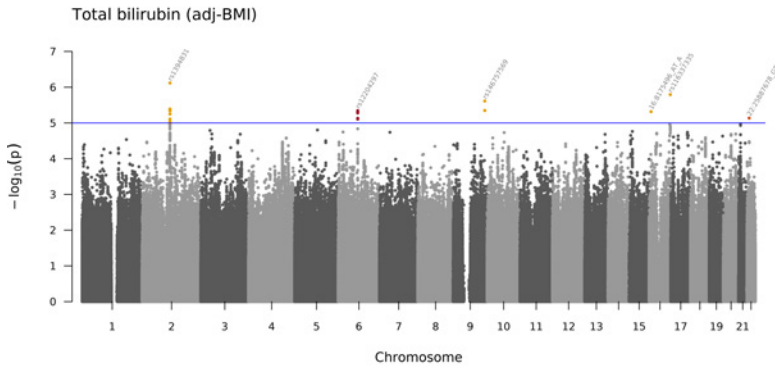
Total bilirubin (adj-BMI)
$-\log_{10}(p)$
Chromosome

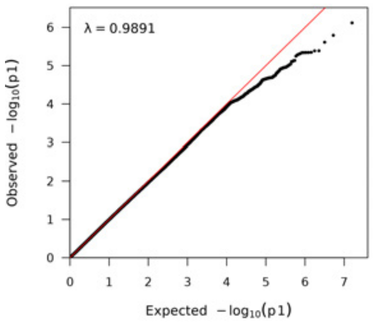
λ = 0.9891
Observed $-\log_{10}(p1)$
Expected $-\log_{10}(p1)$

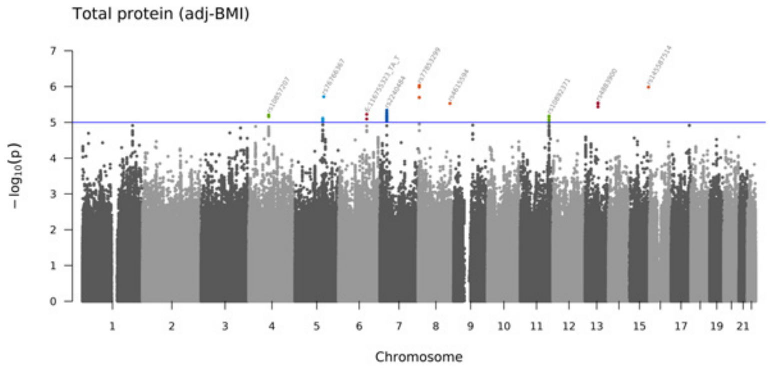
Total protein (adj-BMI)
$-\log_{10}(p)$
Chromosome

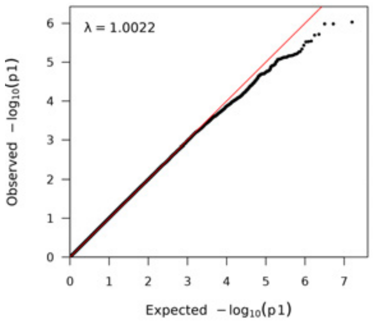
λ = 1.0022
Observed $-\log_{10}(p1)$
Expected $-\log_{10}(p1)$

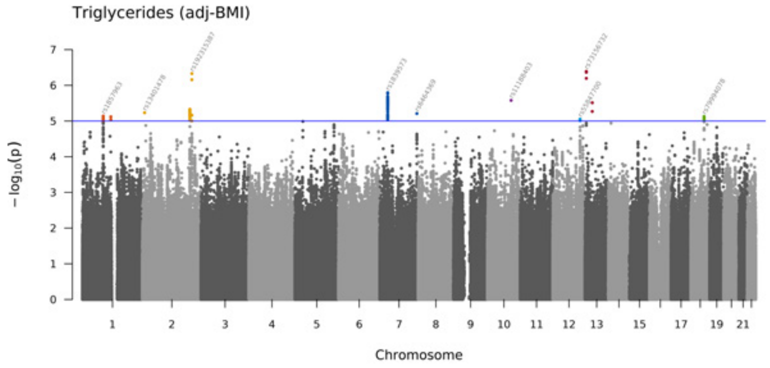
Triglycerides (adj-BMI)
$-\log_{10}(p)$
Chromosome

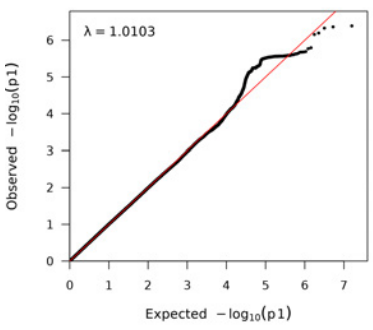
λ = 1.0103
Observed $-\log_{10}(p1)$
Expected $-\log_{10}(p1)$

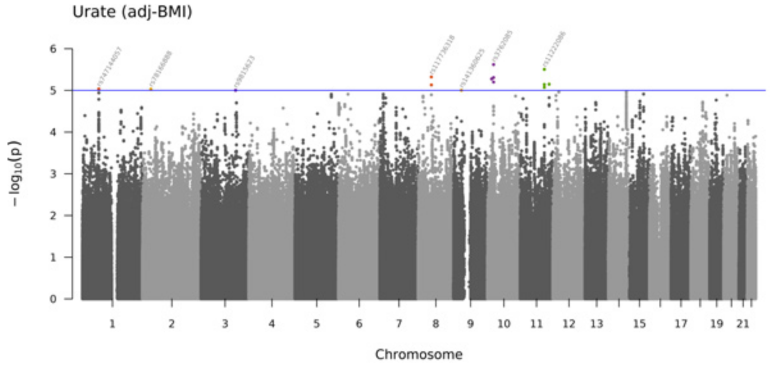
Urate (adj-BMI)
$-\log_{10}(p)$
Chromosome

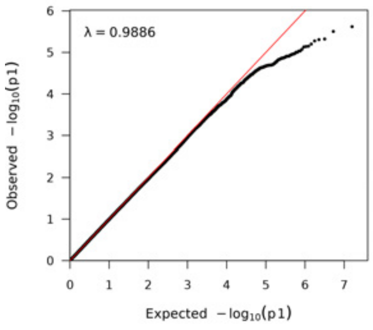
λ = 0.9886
Observed $-\log_{10}(p1)$
Expected $-\log_{10}(p1)$

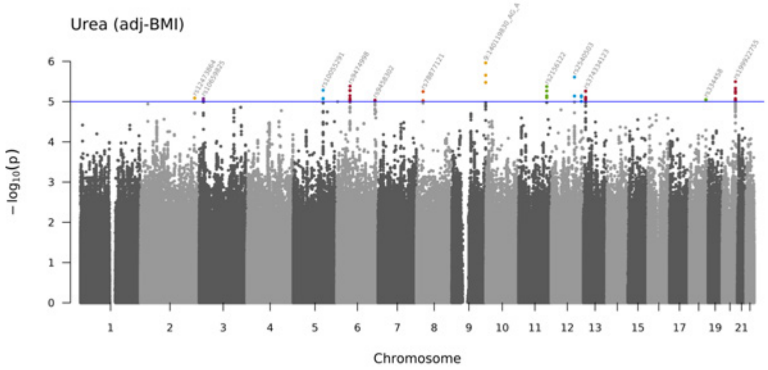
Urea (adj-BMI)
$-\log_{10}(p)$
Chromosome

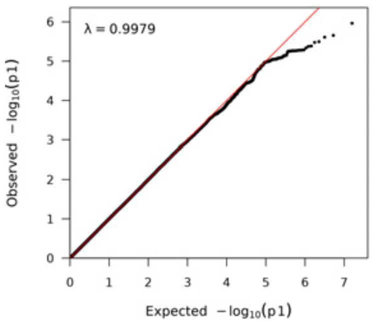
λ = 0.9979
Observed $-\log_{10}(p1)$
Expected $-\log_{10}(p1)$

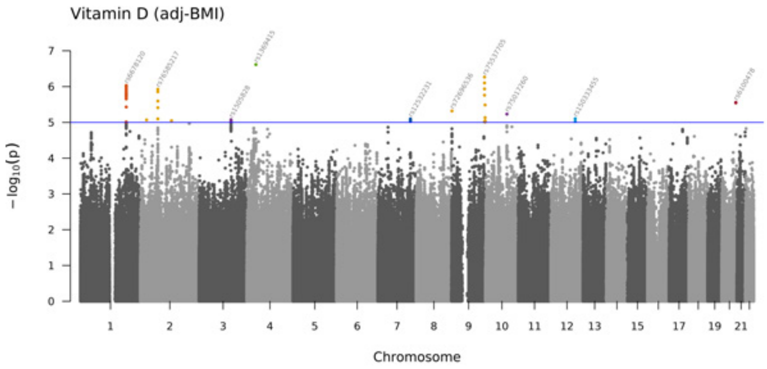
Vitamin D (adj-BMI)
$-\log_{10}(p)$
Chromosome

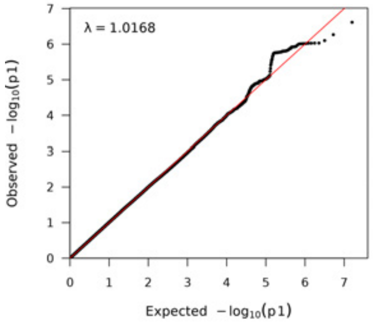
λ = 1.0168
Observed $-\log_{10}(p1)$
Expected $-\log_{10}(p1)$

## S7

MMAA (ENSG00000151611.13) and rs72952628 (chr4_145716082_C_T_b38)

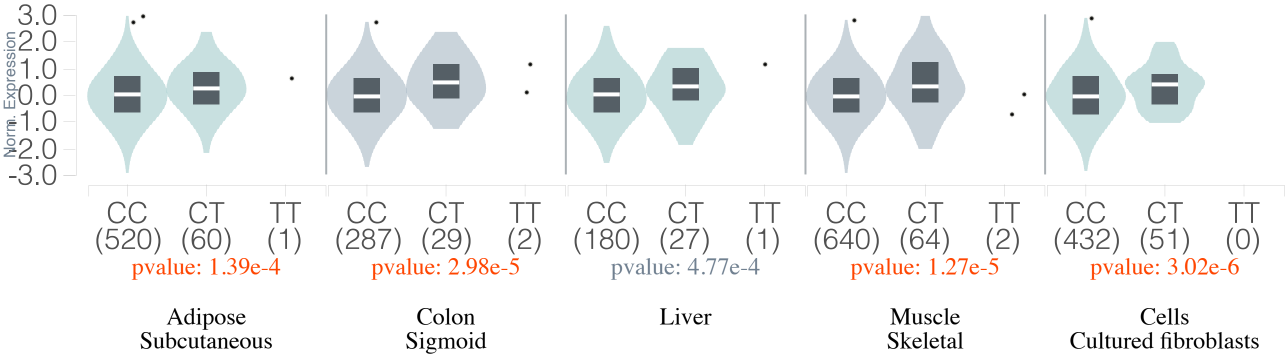

## S8A

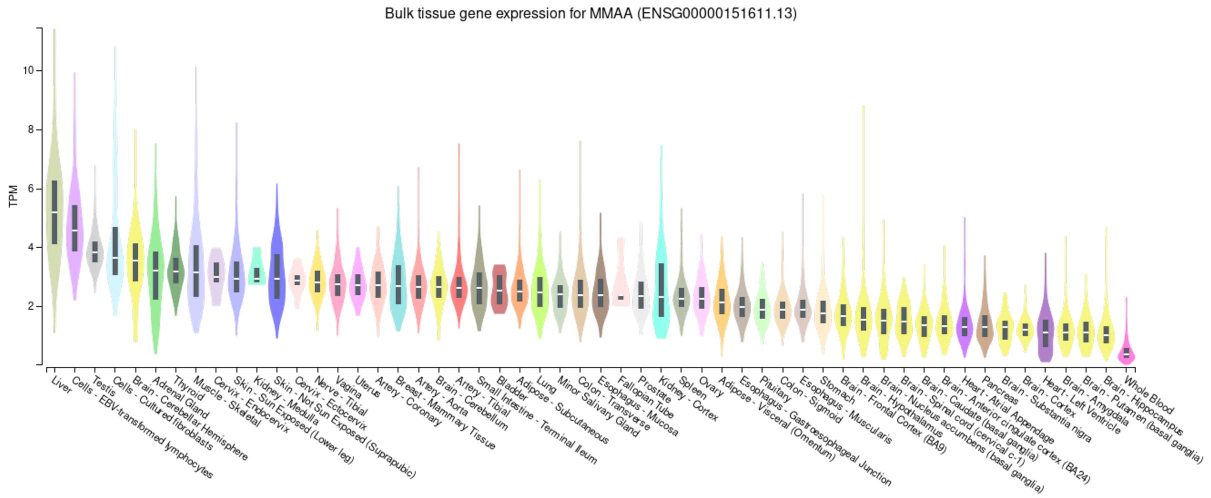
Bulk tissue gene expression for MMAA (ENSG00000151611.13)
TPM
10
8
6
4
2

## S8B

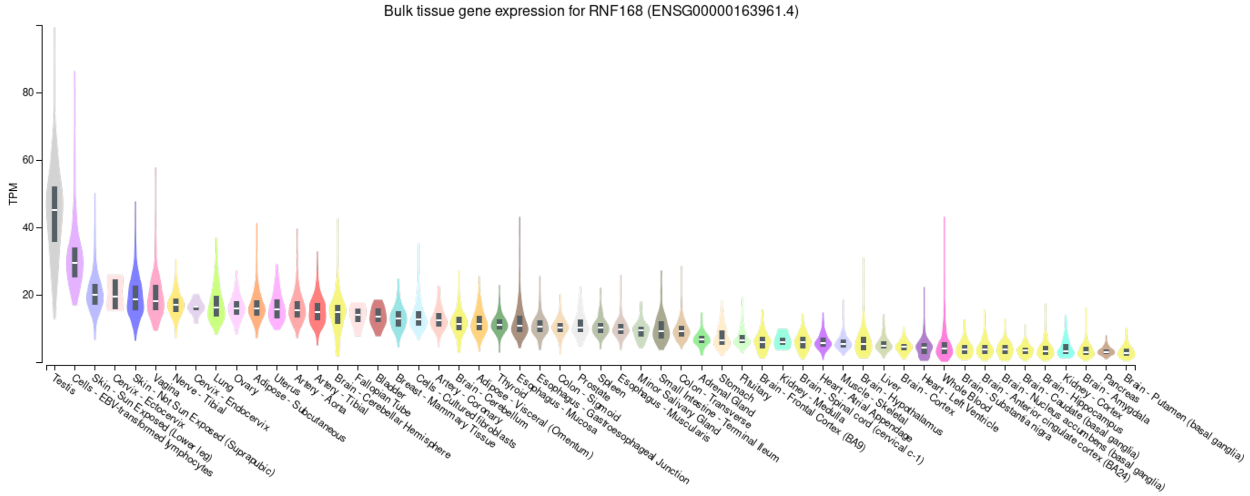
Bulk tissue gene expression for RNF168 (ENSG00000163961.4)
TPM
80
60
40
20

## S8C

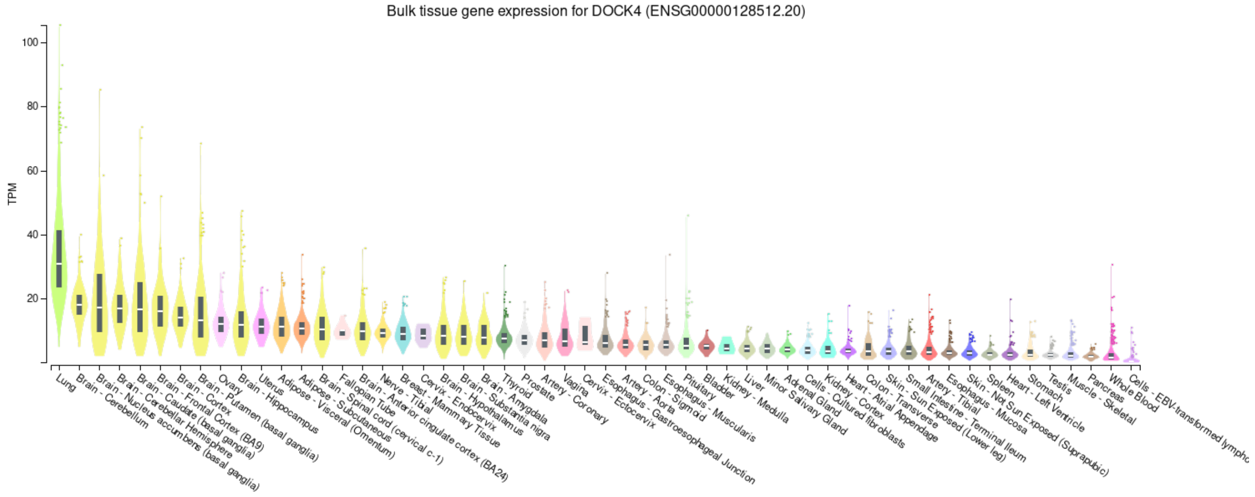
Bulk tissue gene expression for DOCK4 (ENSG00000128512.20)
TPM
100
80
60
40
20

**S9**

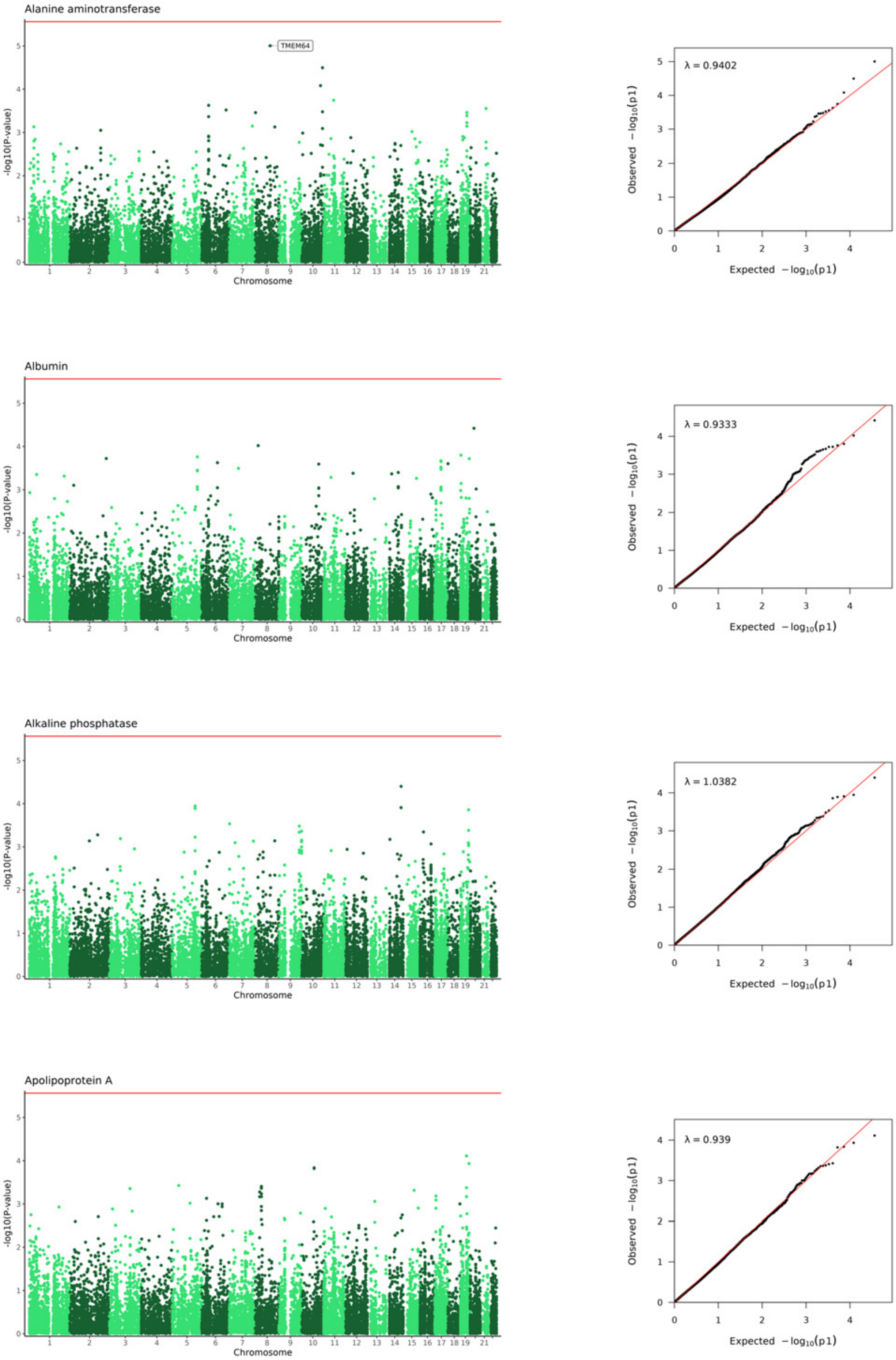
Alanine aminotransferase
TMEM64
-log10(P-value)
Chromosome
λ = 0.9402
Observed −log10(p1)
Expected −log10(p1)
Albumin
-log10(P-value)
Chromosome
λ = 0.9333
Observed −log10(p1)
Expected −log10(p1)
Alkaline phosphatase
-log10(P-value)
Chromosome
λ = 1.0382
Observed −log10(p1)
Expected −log10(p1)
Apolipoprotein A
-log10(P-value)
Chromosome
λ = 0.939
Observed −log10(p1)
Expected −log10(p1)

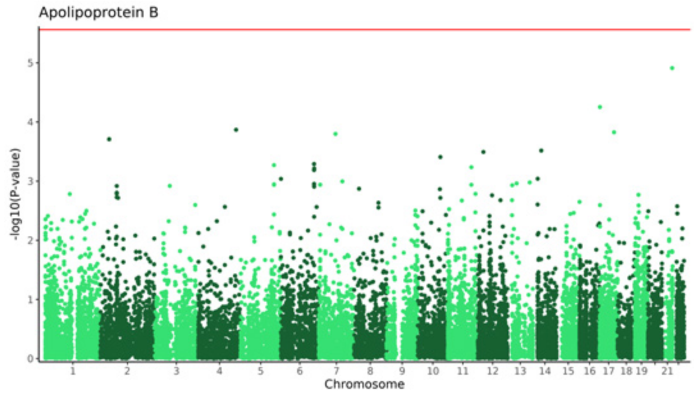
Apolipoprotein B
-log10(P-value)
Chromosome

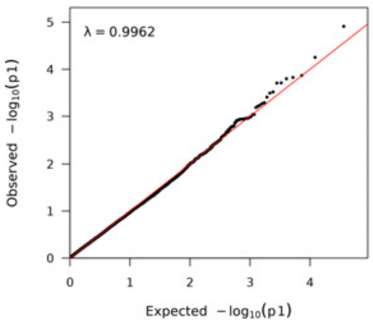
λ = 0.9962
Observed $-\log_{10}(p1)$
Expected $-\log_{10}(p1)$

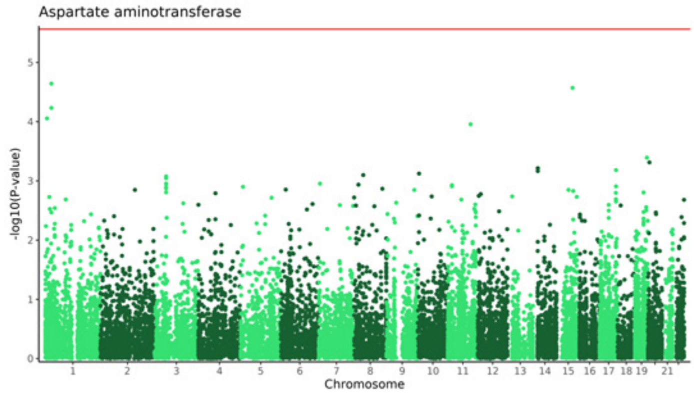
Aspartate aminotransferase
-log10(P-value)
Chromosome

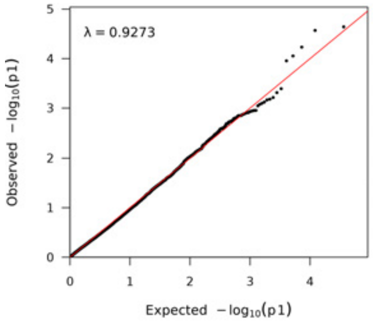
λ = 0.9273
Observed $-\log_{10}(p1)$
Expected $-\log_{10}(p1)$

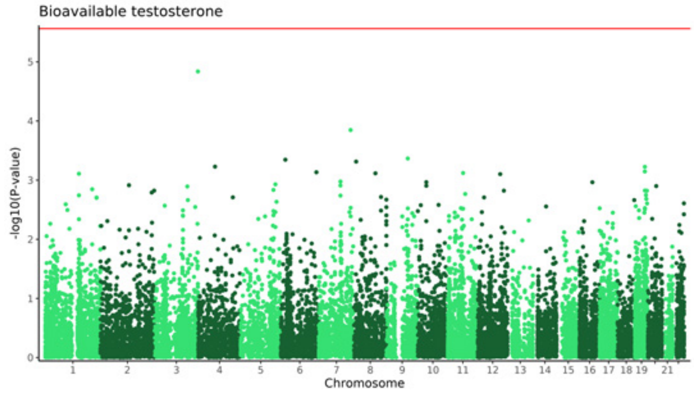
Bioavailable testosterone
-log10(P-value)
Chromosome

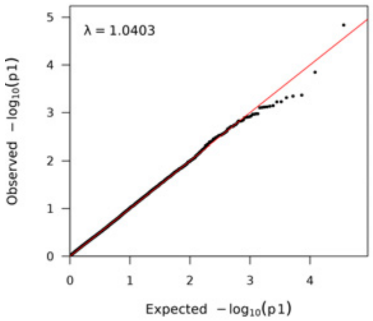
λ = 1.0403
Observed $-\log_{10}(p1)$
Expected $-\log_{10}(p1)$

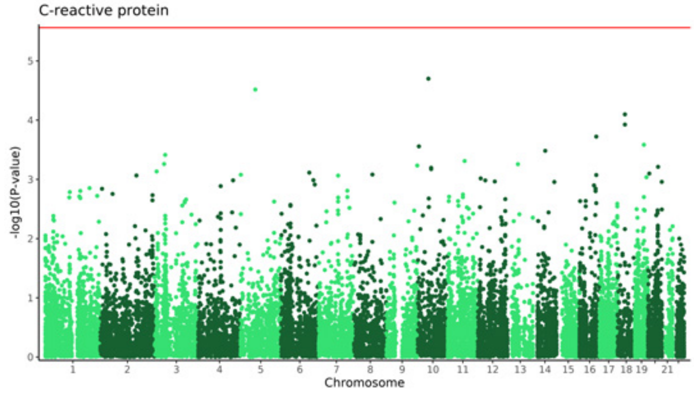
C-reactive protein
-log10(P-value)
Chromosome

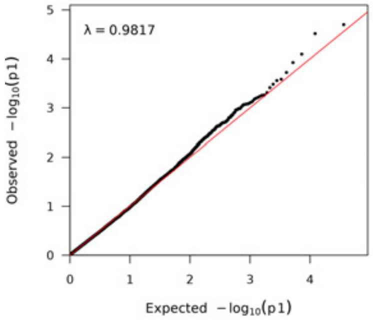
λ = 0.9817
Observed $-\log_{10}(p1)$
Expected $-\log_{10}(p1)$

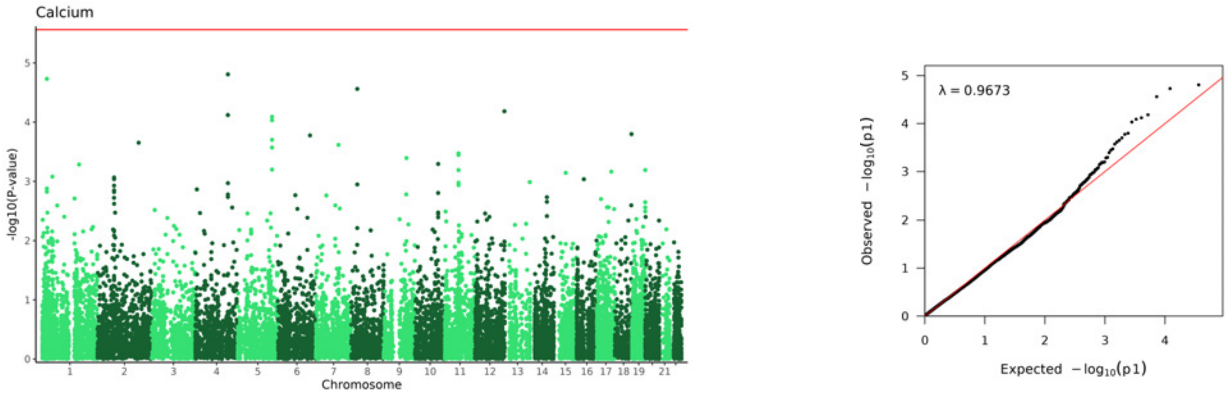
Calcium
-log10(P-value)
Chromosome
λ = 0.9673
Observed −log10(p1)
Expected −log10(p1)

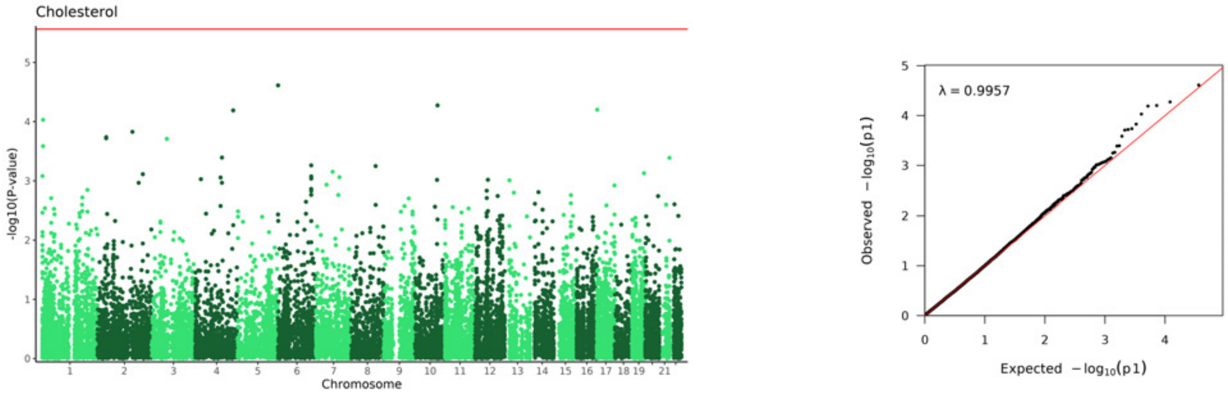
Cholesterol
-log10(P-value)
Chromosome
λ = 0.9957
Observed −log10(p1)
Expected −log10(p1)

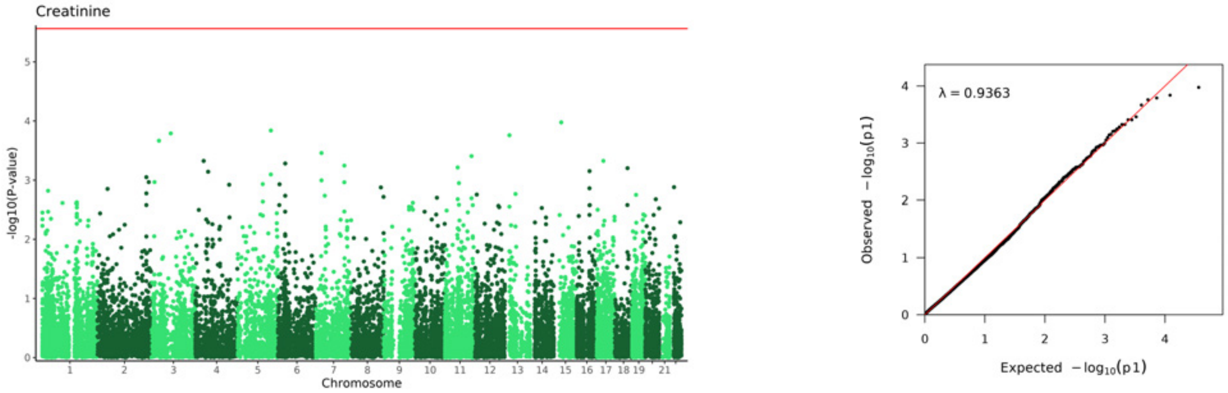
Creatinine
-log10(P-value)
Chromosome
λ = 0.9363
Observed −log10(p1)
Expected −log10(p1)

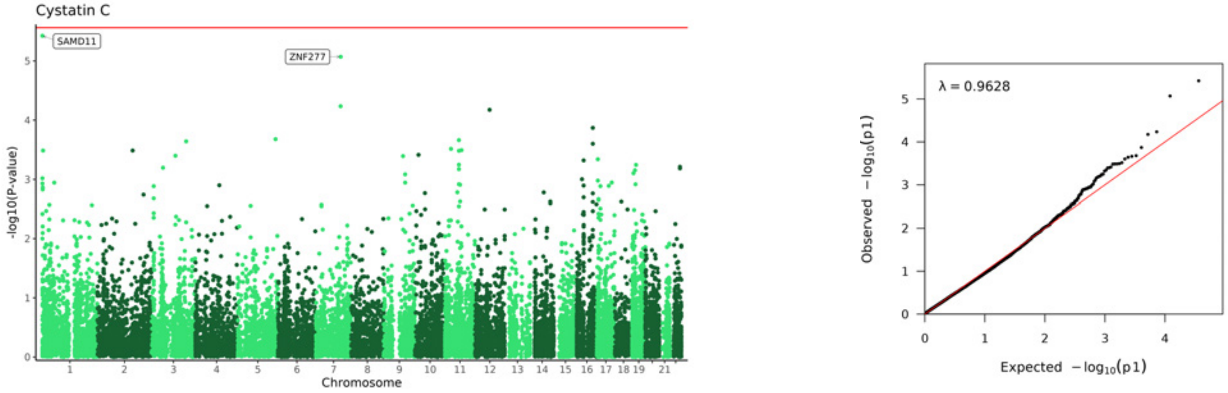
Cystatin C
SAMD11
ZNF277
-log10(P-value)
Chromosome
λ = 0.9628
Observed −log10(p1)
Expected −log10(p1)

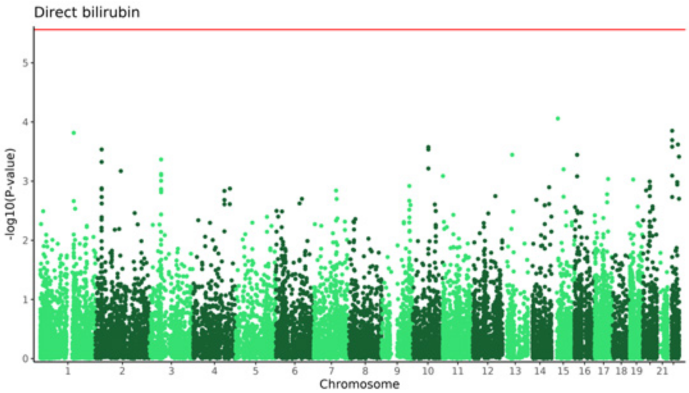
Direct bilirubin
-log10(P-value)
Chromosome

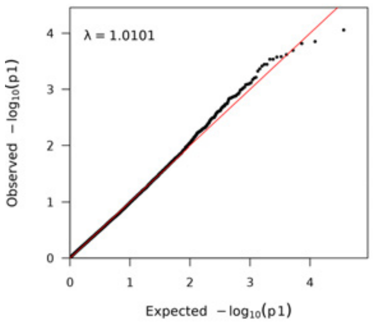
λ = 1.0101
Observed −log10(p1)
Expected −log10(p1)

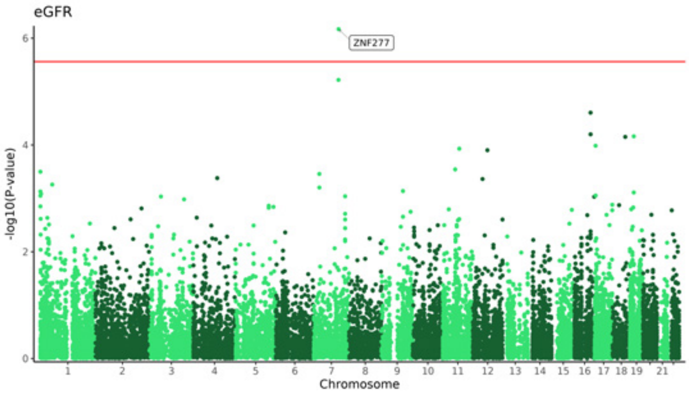
eGFR
ZNF277
-log10(P-value)
Chromosome

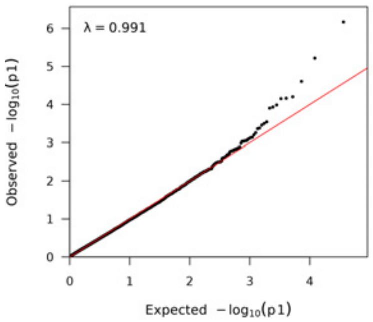
λ = 0.991
Observed −log10(p1)
Expected −log10(p1)

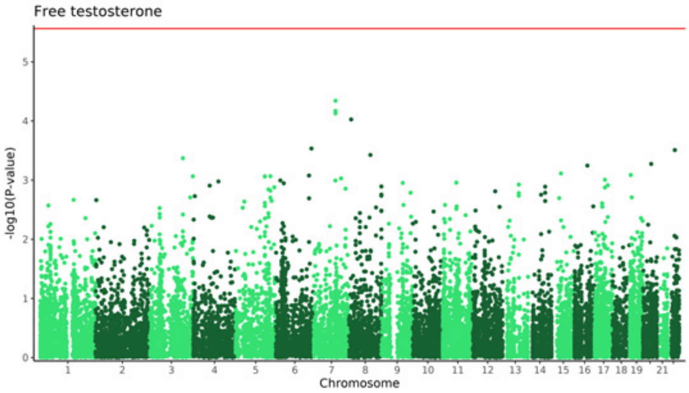
Free testosterone
-log10(P-value)
Chromosome

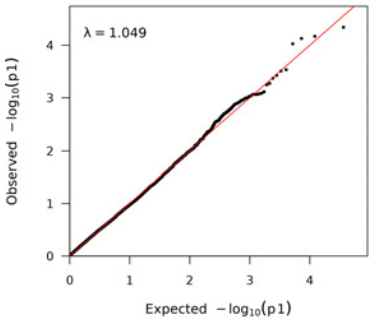
λ = 1.049
Observed −log10(p1)
Expected −log10(p1)

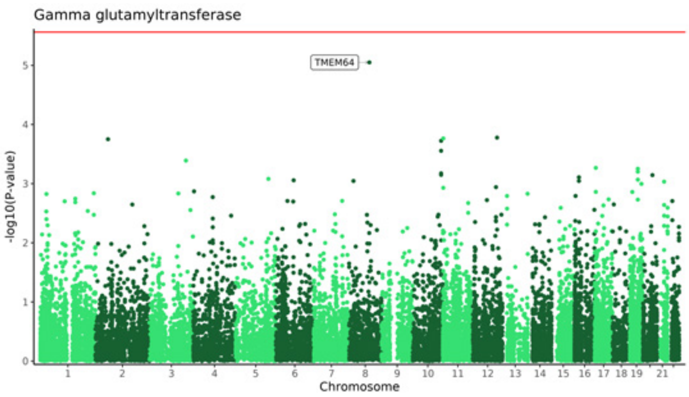
Gamma glutamyltransferase
TMEM64
-log10(P-value)
Chromosome

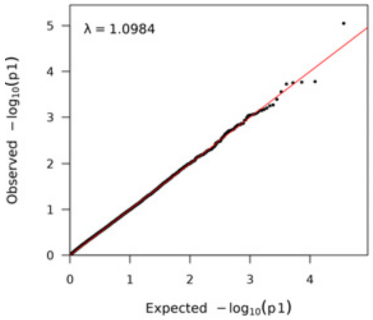
λ = 1.0984
Observed −log10(p1)
Expected −log10(p1)

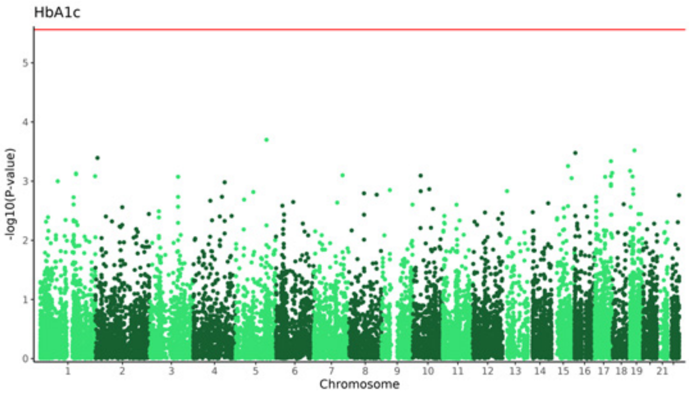
HbA1c
-log10(P-value)
Chromosome

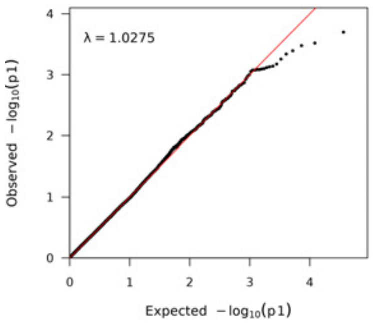
λ = 1.0275
Observed $-\log_{10}(p1)$
Expected $-\log_{10}(p1)$

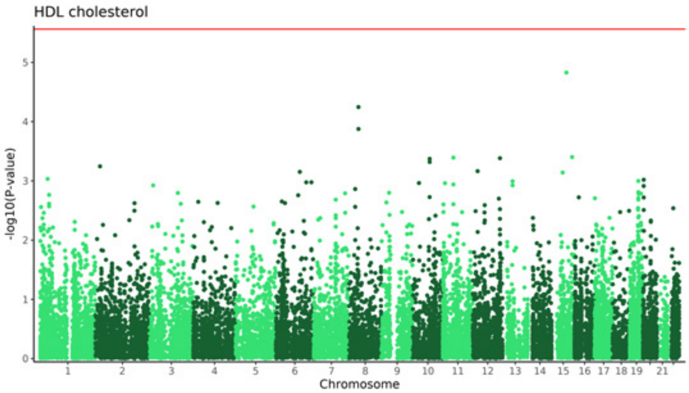
HDL cholesterol
-log10(P-value)
Chromosome

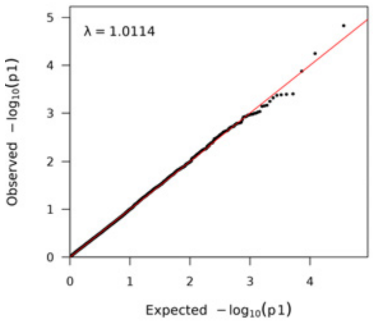
λ = 1.0114
Observed $-\log_{10}(p1)$
Expected $-\log_{10}(p1)$

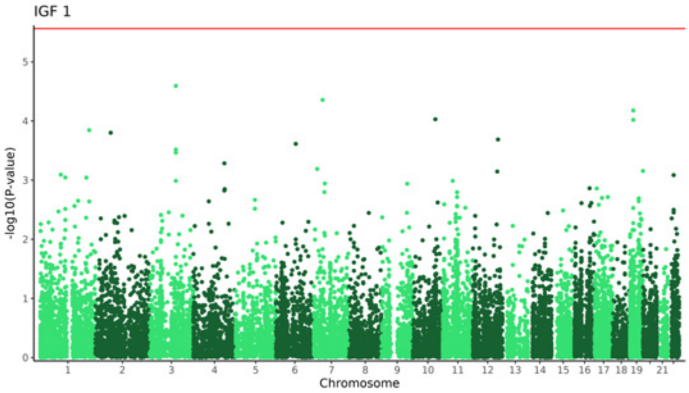
IGF 1
-log10(P-value)
Chromosome

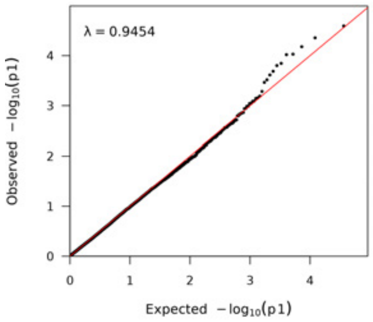
λ = 0.9454
Observed $-\log_{10}(p1)$
Expected $-\log_{10}(p1)$

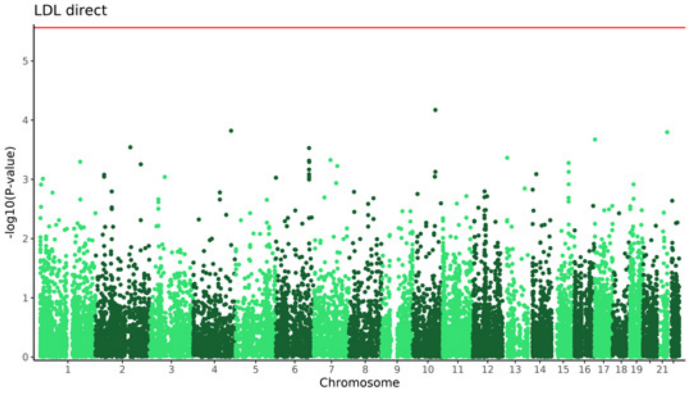
LDL direct
-log10(P-value)
Chromosome

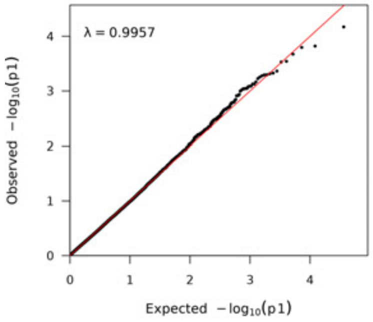
λ = 0.9957
Observed $-\log_{10}(p1)$
Expected $-\log_{10}(p1)$

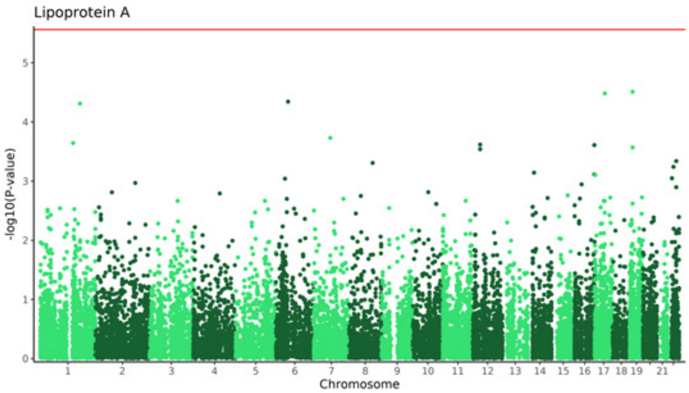
Lipoprotein A
-log10(P-value)
Chromosome

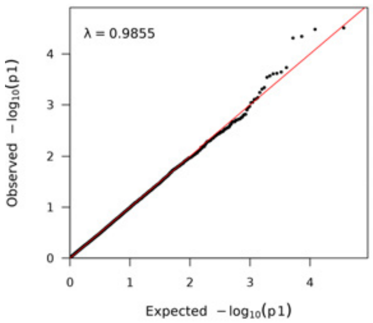
λ = 0.9855
Observed −log10(p1)
Expected −log10(p1)

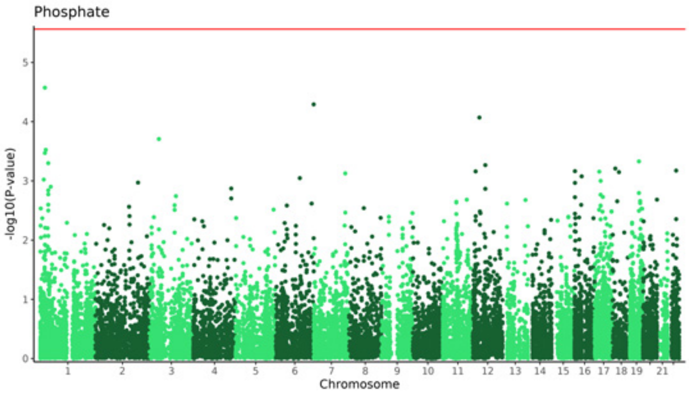
Phosphate
-log10(P-value)
Chromosome

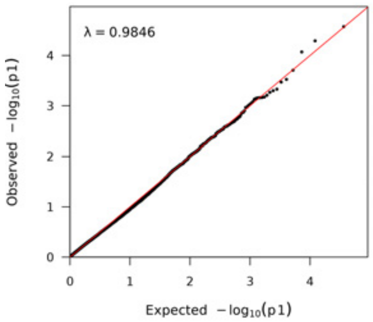
λ = 0.9846
Observed −log10(p1)
Expected −log10(p1)

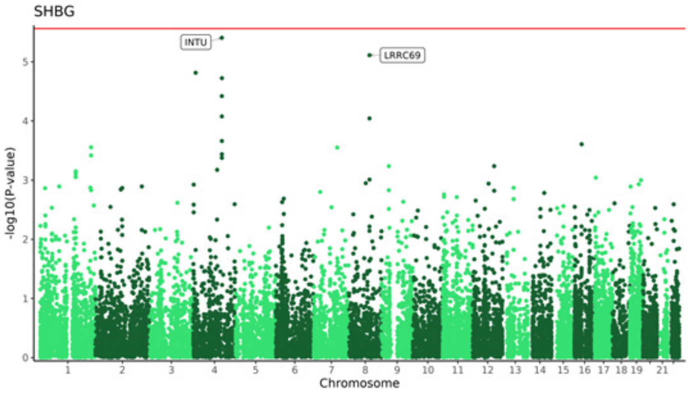
SHBG
INTU
LRRC69
-log10(P-value)
Chromosome

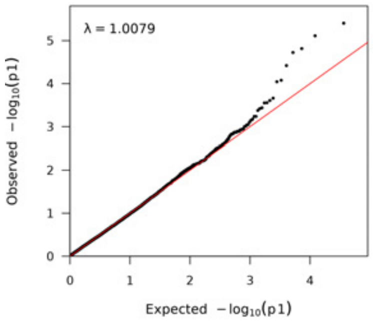
λ = 1.0079
Observed −log10(p1)
Expected −log10(p1)

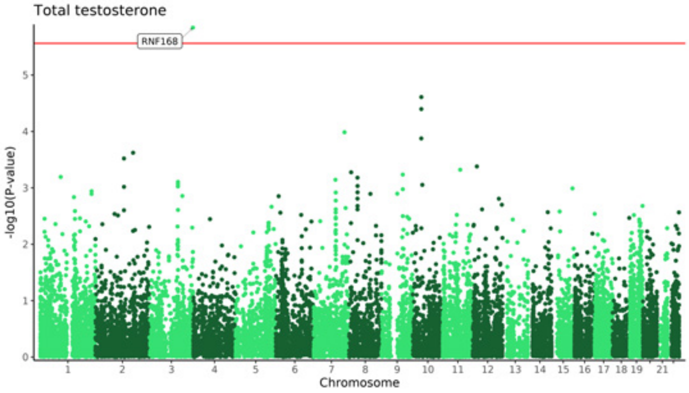
Total testosterone
RNF168
-log10(P-value)
Chromosome

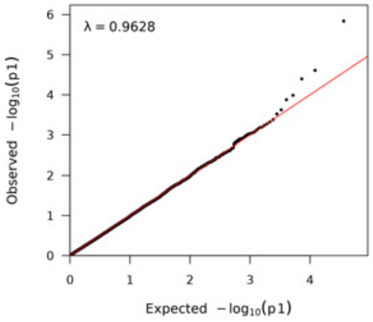
λ = 0.9628
Observed −log10(p1)
Expected −log10(p1)

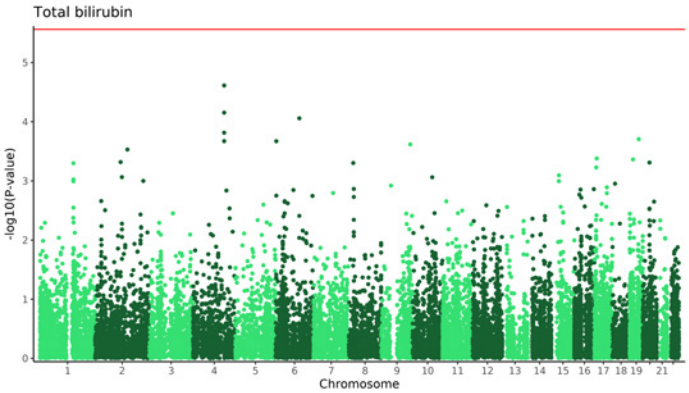
Total bilirubin
-log10(P-value)
Chromosome

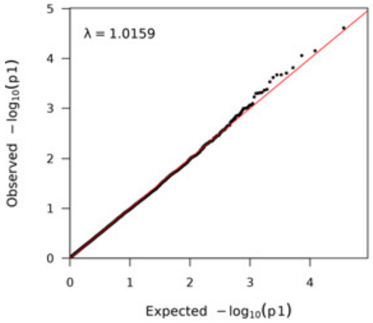
λ = 1.0159
Observed $-\log_{10}(p1)$
Expected $-\log_{10}(p1)$

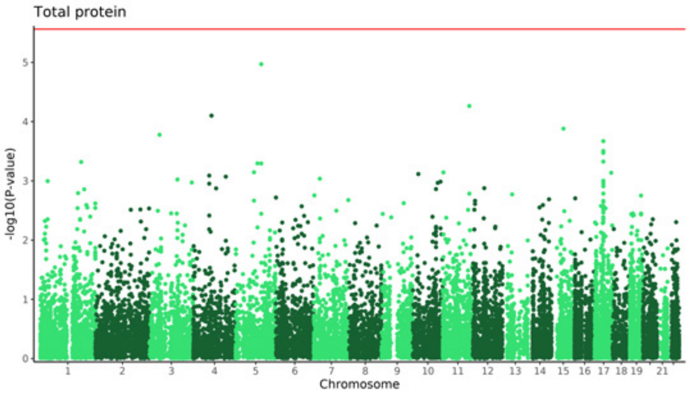
Total protein
-log10(P-value)
Chromosome

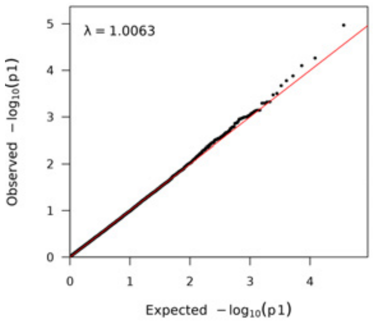
λ = 1.0063
Observed $-\log_{10}(p1)$
Expected $-\log_{10}(p1)$

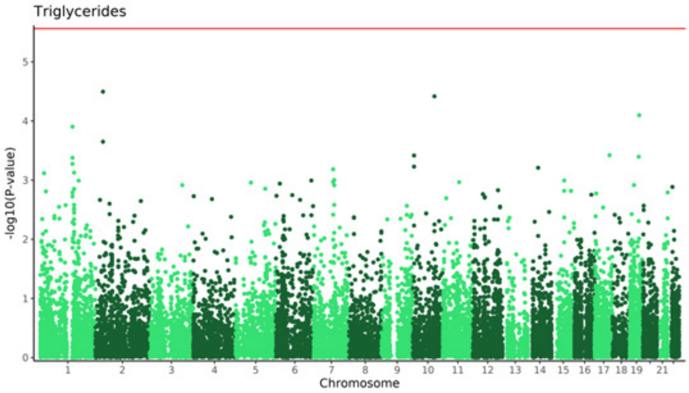
Triglycerides
-log10(P-value)
Chromosome

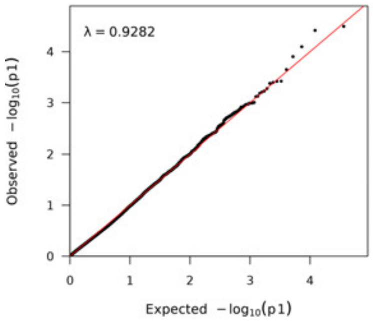
λ = 0.9282
Observed $-\log_{10}(p1)$
Expected $-\log_{10}(p1)$

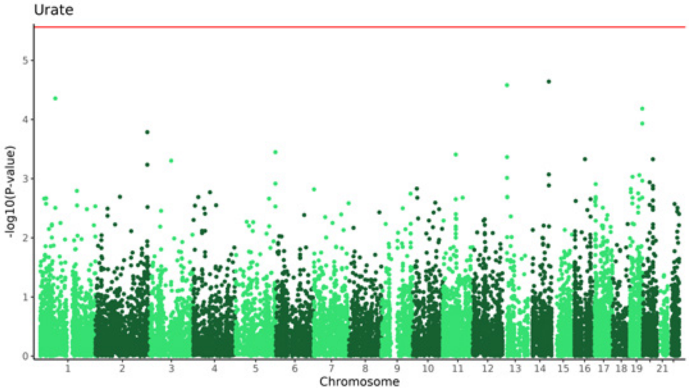
Urate
-log10(P-value)
Chromosome

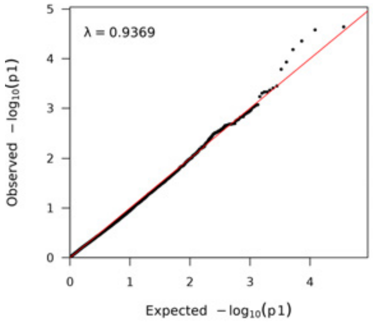
λ = 0.9369
Observed $-\log_{10}(p1)$
Expected $-\log_{10}(p1)$

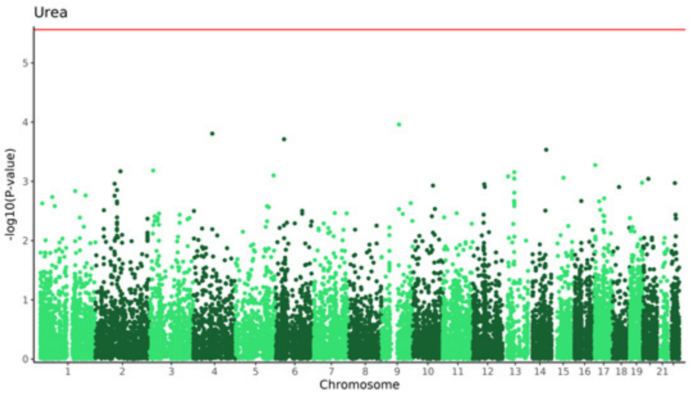
Urea
-log10(P-value)
Chromosome

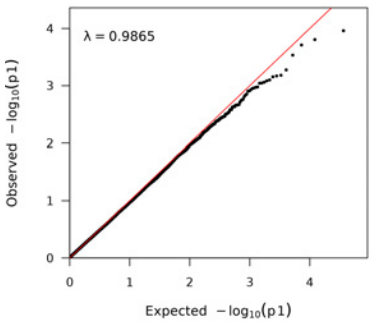
λ = 0.9865
Observed $-\log_{10}(p1)$
Expected $-\log_{10}(p1)$

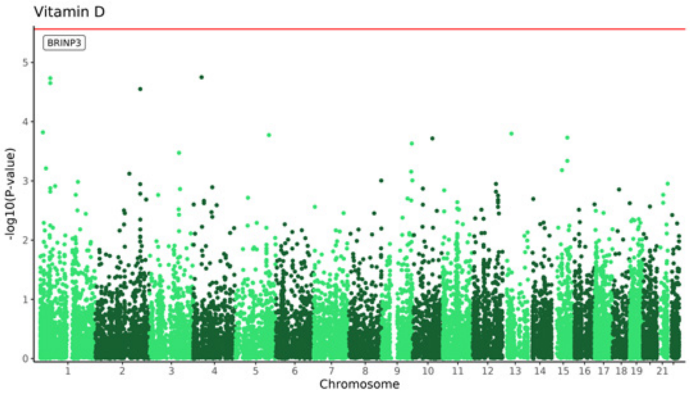
Vitamin D
BRINP3
-log10(P-value)
Chromosome

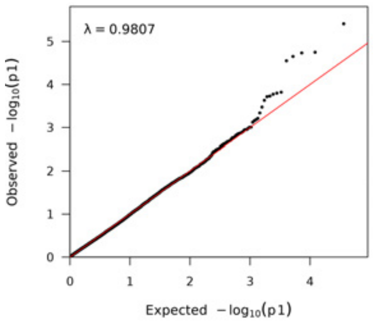
λ = 0.9807
Observed $-\log_{10}(p1)$
Expected $-\log_{10}(p1)$

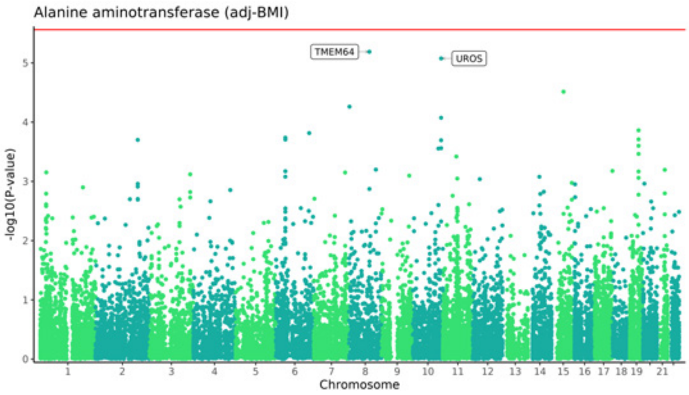
Alanine aminotransferase (adj-BMI)
TMEM64
UROS
-log10(P-value)
Chromosome

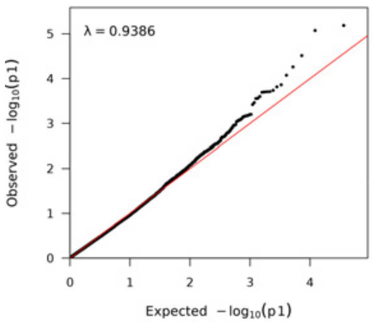
λ = 0.9386
Observed $-\log_{10}(p1)$
Expected $-\log_{10}(p1)$

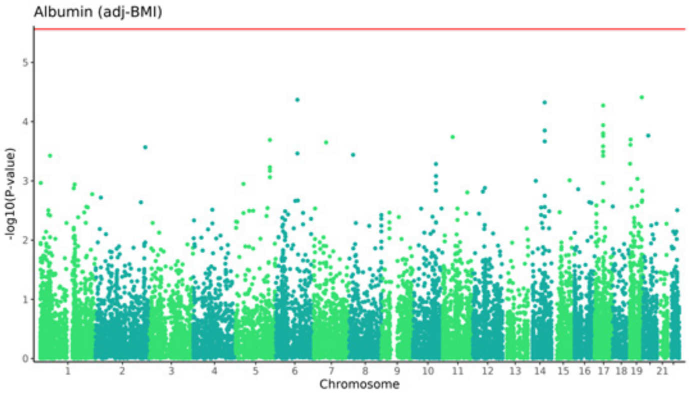
Albumin (adj-BMI)
-log10(P-value)
Chromosome

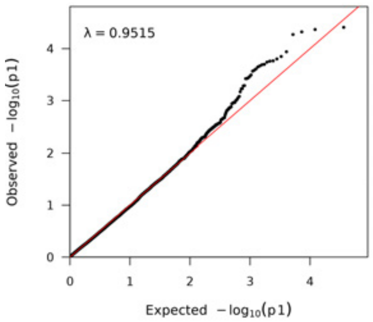
λ = 0.9515
Observed $-\log_{10}(p1)$
Expected $-\log_{10}(p1)$

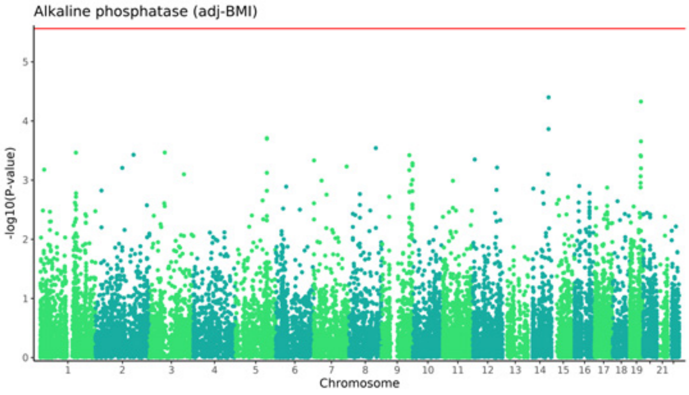
Alkaline phosphatase (adj-BMI)
-log10(P-value)
Chromosome

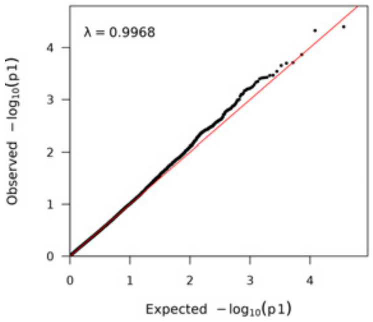
λ = 0.9968
Observed $-\log_{10}(p1)$
Expected $-\log_{10}(p1)$

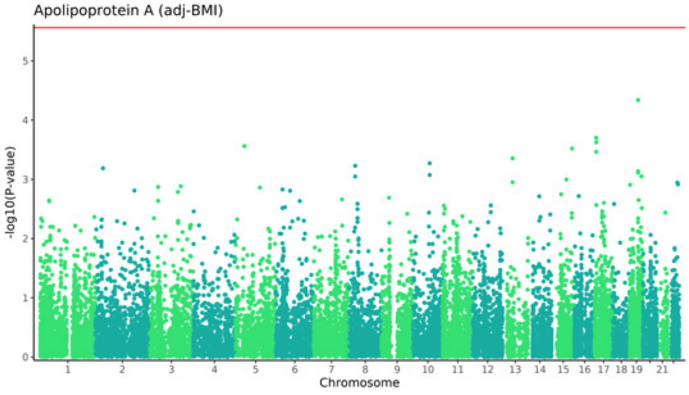
Apolipoprotein A (adj-BMI)
-log10(P-value)
Chromosome

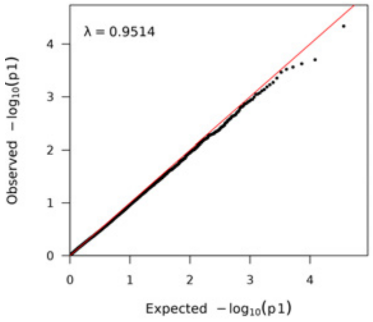
λ = 0.9514
Observed $-\log_{10}(p1)$
Expected $-\log_{10}(p1)$

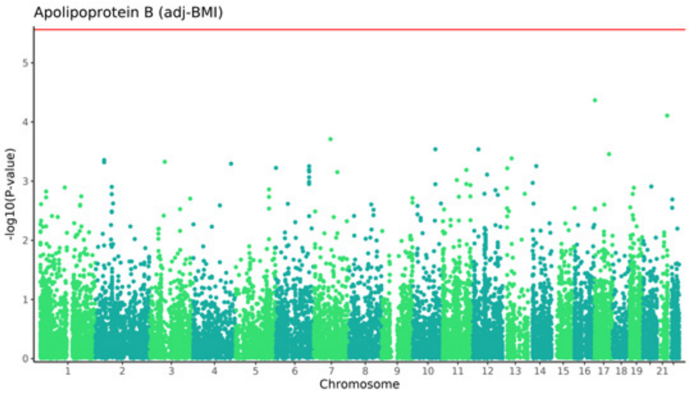
Apolipoprotein B (adj-BMI)
-log10(P-value)
Chromosome

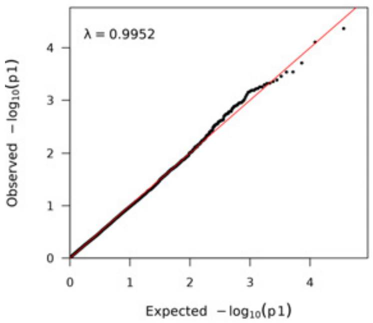
λ = 0.9952
Observed −log10(p1)
Expected −log10(p1)

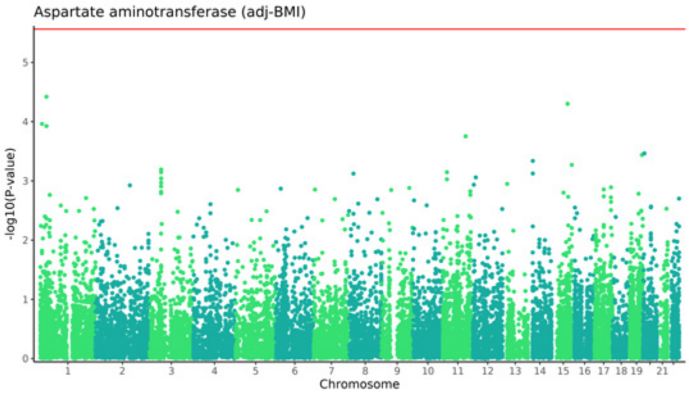
Aspartate aminotransferase (adj-BMI)
-log10(P-value)
Chromosome

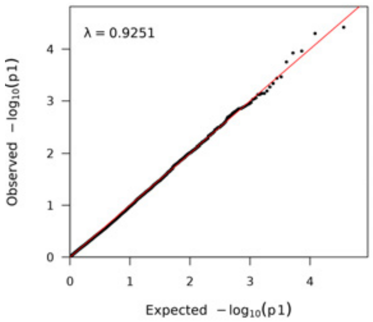
λ = 0.9251
Observed −log10(p1)
Expected −log10(p1)

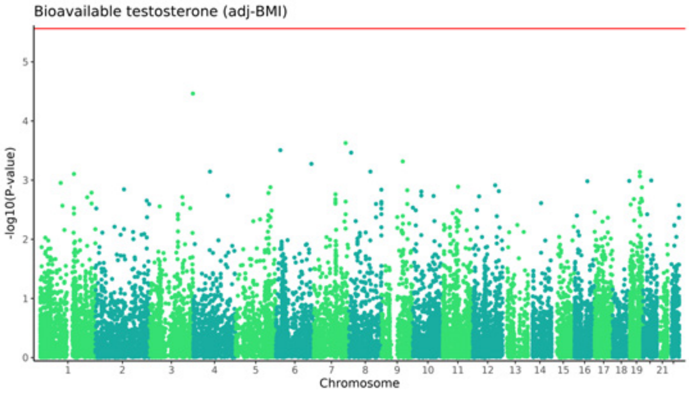
Bioavailable testosterone (adj-BMI)
-log10(P-value)
Chromosome

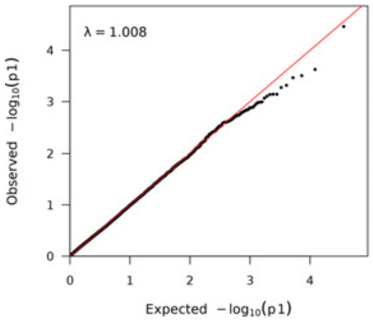
λ = 1.008
Observed −log10(p1)
Expected −log10(p1)

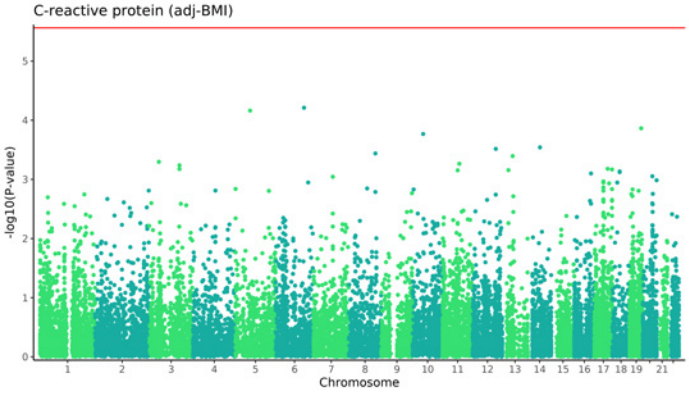
C-reactive protein (adj-BMI)
-log10(P-value)
Chromosome

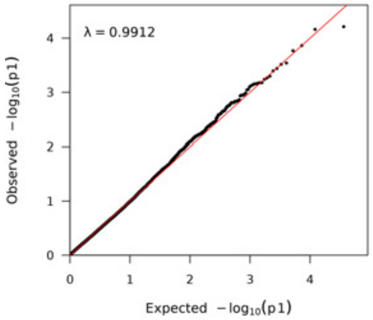
λ = 0.9912
Observed −log10(p1)
Expected −log10(p1)

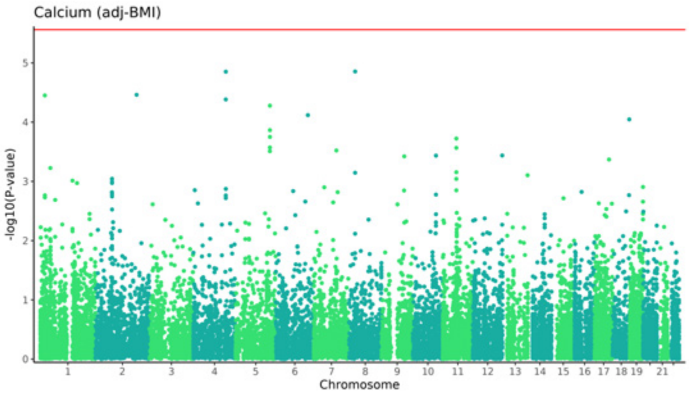
Calcium (adj-BMI)
-log10(P-value)
Chromosome

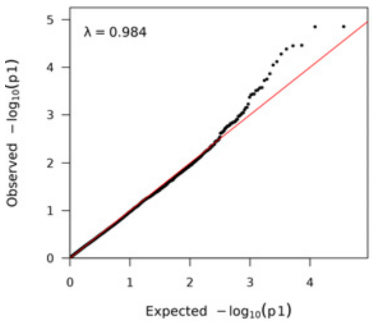
λ = 0.984
Observed $-\log_{10}(p1)$
Expected $-\log_{10}(p1)$

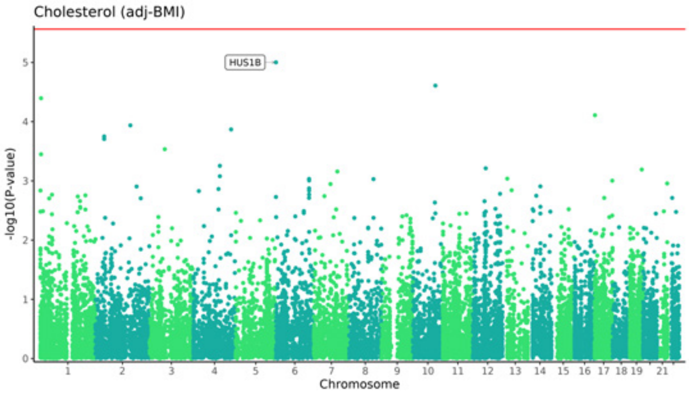
Cholesterol (adj-BMI)
HUS1B
-log10(P-value)
Chromosome

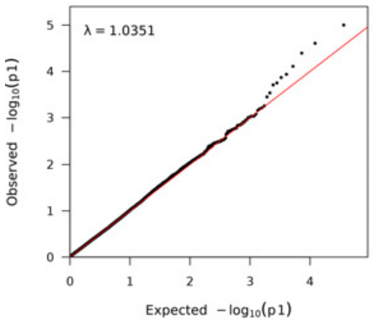
λ = 1.0351
Observed $-\log_{10}(p1)$
Expected $-\log_{10}(p1)$

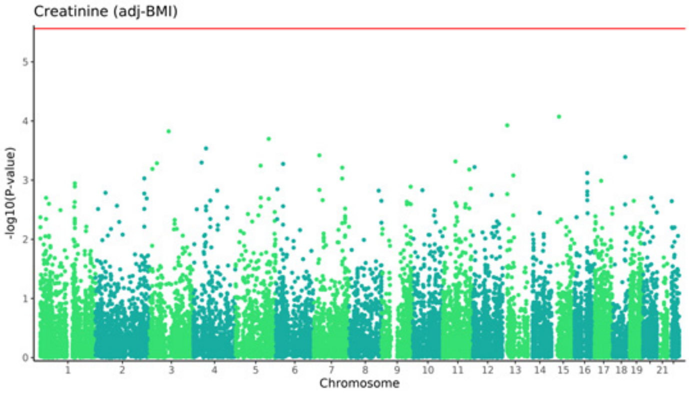
Creatinine (adj-BMI)
-log10(P-value)
Chromosome

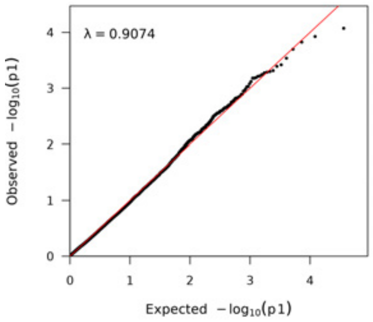
λ = 0.9074
Observed $-\log_{10}(p1)$
Expected $-\log_{10}(p1)$

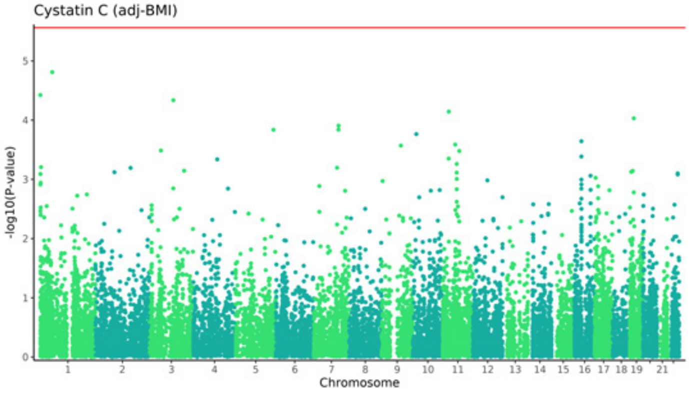
Cystatin C (adj-BMI)
-log10(P-value)
Chromosome

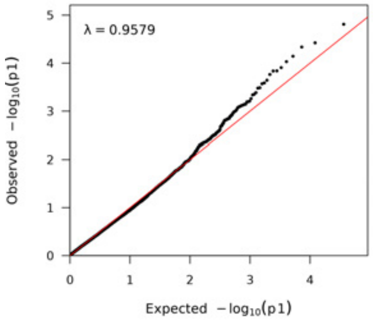
λ = 0.9579
Observed $-\log_{10}(p1)$
Expected $-\log_{10}(p1)$

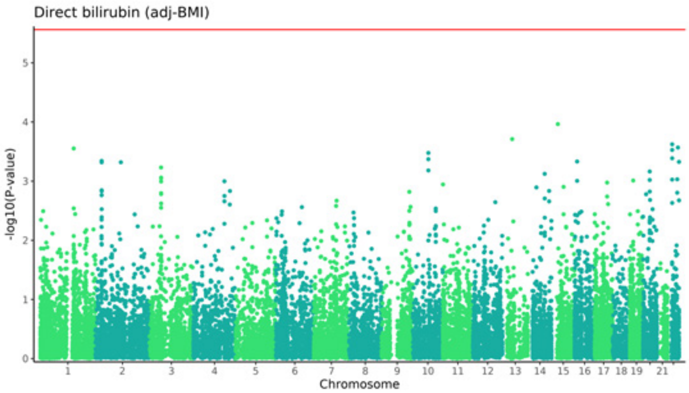
Direct bilirubin (adj-BMI)
-log10(P-value)
Chromosome

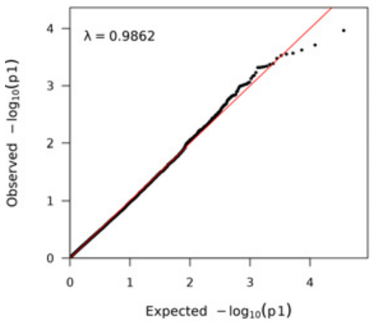
λ = 0.9862
Observed $-\log_{10}(p1)$
Expected $-\log_{10}(p1)$

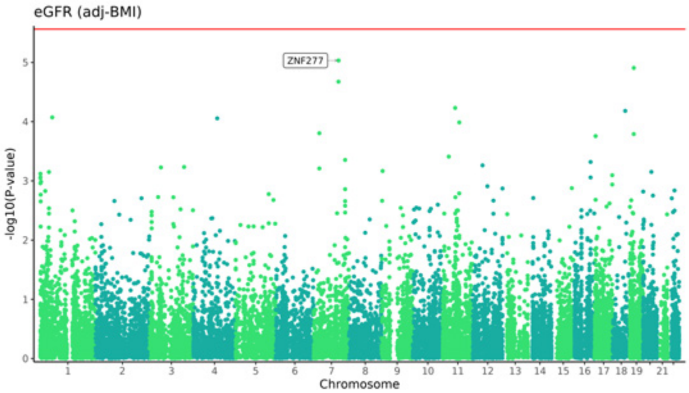
eGFR (adj-BMI)
ZNF277
-log10(P-value)
Chromosome

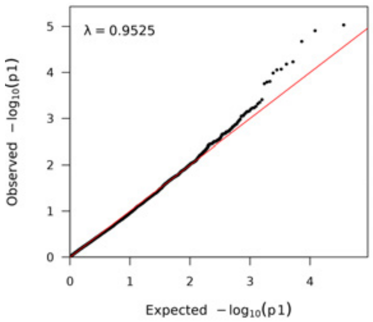
λ = 0.9525
Observed $-\log_{10}(p1)$
Expected $-\log_{10}(p1)$

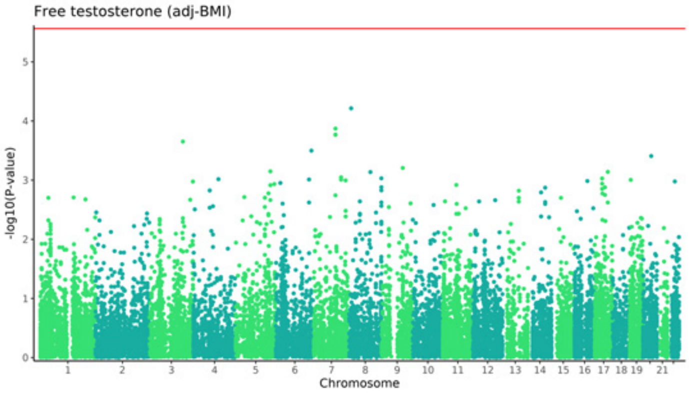
Free testosterone (adj-BMI)
-log10(P-value)
Chromosome

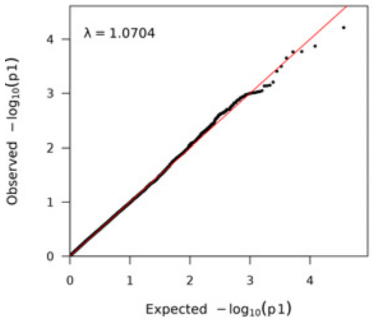
λ = 1.0704
Observed $-\log_{10}(p1)$
Expected $-\log_{10}(p1)$

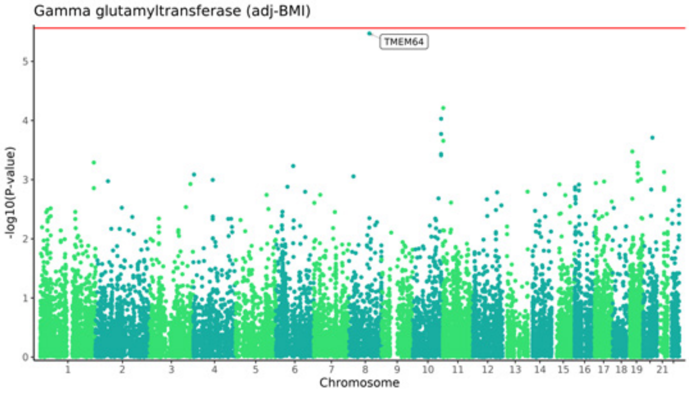
Gamma glutamyltransferase (adj-BMI)
TMEM64
-log10(P-value)
Chromosome

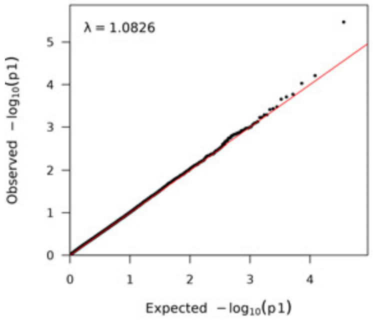
λ = 1.0826
Observed $-\log_{10}(p1)$
Expected $-\log_{10}(p1)$

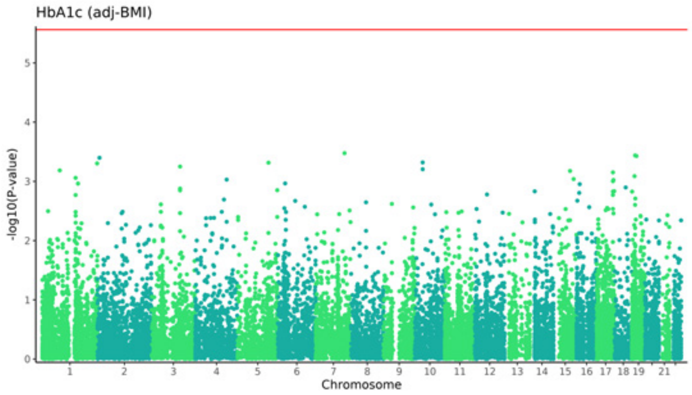
HbA1c (adj-BMI)
-log10(P-value)
Chromosome

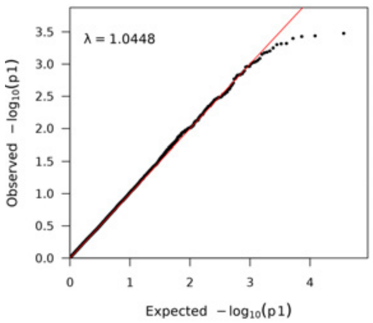
λ = 1.0448
Observed −log10(p1)
Expected −log10(p1)

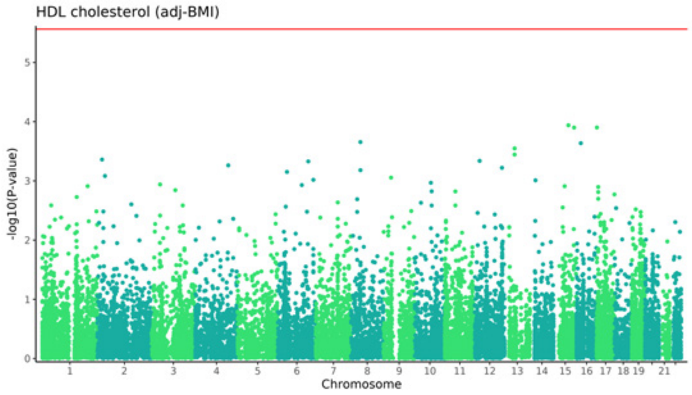
HDL cholesterol (adj-BMI)
-log10(P-value)
Chromosome

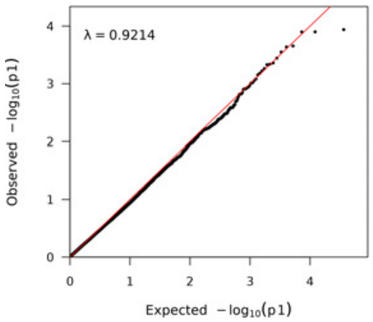
λ = 0.9214
Observed −log10(p1)
Expected −log10(p1)

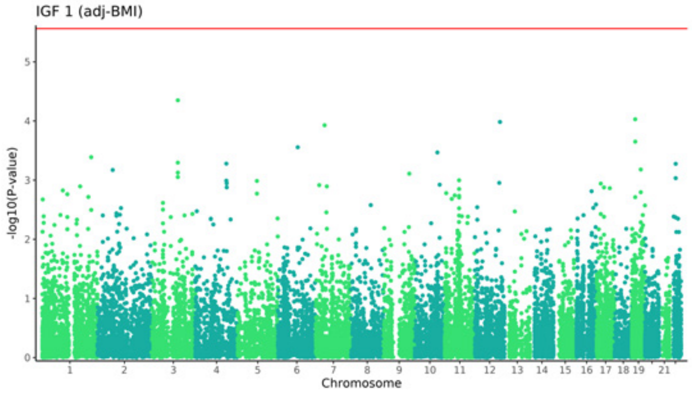
IGF 1 (adj-BMI)
-log10(P-value)
Chromosome

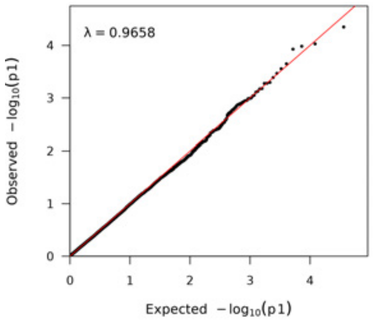
λ = 0.9658
Observed −log10(p1)
Expected −log10(p1)

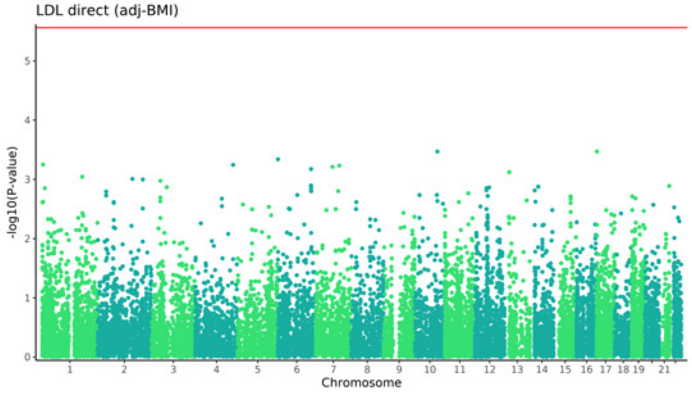
LDL direct (adj-BMI)
-log10(P-value)
Chromosome

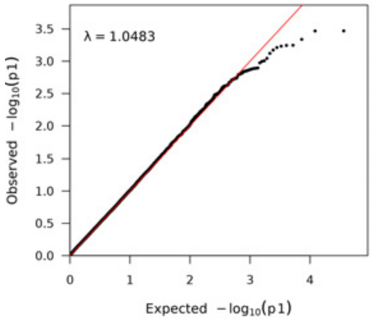
λ = 1.0483
Observed −log10(p1)
Expected −log10(p1)

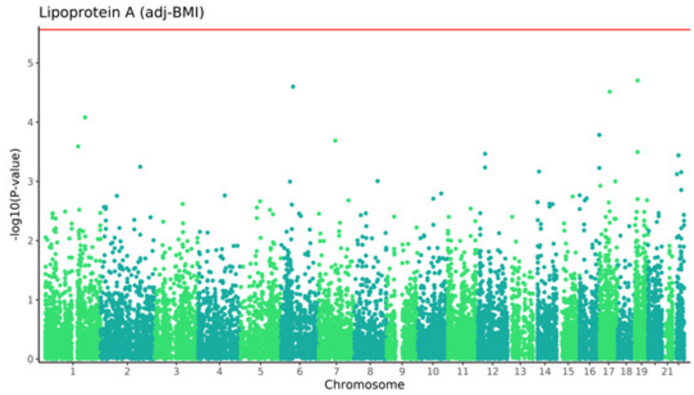
Lipoprotein A (adj-BMI)
-log10(P-value)
Chromosome

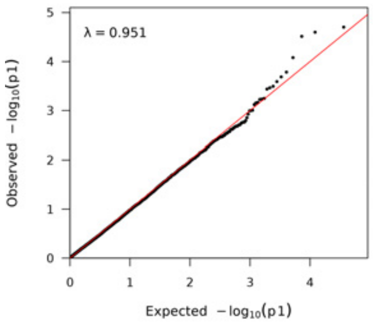
λ = 0.951
Observed $-\log_{10}(p1)$
Expected $-\log_{10}(p1)$

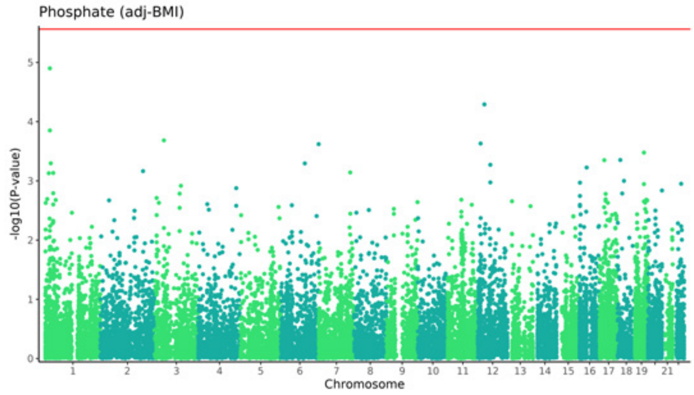
Phosphate (adj-BMI)
-log10(P-value)
Chromosome

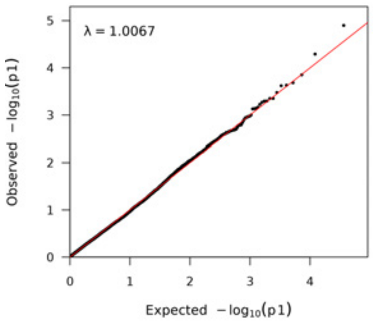
λ = 1.0067
Observed $-\log_{10}(p1)$
Expected $-\log_{10}(p1)$

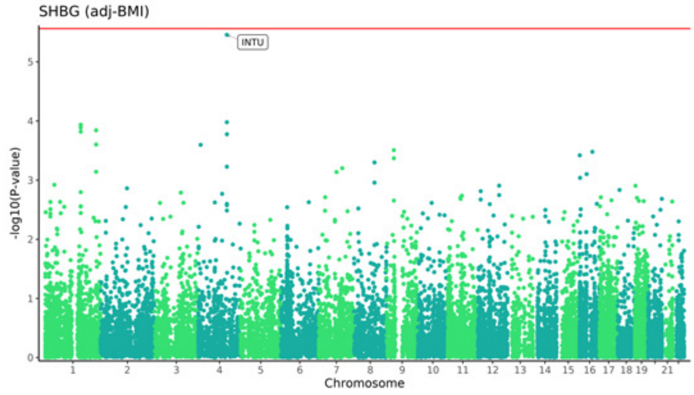
SHBG (adj-BMI)
INTU
-log10(P-value)
Chromosome

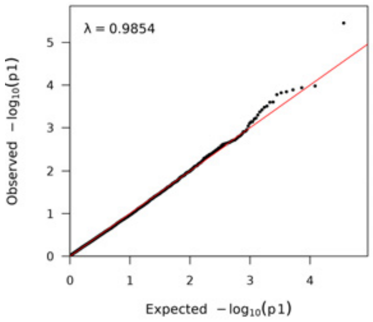
λ = 0.9854
Observed $-\log_{10}(p1)$
Expected $-\log_{10}(p1)$

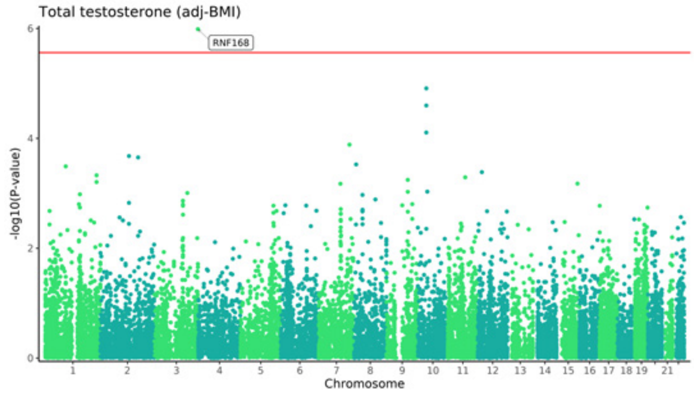
Total testosterone (adj-BMI)
RNF168
-log10(P-value)
Chromosome

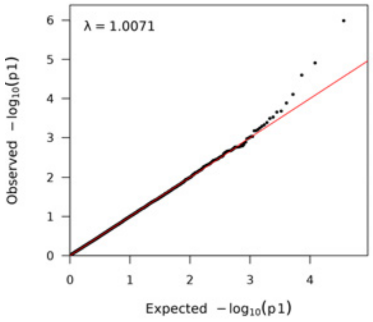
λ = 1.0071
Observed $-\log_{10}(p1)$
Expected $-\log_{10}(p1)$

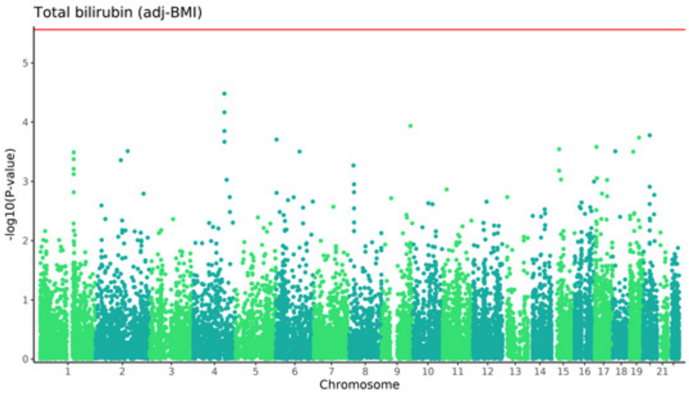
Total bilirubin (adj-BMI)
-log10(P-value)
Chromosome

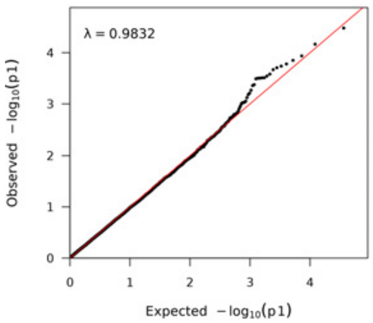
λ = 0.9832
Observed $-\log_{10}(p1)$
Expected $-\log_{10}(p1)$

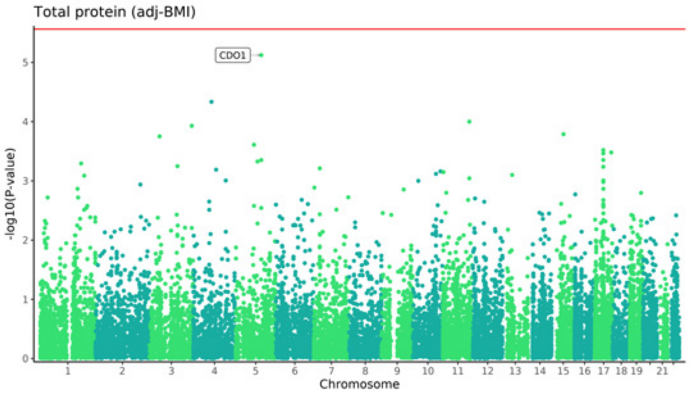
Total protein (adj-BMI)
CDO1
-log10(P-value)
Chromosome

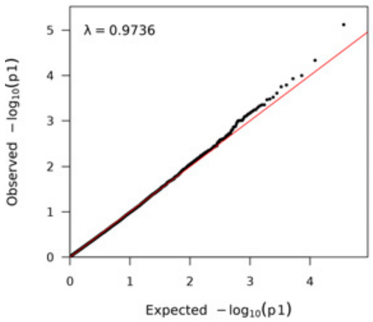
λ = 0.9736
Observed $-\log_{10}(p1)$
Expected $-\log_{10}(p1)$

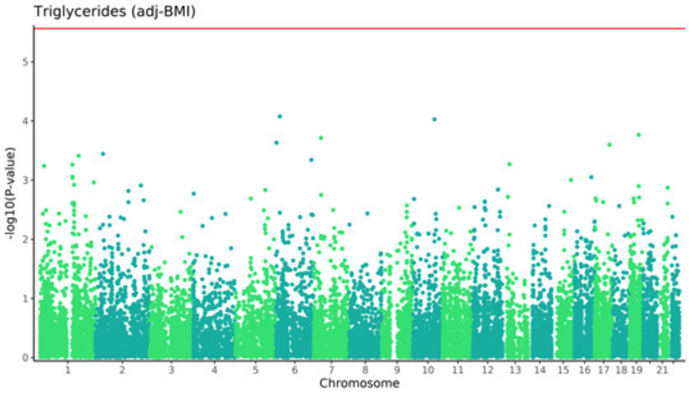
Triglycerides (adj-BMI)
-log10(P-value)
Chromosome

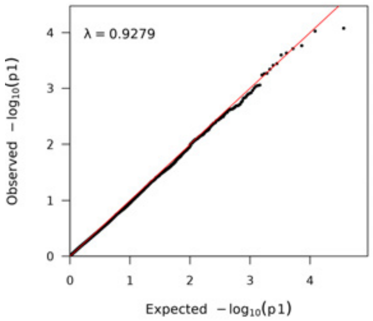
λ = 0.9279
Observed $-\log_{10}(p1)$
Expected $-\log_{10}(p1)$

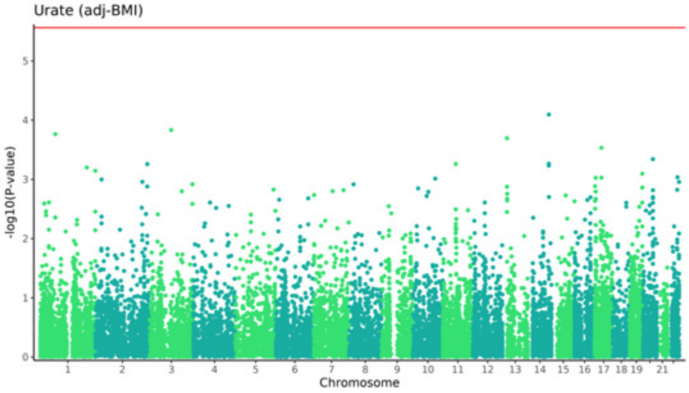
Urate (adj-BMI)
-log10(P-value)
Chromosome

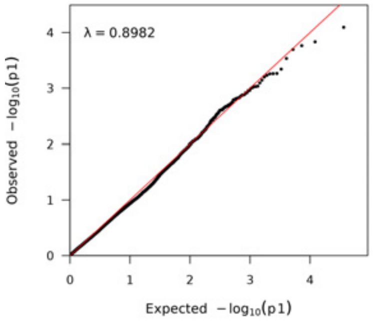
λ = 0.8982
Observed $-\log_{10}(p1)$
Expected $-\log_{10}(p1)$

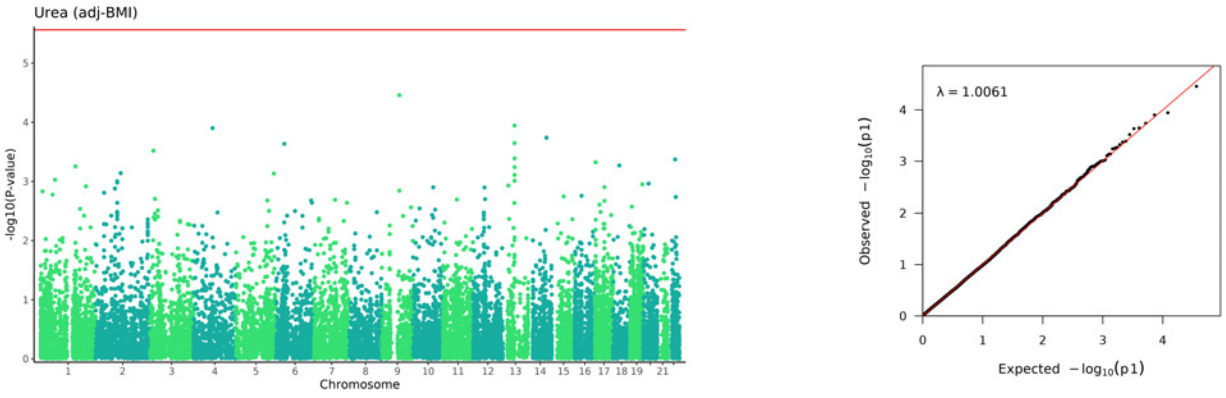
Urea (adj-BMI)
-log10(P-value)
Chromosome
1 2 3 4 5 6 7 8 9 10 11 12 13 14 15 16 17 18 19 21
λ = 1.0061
Observed −log10(p1)
Expected −log10(p1)

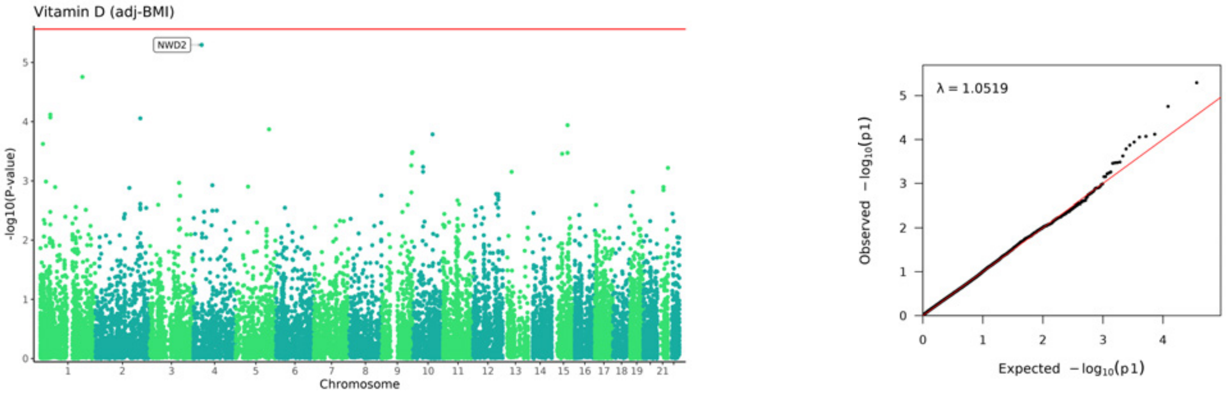
Vitamin D (adj-BMI)
NWD2
-log10(P-value)
Chromosome
1 2 3 4 5 6 7 8 9 10 11 12 13 14 15 16 17 18 19 21
λ = 1.0519
Observed −log10(p1)
Expected −log10(p1)

**S10**

Fish eating frequency of those who, on the same survey,
Answered 0 to:
"How old were you when you last ate any kind of meat?
(Enter "0" if you have never eaten meat in your lifetime)"
N=1230

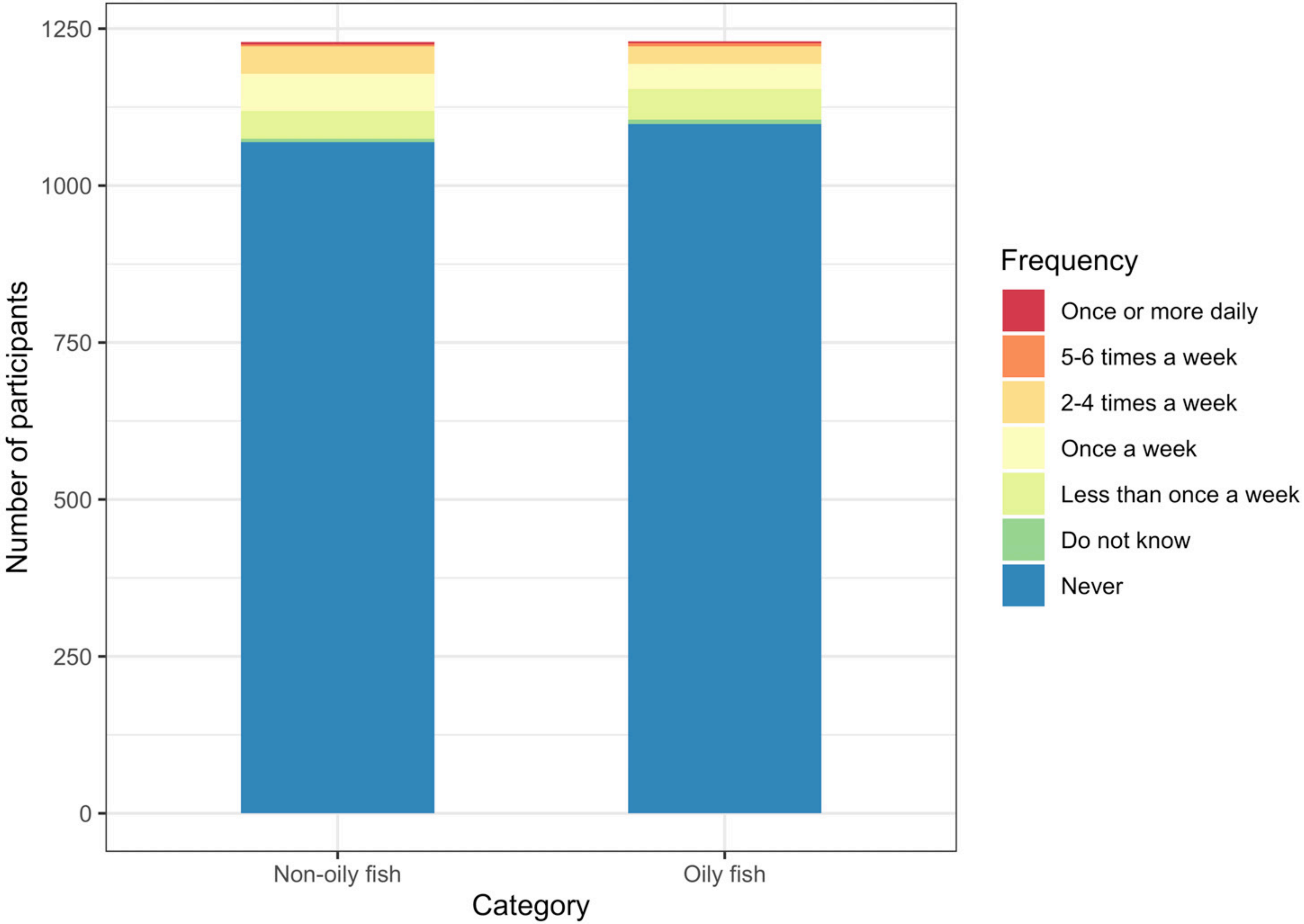
